# Association of country-wide coronavirus mortality with demographics, testing, lockdowns, and public wearing of masks (Update August 4, 2020)

**DOI:** 10.1101/2020.05.22.20109231

**Authors:** Christopher T. Leffler, Edsel Ing, Joseph D. Lykins, Matthew C. Hogan, Craig A. McKeown, Andrzej Grzybowski

## Abstract

**Purpose:** To determine sources of variation between countries in per-capita mortality from COVID-19 (caused by the SARS-CoV-2 virus).

**Methods:** Potential predictors of per-capita coronavirus-related mortality in 200 countries by May 9, 2020 were examined, including age, sex, obesity prevalence, temperature, urbanization, smoking, duration of infection, lockdowns, viral testing, contact tracing policies, and public mask-wearing norms and policies. Multivariable linear regression analysis was performed.

**Results:** In univariate analyses, the prevalence of smoking, per-capita gross domestic product, urbanization, and colder average country temperature were positively associated with coronavirus-related mortality. In a multivariable analysis of 196 countries, the duration of infection in the country, and the proportion of the population 60 years of age or older were positively associated with per-capita mortality, while duration of mask-wearing by the public was negatively associated with mortality (all p<0.001). International travel restrictions and a lower prevalence of obesity were independently associated with mortality in a model which controlled for testing policy. Internal lockdown requirements and viral testing policies and levels were not associated with mortality. The association of contact tracing policy with mortality approached statistical significance (p=0.06). In countries with cultural norms or government policies supporting public mask-wearing, per-capita coronavirus mortality increased on average by just 15.8% each week, as compared with 62.1% each week in remaining countries.

**Conclusions:** Societal norms and government policies supporting the wearing of masks by the public, as well as international travel controls, are independently associated with lower per-capita mortality from COVID-19.

## Introduction

The COVID-19 global pandemic caused by infection with severe acute respiratory syndrome coronavirus 2 (SARS-CoV-2) has presented a major public health challenge. For reasons that are not completely understood, the per-capita mortality from COVID-19 varies by several orders of magnitude between countries.^1^ Numerous sources of heterogeneity have been hypothesized. Higher mortality has been observed in older populations and in men.^2,3^ Patient-level behaviors, such as smoking, might also have an impact.^3^ Other potentially relevant factors include economic activity, and environmental variation, such as temperature.^4^ More urban settings and increased population density would be expected to enhance viral transmission.^5^

In addition, public health responses to the COVID-19 pandemic may influence per-capita mortality. Various strategies have been implemented, ranging from robust testing programs to lockdown or stay-at-home orders, to mandates regarding social distancing and face mask usage. Practices with theoretical benefit, such as social distancing, stay-at-home orders, and implementation of mandates regarding use of masks in public spaces, must be assessed quickly, as implementation has the potential to reduce morbidity and mortality.

Mask usage by the public is postulated to decrease infection by blocking the spread of respiratory droplets,^1^ and was successfully implemented during other coronavirus outbreaks (i.e. SARS and MERS).^6^ In the context of the ongoing pandemic, we assessed the impact of masks on per-capita COVID-19-related mortality, controlling for the aforementioned factors. We hypothesized that in countries where mask use was either an accepted cultural norm or favored by government policies on a national level, the per-capita mortality might be reduced, as compared with countries which did not advocate masks.

## Methods

### Data acquisition

In order to be included in the study, countries had to: 1) have coronavirus mortality data listed in the publicly available Worldometer Database on May 9, 2020;^7^ 2) have dates of first case and first death reported by the European Centre for Disease Prevention and Control (which did tabulate worldwide data);^8^ and 3) have an assessment of viral testing through May 9, 2020 by either: 3a) report on Worldometer of numbers of coronavirus PCR tests performed,^7^ or: 3b) testing and lockdown policies graded by the University of Oxford Coronavirus Government Response Tracker.^9,10^

Oxford University defined and scored several composite government response indices. The stringency index was defined in terms of containment policy and public information.^9^ The government response index incorporated containment, economic measures, public information, and testing and tracing policies.^9^ The containment and health index was defined in terms of containment measures, public information, and testing and tracing policies.^9^

Archived viral testing data for April 2020 were also downloaded.^11^ Mean temperature in each country during the pandemic was estimated using the average monthly temperature in the country’s largest city from public sources.^12,13^

Online news reports and government statements, including those cited by a previous review^14^ and a public database,^15^ were searched to identify countries in which the public wore masks early in the outbreak based on tradition, as well as countries in which the national government mandated or recommended mask-wearing by the public before April 16, 2020.

For each country, the population,^16^ fraction of the population age 60 years and over, and age 14 and under, male: female ratio per country,^17^ surface area,^16,17^ gross domestic product per capita,^18^ percent urbanization,^16,19^ adult smoking prevalence^20-23^ and prevalence of adult obesity^24-43^ were tabulated. Whether a nation was an isolated political entity on an island was also recorded.

### Statistical analysis

The prevalence of an infectious process undergoing exponential growth (or decay) appears linear over time when graphed on a logarithmic scale.^1^ Therefore, we postulated that the logarithm of the country-wide infection prevalence would be linearly related with the duration of the infection in each country. In addition, our analysis postulated that deaths from coronavirus would follow infections with some delay.

On average, the time from infection with the coronavirus to onset of symptoms is 5.1 days,^44^ and the time from symptom onset to death is on average 17.8 days.^45^ Therefore, the time from infection to death is expected to be 23 days.^1,46^ These incubation and mortality times were prespecified.^1,46^ Therefore, the date of each country’s initial infection was estimated as the earlier of: 5 days before the first reported infection, or 23 days before the first death.^8,11,47^ Deaths by May 9, 2020 would typically reflect infections beginning 23 days previously (by April 16). Therefore, we recorded the time from the first infection in a country until April 16. We also recorded the period of the outbreak: 1) from when public mask-wearing was recommended until April 16, 2) from the mandating of international travel restrictions or quarantine until April 16, and 3) from the start of mandated limits on internal activities (e.g. closures of schools or workplaces, limits on public gatherings or internal movement, or stay-at-home orders) until April 16. For countries scored by Oxford University, the Oxford data were used to determine the start of international travel restrictions and lockdowns on internal activity. In addition, we calculated the mean time-weighted score for each lockdown and testing policy as graded by the University of Oxford for the duration of the country’s outbreak, from beginning through April 16.^9^ For instance, if the school closure score was 1 for half the outbreak and 2 for the other half, then the mean score was 1.5.

Per-capita mortality can be analyzed as a binary outcome (low or high), or as a continuous variable. Each approach has strengths and weaknesses. Analysis of a binary outcome is not unduly influenced by outliers. Countries with extremely low or high mortality are included in the appropriate group, but the exact mortality value does not change the results. Moreover, analysis of a binary outcome facilities clear communication, because one can describe the characteristics of low and high mortality countries.

On the other hand, per-capita mortality is in fact a continuous variable, and the separation of countries just below or just above a threshold value is somewhat arbitrary, or susceptible to chance variation. Analysis of mortality as a continuous variable uses all the information available, and can appropriately model the exponential growth of an infection. We view the binary and continuous analyses as complementary. When one sees that a univariate association is found with both types of analysis, one gains confidence that the association is not an artifact of the analytic method selected.

In univariate analysis, characteristics of countries with above-median per-capita mortality were compared with the remaining (lower mortality) countries by the two-sample t-test using groups.

Significant predictors of per-capita coronavirus mortality in the univariate analysis were analyzed by stepwise backwards multivariable linear regression analysis. The dependent variable was the logarithm (base 10) of per-capita coronavirus-related mortality. Because of the importance relative to public health, the weeks the country spent in lockdown, with international travel restrictions, and using masks, and per-capita testing levels, were retained in the model. In addition, because of their biological plausibility and presumed importance, urbanization, prevalence of obesity, and average ambient temperature were retained in most of the multivariable models presented below. Statistical analysis was performed with xlstat 2020.1 (Addinsoft, New York). An alpha (p value) of 0.05 was deemed to be statistically significant. The study was approved by the Virginia Commonwealth University Office of Research Subjects Protection.

## Results

We studied coronavirus mortality in 200 countries, of which 183 had testing data,^7^ 169 had government policies scored by Oxford University,^9^ and 152 fell into both categories.

The 100 lower-mortality countries had 0.99 deaths per million population, in contrast with an average of 93.3 deaths per million population in the 100 higher-mortality countries (p<0.001, Table 1, Appendix Table A1). The median value was 4.0 deaths per million population.

**Table 1.** Characteristics of countries with low and high per-capita coronavirus mortality by May 9, 2020 in 200 countries.

|  | Mean (SD) |  | p value |
| --- | --- | --- | --- |
|  | Low Mortality | High Mortality |  |
| Deaths (per million) | 0.99 (1.14) | 93.3 (182.7) | <0.001 |
| Deaths (per capita, log) | -6.47 (0.75) | -4.55 (0.64) | <0.001 |
| Duration infection (weeks) | 6.51 (2.87) | 7.84 (2.31) | <0.001 |
| Duration infection without masks (weeks) | 4.74 (2.33) | 6.71 (2.34) | <0.001 |
| Time without international travel restrictions (weeks). | 1.44 (1.96) | 2.62 (2.38) | <0.001 |
| Duration infection without internal lockdown (weeks) | 1.79 (1.85) | 2.83 (2.08) | <0.001 |
| Temperature, mean (C) | 22.2 (7.6) | 14.1 (9.1) | <0.001 |
| Urban population (%) | 51.5 (22.6) | 70.4 (20.0) | <0.001 |
| GDP per capita (\$) | 9,060 (16,960) | 27,140 (27,500) | <0.001 |
| Age 14 & under (% of pop.) | 32.4 (9.8) | 20.2 (6.6) | <0.001 |
| Age 60 & over (% of pop.) | 8.8 (5.3) | 18.2 (7.9) | <0.001 |
| Surface area (million km<sup>2</sup>) | 0.563 (1.13) | 0.796 (2.44) | 0.39 |
| Population (million) | 51.9 (20.0) | 24.7 (48.0) | 0.19 |
| Prevalence males (%) | 50.1 (2.1) | 50.2 (4.2) | 0.95 |
| Smoking prevalence, adult (%) | 13.7 (7.9) | 18.4 (7.7) | <0.001 |
| Obesity prevalence, adult (%) | 14.6 (9.0) | 24.0 (7.3) | <0.001 |
| Tests per cap. (log) by Apr 4 | -3.73 (1.20) | -2.65 (0.76) | <0.001 |
| Tests per cap. (log) by Apr 16 | -3.09 (0.87) | -2.31 (0.67) | <0.001 |
| Tests per cap. (log) by May 9 | -2.76 (0.86) | -1.92 (0.62) | <0.001 |

Durations run from the estimated date of first infection in the country until 23 days before May 9, 2020 (i.e. April 16), or the stated event (mask recommendation or lockdown). Obesity data available for 196 countries. Testing data available for 135 countries by April 4, 162 countries by April 16, and 183 countries by May 9.

We assumed that island nations might find it less challenging to isolate and protect their populations. However, 19 of 100 low-mortality countries were isolated on islands, compared with 28 of 100 high-mortality countries (p=0.18). Country surface area and population were not associated with coronavirus mortality (Table 1).

### Population characteristics

Countries with older populations suffered higher coronavirus mortality. Countries with low mortality had on average 8.8% of their population over age 60, as compared with 18.2% in the high-mortality countries (p<0.001, Table 1). The proportion of the population which was male was not associated with country-wide mortality (p=0.95, Table 1). Smoking prevalence was on average 13.7% in low mortality countries and 18.4% in high-mortality countries (p<0.001, Table 1). The prevalence of obesity was on average 14.6% in low-mortality countries and 24.0% in high-mortality countries (p<0.001, Table 1).

### Temperature

Colder countries were associated with higher coronavirus mortality in univariate analysis. The mean temperature was 22.2 C (SD 7.6 C) in the low-mortality countries, and 14.1 C (SD 9.1 C) in the high-mortality countries (p<0.001, Table 1).

### Economics

Urbanization was associated with coronavirus mortality in univariate analysis. In low-mortality countries, on average 52% of the population was urban, as compared with 70% of the population in the high-mortality countries (p<0.001, Table 1). Richer countries suffered a higher coronavirus related mortality. The mean GDP per capita was $9,060 in the low-mortality countries, and was $27,140 in the high-mortality countries (Table 1, p<0.001).

### Masks: Early Adoption

The World Health Organization initially advised against widespread mask wearing by the public, as did the United States CDC.^1,48^ The WHO reversed course and recommended masks in public on June 5, 2020.^49^

Despite these initial recommendations, a number of countries did favor mask wear by the public early in their outbreak, and such countries experienced low coronavirus-related mortality (**Table 2**, Table A1, Figure 1).^50-68,S1-S301^ It is likely that in Mongolia and Laos, both of which reported no coronavirus-related mortality by May 9, the public began wearing masks before any cases were confirmed in their countries (Table 2). We identified 22 additional countries with recommendations or cultural norms favoring mask-wearing by the public within 20 days of the estimated onset of the country’s outbreak:^1^ including (beginning with those favoring masks earliest in the course of their outbreak): Japan, the Philippines, Macau, Hong Kong, Sierra Leone, Cambodia, Timor-Leste, Vietnam, Malaysia, Bhutan, Venezuela, Taiwan, Slovakia, St. Kitts and Nevis, South Korea, Indonesia, Brunei, Grenada, Mozambique, Uzbekistan, Thailand, and Malawi (Table 2). The average mortality by May 9 for these 24 early mask-wearing countries was 1.5 per million (SD 2.0). Twenty of the 24 were lower-mortality countries (p=0.001).

**Table 2.** Countries in which masks were widely used by the public or recommended by the government within 31 days of the estimated local onset of the outbreak, by timeliness of mask-wearing.

|  | Mask Delay (days) | First Case Date | Mortality (per mil.), by May 9. | Comment. |
| --- | --- | --- | --- | --- |
| Mongolia | 0 | Mar. 10 | 0.0 | The public began wearing masks in January.<sup>S183</sup> The mayor of Ulaanbaatar ordered organizations to implement mask-wearing on January 27, 2020.<sup>S184</sup> Public mask wearing was quite common by mid-February when the government encouraged mask usage by denying service on transport for those not wearing masks.<sup>50</sup> |
| Laos | 0 | Mar. 24 | 0.0 | Health officials in Laos advised mask-wearing by March 6,<sup>S156</sup> and the public began wearing masks even before any cases were reported in the country.<sup>S157</sup> |
| Japan | 5 | Jan. 16 | 4.8 | Public use of masks is traditional.<sup>48</sup> Surveys indicate that 64% of adults habitually wore a mask in Winter.<sup>51</sup> Public masking was manifest by Jan. 16 when the first domestic case was announced.<sup>S144-S146</sup> The government initially recommended masks when in “confined, badly ventilated spaces”.<sup>48</sup> One survey documented mask wear prevalence over 60% by March 14, increasing to over 75% by April 12.<sup>52</sup> In another poll, 62% indicated wearing a mask in public by March 17, 76% by April 13, 81% by April 20, and 86% by May 4, 2020.<sup>53</sup> |
| Philippines | 5 | Jan. 30 | 6.4 | Masks were used extensively as early as Jan. 30.<sup>S215</sup> In a poll, 60% indicated wearing a mask in public on Feb. 24, 76% on March 23, and 81% to 84% from March 30 through June 22.<sup>53</sup> Masks were mandated on April 2. |
| Macau | 6 | Jan. 22 | 0.0 | Mask use is traditional. By Jan. 23, the government had implemented a mask distribution program for the public.<sup>S169</sup> |
| Hong Kong | 6 | Jan. 23<sup>S121</sup> | 0.5 | Surgical masks were traditionally used, and also were recommended on public transport and in crowded places, on January 24, 2020. <sup>48,S120</sup> Surveys indicated that masks were worn by about 73% in the week of Jan. 21, and by 98% of the public by mid-February, which persisted into May. <sup>S122</sup> In February 2020, 94.8% of pedestrians were observed to wear masks, and 94.1% believed mass masking reduces the chance of community outbreak. <sup>54</sup> A poll consistently found that 85% or more wore masks in public between Feb. 25 and June 22, 2020. <sup>53</sup> |
| Sierra Leone | 6 | Mar. 31<sup>S236</sup> | 2.3 | Masks were recommended in public on April 1.<sup>S237</sup> Compliance has been incomplete.<sup>S238</sup> |
| Cambodia | 6 | Jan. 27<sup>S44</sup> | 0.0 | Masks were widely used by the public by January 28.<sup>S45,S46</sup> |
| Timor-Leste | 7 | Mar. 21<sup>S270</sup> | 0.0 | Masks were required in stores and other venues as part of a state of emergency beginning March 28.<sup>S271</sup> |
| Vietnam | 9 | Jan. 23 | 0.0 | Masks were widely used by the public by January 27,<sup>S295,S296</sup> and were mandated by the government on March 16. One survey found the prevalence of mask wear consistently from 85-90% from March 12 to April 14.<sup>52</sup> A poll reported 59% wore a mask on March 23, and between 79% and 87% from March 30 to June 8.<sup>53</sup> From March 31 to April 6, 2020, 99.5% of respondents reported using a mask when outside.<sup>55</sup> |
| Malaysia | 10 | Jan. 25 | 3.3 | Masks were used by the public by January 30.<sup>S173</sup> A poll reported 55% wore a mask in public on Feb. 24, 69% on Mar. 23, 82% on Apr 6, and 85-88% from May 4 to June 8.<sup>53</sup> |
| Bhutan | 10 | Mar. 6<sup>S31</sup> | 0.0 | On Mar. 11, the Ministry of Health advised wearing of masks in “a crowded place”.<sup>S32</sup> |
| Venezuela | 10 | Mar. 13<sup>S291</sup> | 0.4 | The first death was announced on March 26.<sup>S293</sup> President Maduro demonstrated wearing of masks on live television on March 13 (the day the first case was confirmed), and required masks on public transport.<sup>S290,S291</sup> Masks were required in any public space by March 17.<sup>S292,S293</sup> |
| Taiwan | 11 | Jan. 21 | 0.3 | Use of masks is traditional. By January 24, Taiwan banned the export of surgical masks.<sup>56,57</sup> By January 27, the government had to limit mask exports and limit sales from pharmacies to those needed for personal use.<sup>S265</sup> On January 28, the government began releasing 6 million masks daily, with each resident able to purchase 3 masks weekly at a set price.<sup>56</sup> A poll consistently found over 80% wore a mask from Feb. 25 to June 22, 2020.<sup>53</sup> |
| Slovakia | 13 | Mar. 7 | 4.8 | Masks were mandated in shops and transit on March 15,<sup>S243</sup> and more broadly in public on March 25.<sup>S244</sup> |
| St. Kitts and Nevis | 14 | Mar. 24<sup>S223</sup> | 0.0 | On April 2, Chief Medical Officer Dr. Hazel Laws recommended wearing a mask in public on the grounds that masks could block droplets, and viral particles could remain suspended for 3 hours. <sup>S224</sup> The requirement to wear masks in public became mandatory on April 7. <sup>S225</sup> |
| South Korea | 15 | Jan. 20 | 5.0 | Use of masks is traditional.<sup>48</sup> The alert level was raised from yellow to orange on Jan. 27.<sup>58</sup> Children were advised to wear masks at school by January 30.<sup>S249</sup> By Feb. 2, mask sales increased 373 times year-over-year.<sup>58</sup> Stores were selling out of masks by February 3.<sup>S250</sup> A superspreader event in mid-February was associated with a religious group which did not use masks at their gatherings.<sup>59</sup> South Korea initially had trouble obtaining enough masks, but at the end of February the government began to control the distribution of masks to the public.<sup>S251</sup> On Feb. 22, the government instructed the wearing of masks in the epidemic area.<sup>58</sup> |
| Indonesia | 15 | Mar. 2<sup>S127</sup> | 3.5 | The first death occurred on March 3.<sup>S128</sup> The public scrambled to buy face masks in early February.<sup>S126</sup> The proportion of Indonesian adults wearing a mask in public was 54% on Feb. 24, 2020, 47% on March 9, 59% on March 23, 71% on March 30, 79% on April 13, 81% on April 20, and from 82%-84% from May 4 to June 9.<sup>53</sup> During March and April, 76% of students indicated that they wore a mask outside the home.<sup>60</sup> Masks were mandated in public on April 5.<sup>S129</sup> |
| Brunei | 18 | Mar. 9<sup>S39</sup> | 2.3 | On March 22, Sultan Hassanal Bolkiah advised the people to wear masks in public.<sup>S40</sup> |
| Grenada | 18 | Mar. 21<sup>S110</sup> | 0.0 | On April 3, the Ministry of Health recommended all wear a mask, which could be purchased at a pharmacy, to “prevent asymptomatic people from transmitting the disease unknowingly”.<sup>S111</sup> Masks were mandated outside the home on April 6.<sup>S112</sup> |
| Mozambique | 18 | Mar 22<sup>S188</sup> | 0.0 | Masks were recommended by health authorities on April 4,<sup>S189</sup> and were required on public transport or in gatherings on April 8.<sup>S190</sup> |
| Uzbekistan | 19 | Mar. 15<sup>S288</sup> | 0.3 | The first coronavirus death was on March 29. Masks were mandated on March 25.<sup>S289</sup> |
| Thailand | 20 | Jan. 13 | 0.8 | Masks, including N95 masks, were already worn outdoors in early January to combat smog. The Thai government was handing out masks and advising wearing of masks in public to prevent coronavirus by January 28, 2020.<sup>S266-S269</sup> The recommendation of cloth masks for the public was reaffirmed by the |
|  |  |  |  | Ministry of Public Health on March 3, 2020.<sup>61</sup> Enforcement of a mask mandate on public transport began on March 26.<sup>61</sup> One survey reported high mask-wearing: 73% by Feb. 24, 80% by March 23, and between 84 and 89% between March 30 and June 22.<sup>53</sup> During March 2020, another survey found masks were worn “all the time” by 14% of COVID-19 cases and 24% of controls, and “some of the time” by 38% of cases and 15% of controls.<sup>61</sup> |
| Malawi | 20 | Apr. 2 | 0.2 | The first death was on April 7.<sup>S171</sup> The public was required to wear masks on April 4.<sup>S172</sup> A survey in Karonga from April 25 to May 23 found that 22% of urban residents and 5% of rural residents wore a mask.<sup>62</sup> |
| São Tomé and Príncipe | 21 | Apr. 6<sup>S230</sup> | 22.8 | On April 22, it was announced that masks would be mandatory in public beginning April 24.<sup>63</sup> |
| Czechia | 23 | Mar. 1<sup>S70</sup> | 25.8 | Masks were required in public on March 19.<sup>S71</sup> |
| Dominica | 23 | Mar. 22<sup>S73</sup> | 0.0 | Prime Minister Skerit and Health Minister McIntyre wore masks during an interview on March 30.<sup>S75</sup> When Dr. Adis King demonstrated mask-wearing to the legislative assembly on April 7, all in attendance wore masks.<sup>S76</sup> President Savarin recommended the wearing of masks in public on April 9.<sup>S74</sup> Others,<sup>S79</sup> including the state epidemiologist,<sup>S80</sup> repeated this recommendation in coming days. On April 21, physician Sam King estimated that 95% of the population was wearing masks in public.<sup>S78</sup> Masks were mandated on public transport on April 25.<sup>S77</sup> |
| Bangladesh | 24 | Mar. 8<sup>S18</sup> | 1.3 | The first death occurred on March 18.<sup>S19</sup> From March 11-19, 2020, when students age 17 to 28 were asked if they were wearing a surgical face mask in public, 53.8% responded “yes” and an additional 6.6% responded “occasionally”.<sup>64</sup> A survey from March 29 to April 29 found that 98.7% reported wearing a face mask in crowded places.<sup>65</sup> |
| Zambia | 24 | Mar. 18 | 0.4 | The first death was recorded on April 2. On April 4, masks were recommended for the public “at all times” by the Zambian Minister of Health.<sup>S298</sup> This spurred the manufacture of cloth masks.<sup>S299</sup> On April 16, masks were mandated for the public.<sup>S300</sup> |
| Chad | 24 | Mar. 19<sup>S52</sup> | 1.9 | On April 13, the office of the president announced that a mask or suitable alternative (e.g. turban, veil) would be mandatory in public on April 14. <sup>66,S53</sup> On April 14, the government had to backtrack on enforcement due to lack of supplies. <sup>S56</sup> Specific penalties for failing to wear a mask in public were announced on May 7. <sup>S54</sup> |
| Benin | 26 | Mar. 16<sup>S27</sup> | 0.2 | Masks were recommended in public on April 6,<sup>S28</sup> mandated on April 7,<sup>S29</sup> and enforced by police beginning April 8.<sup>S30</sup> |
| Sudan | 27 | Mar. 12 | 1.5 | The first death occurred on March 12. Masks were dispensed by pharmacists for free in Sudan by March 16.<sup>S261,S262</sup> A survey from March 25 to April 4 of 2336 adults found that 703 (30.1%) had been to a crowded area, and 1153 (49.4%) had worn a mask outside the home in the previous few days.<sup>67</sup> |
| El Salvador | 27 | Mar. 18<sup>S89</sup> | 2.6 | The first death was reported March 31. President Bukele recommended universal mask wear in public on April 4.<sup>S90</sup> Masks were mandated in San Salvador on April 7.<sup>S91</sup> On April 11, the president announced a nationwide mask mandate, effective April 14.<sup>S92</sup> |
| Antigua and Barbuda | 28 | Mar. 13<sup>S9</sup> | 30.6 | Masks were required in all public spaces on April 5.<sup>S10</sup> |
| Myanmar | 28 | Mar. 23<sup>S191</sup> | 0.1 | In Myanmar, the first death occurred on March 31.<sup>S192</sup> A study from March 3-20, 2020 found that 72% of adults were confident they would wear a surgical mask whenever visiting a crowded area.<sup>68</sup> On April 5, the Ministry of Health recommended masks in crowded places, and cited the US CDC recommendation for the use of cloth masks by the public.<sup>S194</sup> On April 7, State Counsellor Daw Aung San Suu Kyi announced that she would make a mask for herself.<sup>S195</sup> By April 16, some regions mandated masks in public.<sup>S196</sup> A survey from May 7-23, 2020 conducted by the Ministry of Health found that 80% of the public wore a mask each time they went out.<sup>S193</sup> |
| Bosnia and Herzegovina | 29 | Mar. 5<sup>S33</sup> | 31.1 | Masks were required in public by March 29.<sup>S34,S35</sup> |
| Côte d'Ivoire | 29 | Mar. 11<sup>S140</sup> | 0.8 | On April 4, senior health officials recommended masks when in public.<sup>66,S141</sup> |
| South Sudan | 29 | Apr. 5<sup>S252</sup> | 0.0 | On April 29, the High Level Task Force approved the use of locally-manufactured cloth masks to be worn in public.<sup>S253</sup> |
| Kenya | 30 | Mar. 12<sup>S150</sup> | 0.6 | The March 12 case had arrived from the U.S. on March 5.<sup>S150</sup> The first death was on March 26, of a man who arrived in Kenya on March |
|  |  |  |  | 13.<sup>S151</sup> Masks were mandated in Kenya on public transport on April 2, and more broadly in public on April 4.<sup>S152,S153</sup> A survey in Nairobi published on May 5, 2020 found that 89% had worn a face mask in the previous week, and 73% said they always did so outside the home.<sup>S154</sup> |
| Saint Lucia | 30 | Mar. 13<sup>S226</sup> | 0.0 | Face masks were recommended to be worn when shopping by the chief medical officer on April 7.<sup>S227,S228</sup> |
| Barbados | 30 | Mar. 17<sup>S20</sup> | 24.4 | By April 11, cloth face masks were required when shopping.<sup>S21-S23</sup> Masks were mandatory on buses by May 11.<sup>S24</sup> |

The delay was the number of days from the start of the outbreak until masks were recommended by the government or became widespread due to cultural norms. The estimated start of the outbreak was 5 days before the first infection was reported, or 23 days before the first death (whichever was first).

**Figure 1.**
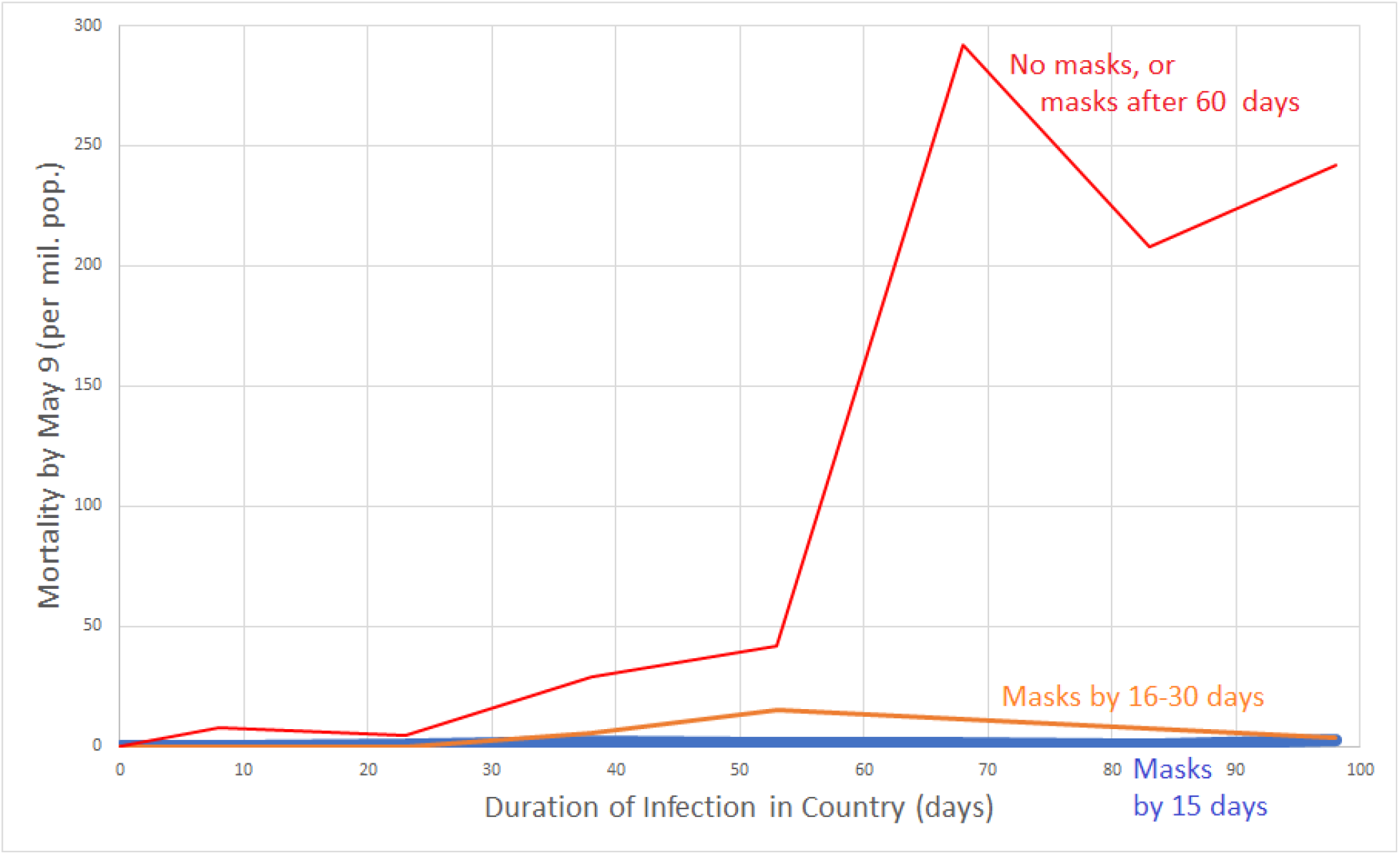
Per-capita mortality by May 9 versus duration of infection according to whether early masking was adopted. Data grouped by whether country did not recommend masks by April 16, 2020 or recommended them more than 60 days after outbreak onset (red line); recommended masks 16 to 30 days after onset of the country’s outbreak (orange line); or recommended masks (or traditionally used masks) within 15 days of the outbreak onset (blue line close to the x-axis). Country mortality was averaged for the following country groups of infection duration: 0-15 days, 16-30 days, 31-45 days, 46-60 days, 61-75 days, 76-90 days, 91-105 days. For instance, per-capita mortality for all non-mask or late-masking countries with infection duration between 61 and 75 days was averaged, and graphed at the x-value 68 days. Data for graph derived from 200 countries.

An additional 17 countries recommended that the public wear masks within 30 days of the estimated onset of their outbreak: São Tomé and Príncipe, Czechia, Dominica, Bangladesh, Zambia, Chad, Benin, Sudan, El Salvador, Antigua and Barbuda, Myanmar, Bosnia and Herzegovina, Côte d’Ivoire, South Sudan, Kenya, Saint Lucia, and Barbados (Table 2). The average mortality by May 9 for this group was 8.5 per million (SD 12.4).

### Masks in Asia

Throughout much of East, South, and Southeast Asia, masks were worn by the public as a preventive measure, rather than a policy implemented after evidence emerged of health system overload (Table 2). The public sometimes implemented masks before government recommendations were issued.

As the country where the pandemic started, China is a noteworthy case of a nation which traditionally has favored mask-wearing by the public for respiratory illnesses, but which did not deploy masks immediately. The first cases in China had begun by December 1, 2019.^69^ By the time human-to-human transmission was confirmed on Jan. 20, 2020, many in Beijing were already wearing masks.^S58^ The government required masks in public in Wuhan on Jan. 22.^S59^ From Jan. 23-25, thirty regions in China mandated masks in public.^58,70^ Masks were ordered throughout China when around others in public on Jan. 31.^S60^ China suffered a very significant outbreak in Wuhan, but appears not to have experienced the same level of infection in other regions. Surveys indicate that the prevalence of public mask wear in China remained between 82% and 90% between February 24 and June 22.^53^ Another survey confirmed mask wear from 80-90% from March 12 to April 14.^52^ The reported country-wide per-capita mortality by May 9, 2020 was 3.2 per million population.

For several countries in South or Southeast Asia with mortality lower than in the West, we did not score the country as mask-wearing in the primary analysis until their governments issued recommendations to do so. Nonetheless, there is evidence of significant mask wear by the public before the recommendations in Nepal, India, and Sri Lanka.

In Nepal, facemasks are commonly seen in urban centers due to air pollution.^71^ The first case of COVID-19 in the country was reported on January 13, in a traveler returning from Wuhan.^72^ However, no subsequent cases were reported in Nepal until the second week of March.^72^ By January 29, all students at some schools were wearing masks.^S198^ By February 3, pharmacies were selling out of masks due to increased demand.^S199^ With the outbreak, tailors began sewing cloth masks.^71^ By February 8, 2020, “a majority” of the public was wearing masks.^S200^ The recommendation to wear masks in public became more formalized on March 25.^S201^ The Ministry of Health distributed masks to children and elderly in shelters by March 25.^S202^ Surveys in Nepal found that 83% of respondents agreed that asymptomatic people should wear masks to prevent COVID-19 infection at the end of March,^72^ and 96% agreed with this statement from May 15 to June 20.^73^ As of May 9, Nepal reported no coronavirus-related mortality. We used the March 25 recommendation as the date in the mask analysis, but earlier mask use might have forestalled the epidemic in Nepal.

In India, the first case of coronavirus was diagnosed on January 30.^S124^ The Health Ministry recommended homemade face masks on April 4, 2020.^S125^ However, mask wear was high both before and after the recommendation. According to one poll, masks were worn by 60% of the public from March 12-14, 67% from March 19-21, and then from 73% to 76% between March 26 through April 12.^52^ According to another poll, masks were worn by 43% of the public on March 16, 46% on March 20, 65% on March 27, 71% on April 3, 79% on April 10, and 81-84% between April 17 and May 1.^53^ A survey conducted in March 2020 found that 75% of the public believed that masks should be worn even by asymptomatic people, and 77% of respondents indicated that the N95 mask was most protective.^74^ By May 9, the per-capita mortality was 1.5 per million.

In Sri Lanka, the public immediately bought masks at the end of January when the first cases were identified.^S257^ Masks were mandated in public on April 11.^S258^ The per-capita mortality by May 9 was 0.4 per million.

Singapore was slower than its Asian neighbors to embrace masks, but when the government shifted course, the public was ready to respond. On March 27, only 27% of respondents indicated that they wore a mask.^53^ On April 3, when the government announced that it would no longer discourage mask-wearing by the public, and would instead distribute masks,^S239-S241^ 37% indicated that they wore a mask.^53^ Mask wearing by the public reached 73% on April 10, 85% on April 17, and 90% on April 24, where it remained through June 19. ^53^

Early in the pandemic, masks were noted to be “somewhat common” in Afghanistan.^75^ By March 29, 2020, the Taliban had begun distributing masks to the public in areas under their control.^S1^

In March 2020, 78% of Pakistanis in Sargodha were in favor of wearing a mask to prevent coronavirus.^76^ Another survey conducted from April 1-12 indicated that 80% of Pakistanis believed the government should mandate mask wearing for adults outside the home.^77^ Masks were mandated when in crowded spaces in Pakistan on May 31.^S209^

### Masks in the Middle East

In parts of the Middle East, masks were embraced by the public even before government requirements. In the United Arab Emirates, the first cases were reported on January 29.^S280^ By February 29, mask usage had become “more prominent”, but the Ministry of Health and Community Protection advised that N95 masks should be reserved for medical personnel treating coronavirus patients, and could cause “respiratory illness” if worn by the public.^S281^ Despite this warning, a poll of UAE residents found that masks were worn by 39% of the public on March 18, and 44% on March 25.^53^ On March 27, the government followed the people’s lead, and mandated masks when indoors.^S282^ Subsequently, masks were worn by 63% on April 1 and between 78% and 81% between April 14 and June 17.^53^ By May 9, the per-capita mortality was 18.7 per million.

In Saudi Arabia, the first case was announced on March 2.^S231^ A poll of Saudi residents found that 35% wore a mask on March 18, 54% on April 1, and 59% on April 14,^53^ despite the lack of any official guidance to do so. A different survey conducted from April 2-5, during a period of lockdown, found that 16.9% had worn a mask even without symptoms.^78^ Public mask-wearing was recommended by the Saudi government on April 28,^S232^ and mandated on May 30.^S233^ Mask-wearing reached 63% on May 4, and 72% on June 3.^53^ A survey of Saudi nursing students which concluded on June 30 found that 87% had worn a mask when going out in recent days.^79^ By May 9, the per-capita mortality was 6.9 per million.

In Lebanon, the first case was reported on February 21.^S159^ Masks were popular among the public from mid-March to early April.^S160,S161^ Masks were recommended by the health minister on April 25.^S161^ By May 9, the per-capita mortality was 3.8 per million.

In March 2020 in Egypt, 76.4% of adults expressed an understanding of the value of wearing a mask in public, but only 36.4% agreed that they actually did so.^80^ At this time, the government was not mandating masks, but by March 20, prices of masks had soared, and volunteer organizations were advocating public masking in Egypt.^S87^ Masks were mandated in public in Egypt on May 31.^S88^

In Iran, no infections were announced until February 19, when two deaths were reported.^S130^ By March 12, satellite imagery demonstrated the digging of mass graves in Qom.^S131^ In accord with WHO guidelines, the guidance of the Iranian Health Ministry available on March 24, 2020 advised that the public wear a mask only if symptomatic or caring for the sick (personal communication, Linnea I. Laestadius, June 7, 2020).^14^ However, a new guidance which recommended universal masking in gyms, parks, and public transit was issued by the Ministry by March 29,^14^ an estimated 62 days after the start of the outbreak (assuming the reported deaths were really the first). A survey conducted from February 25 to April 25 found that 64% of the public reported wearing a mask and gloves in crowded places.^81^ By May 9, the reported per-capita mortality in Iran was 78.4 per million, though many, even those within the Iranian government, have questioned the official figures.^82,83,S132^

In Jordan, a study conducted from March 19-21, 2020 found that 39.8% of university students wore a face mask when leaving home.^84^ King Abdullah recommended that the public wear masks when shopping on April 27.^S147^

In Yemen, 90% of women wear the niqab, which local doctors believe might reduce transmission of the virus by functioning as a mask.^85^ By May 9, the per-capita mortality in Yemen was 0.2 per million.

In Syria, a survey of university students conducted from March 19-21, 2020 found that 52% of respondents indicated that everyone should wear a mask when outside, but that 25% indicated that they did so at least sometimes, and 75% never wore a mask on the street.^86^

Government mandates or recommendations for mask wearing by the public were issued in Kuwait for gatherings by March 23,^14^ in Israel on April 1,^S135^ and in Bahrain on April 9.^S17^

### Masks in Africa

As noted above, 11 African countries recommended or mandated masks within 31 days of the onset of their outbreak: Sierra Leone, Mozambique, Malawi, São Tomé and Príncipe, Zambia, Chad, Benin, Sudan, Côte d’Ivoire, South Sudan, and Kenya (Table 2). In addition, the public widely sought masks to wear early in the outbreak in Gambia.^S100,S101^

In Ethiopia, 75.7% of chronic disease patients surveyed from March 2-April 10, 2020 agreed that it was important to wear a mask outside the home to prevent infection with coronavirus.^87^ A survey from March 20-24 found that 87% of the public believed wearing a mask could prevent spread of the virus, but only 14% had done so in the few days before the survey.^88^ Another survey from April 1-15 in southern Ethiopia found that 84% believed that wearing a mask was protective, 160 respondents (36%) had been to a crowded place in recent days, and 129 respondents (29%) had worn a mask when leaving home in recent days.^89^ Masks were mandated in public on April 11.^S95^ In a survey in that country from April 15-22, 84% believed a mask could provide protection from coronavirus, 137 people (40%) had gone to a crowded place after the onset of the pandemic, and 82 people (24%) had worn a mask outside the home.^90^ By May 9, Ethiopia had reported no deaths from coronavirus.

In Cameroon, the first cases of coronavirus were identified on March 6.^91^ From March 10-18, a study found that 93.5% of the public viewed the wearing of face masks as protective, and 21.7% had already purchased them.^91^ A study in Northern Cameroon conducted from March 1-28 found that only 13% wore a mask outside the home.^92^ A survey in Cameroon conducted from April 1 to 25 found that 83.6% reported wearing a mask at gatherings.^93^ On April 9, it was announced that masks would be mandatory in public beginning April 13.^66,S47,S48^ By May 9, the per-capita mortality was 4.1 per million.

In a city in the Democratic Republic of the Congo not yet affected by the pandemic at the time of a survey conducted from April 17 to May 11, 61% of respondents were aware of the value of wearing a face mask, 27% reported wearing a face mask since the pandemic began, and 65% felt that wearing a face mask was difficult.^94^

In Ghana, a study from March 27-29 of 43 public transport stations found that masks were worn by many people at one station, worn by a few people at 27 stations, and not worn at the remainder.^95^ On April 19, 2020, the president of Ghana announced that masks would be required in public.^S107,S108^

Masks were required in public in Nigeria on April 14.^96,S206^ A study in Nigeria from May 7 to 18 found that 65% of respondents had worn a mask outside the home in recent days.^97^

In South Africa from April 8-24, 2020, 85.6% of the public agreed that wearing a mask could help to prevent coronavirus infection.^S246^ South African health officials recommended mask wear in public on April 10.^S247^

In addition, government mandates or recommendations for mask wearing by the public were issued by April 16 in: Mauritius on March 31;^S178^ Tunisia^S274^ and Morocco^S187^ on April 6; Guinea on April 13;^S114^ Gabon on April 15;^63,66^ Equatorial Guinea on April 14;^S93^ and Libya on April 16.^S164^

### Masks in Europe

Most countries in Europe and North America failed to embrace masks early in their outbreaks, and only adopted mask policies after signs of health system overload became apparent. Only 3 countries in Europe appear to have had government recommendations for the public to wear masks within 31 days of the onset of their outbreak: Slovakia, Czechia, and Bosnia and Herzegovina (Table 2).

The first country in Europe to be strongly affected by the outbreak was Italy, which reported its first cases on January 31, among a family who arrived from China on January 23.^S136^ By March 10, doctors in Lombardy indicated that all intensive care beds were taken, and the system did not have enough respirators for the affected.^208X^ A poll found that only 26% of Italians wore a mask in public on March 11, but, with the rising health system overload, 59% did so on March 19^53^—at least 53 days from the local onset of the outbreak. Another poll confirms that the prevalence of mask wear exceeded 50% for the first time from March 19-21.^52^ Lombardy (April 5) and Tuscany (April 6) required the public to wear masks in early April.^S138^ A nationwide mandate to wear masks in shops and public transport was announced on April 28, to take effect on May 4.^S139^ Mask wear in public remained between 85% and 89% between April 16 and June 10.^53^ By May 9, the per-capita mortality in Italy was 502.7 per million.

The next country to suffer was Spain, which reported its first case on January 31,^S254^ and experienced its first death from the virus on February 13.^S255^ The prevalence of mask wear among the Spanish public was 5% on March 12, 25% on March 19, 42% on March 25, and 56% on April 8^53^—potentially 72 days after the entry of the virus into the country. Masks were mandated when in transit beginning April 11.^S256^ Mask wearing in public had climbed to 65% by April 16, 72% by April 30, and remained between 84% and 87% between May 20 and June 12.^53^ According to another survey, the prevalence of mask wear was 50% by March 21, 53% by April 4, and 61% by April 12.^52^ The per-capita mortality by May 9 was 566.3 per million.

In France, the first case of coronavirus was reported on January 24,^S97^ and the first death on February 14, of a man who arrived from China on January 16.^S98^ A poll found on March 10 that only 5% of those in France wore a mask in public.^53^ This number increased to 22% on March 27 and 25% on April 3,^53^ the day that the Académie Nationale de Médecine announced that masks should be compulsory in public^S99^—at least 72 days into their outbreak. Polls indicated that mask wear among the public climbed to 38% on April 10, 43% on April 17, 56% on May 1, 76% on May 20, and 75% on June 12.^53^ Mask wear below 50% in early April was confirmed in another survey.^52^ On May 7, it was announced that throughout France, including its overseas departments, masks would be mandatory on transport, starting May 11.^98^ By May 9, the per-capita mortality in France was 403.1 per million.

In Germany, the first case of COVID-19 was reported on January 27. The patient had contact with a colleague visiting from China beginning January 19.^S103^ By March 30, only 7% of the public reported wearing a mask in public.^53^ On March 31, the city of Jena mandated use of masks by the public.^S104^ The Robert Koch Institute recommended that the public wear masks on April 1^S105^—at least 70 days from the onset of the outbreak. Masks were worn by 14% of the public on April 6, 17% on April 13, 24% on April 20, 62%-64% from May 4 through June 18.^53^ Another survey confirms mask wear at or below 20% in March and early April.^52^ All German states had mandates relating to mask wear in public by April 22.^S106^ By May 9, the per-capita mortality was 90.1 per million.

In the United Kingdom, the first cases of coronavirus were reported on January 31.^S283^ Here, 2% of the population wore a mask by March 20, 11% by April 17, 20% on May 1, and 27% on June 17.^53^ Another survey confirms mask wear below 20% from March 12 to April 12.^52^ Masks were recommended in England on public transport and in shops on May 11^S284^—over 100 days after the local outbreak onset. On June 4, English authorities announced that masks would be mandatory on public transit, beginning June 15.^S285^ By May 9, the per-capita mortality was 465.3 per million.

In the Netherlands, from April 1 to 19, the prevalence of mask wear was approximately 7%.^99^ The prime minister announced on May 6, 2020 that beginning June 1, masks would be required on public transport due to their value in situations where social distancing was not possible.^S203^

In Belgium, from April 4 to 19, the prevalence of mask wear increased from about 30 to 37%.^99^ The Prime Minister of Belgium announced on April 24 that masks would be mandatory on public transport effective May 4.^S25^

In the Scandinavian countries of Sweden, Norway, Denmark, and Finland, polls repeatedly showed masks to be worn by 10% or less of the population from March 16 through June 9.^53^ This low usage occurred despite the fact that the government in Finland began recommending that the public wear masks on April 14.^S96^

In Switzerland, the chief of the Communicable Diseases Department recommended masks on public transport on June 15.^S264^ However, a survey released June 18 found that only 6% of Swiss public transport riders did so.^S264^

In Poland, the health minister announced on April 9 that a public mask mandate would go into effect on April 16, and mask vending machines began to be installed.^S216^ In Poland, from April 12-14, 2020, 60.4% of Polish students age 18 to 27 wore a face mask in the previous 7 days.^100^ By May 9, the per-capita mortality was 20.7 per million.

The first cases of coronavirus in Russia were reported on January 31, 2020.^S220^ In Russia, the prevalence of mask wear among the public was 11% by March 14, 19% by March 21, 36% by March 28, and 57% by April 4^52^—69 days after the estimated start of the outbreak. Mask wearing prevalence had increased to 59% by April 12.^52^ On May 11, it was announced that masks would be mandatory in shops and public transport (Time/Russia). By May 9, the per-capita mortality was 12.5 per million.

In Serbia, in April 2020, 60% of the public agreed they were willing to wear a mask during a pandemic, and respondents on average answered 3.25 (SD 1.6) on a 1 to 5 scale when asked if they wore masks, where 4 represented “agree” and 5 represented “strongly agree”.^101^

Some additional Western governments mandated or recommended mask-wearing in public by April 16, 2020. By March 29, masks were mandated in indoor public spaces in Slovenia.^S245^ In Austria, a mandate to wear masks in shops was announced on March 30, with the expectation that masks would be available by April 1.^S13^ In addition, the requirement to wear masks on public transit was announced there on April 6.^S14^ Masks were recommended for the public in Bulgaria^S41^ and Ukraine^S278,S279^ on March 30. In Lithuania, masks were recommended for the public on March 26,^S166^ and mandated on April 8.^S167^ Government mandates or recommendations for mask wearing by the public were also issued in: Turkey,^S275^ and Cyprus^S69^ on April 3; Estonia on April 5;^S94^ and Luxembourg on April 15.^S168^

### Masks in the United States and Canada

The earliest case of COVID-19 in the United States was a man who returned from China on January 15, 2020, and presented at an urgent care clinic on January 19.^102^ In the United States, the prevalence of mask wear in public was 7% on March 2, 5% on March 17, and 17% on March 30.^53^ The U.S. C.D.C. began recommending that asymptomatic people wear a mask in public on the evening of April 3^103^—at least 79 days after the virus had entered the country. Subsequently, the prevalence of mask wear was 29% on April 6, 49% on April 13, 58% on April 20, 63% on April 27, 68% on May 26, and 66% on June 8.^53^ Another survey found that the prevalence of mask wear was 32% from April 2-4, and 50% from April 9-12.^52^ According to another survey, from April 14-20, 36% of U.S. adults always wore a mask outside the home, 32% did so sometimes, and 31% never did.^S286^ Mask wearing varied by region. In Vermont, from May 16 to 30, 76% of people entering businesses were observed to wear a mask.^104^ On the other hand, in Wisconsin from June 3-9, only 42% of shoppers were observed to wear a mask.^105^ By May 9, the per-capita coronavirus-related mortality was 241.8 per million.

In Canada, the prevalence of mask wear was 6% on March 17, and 18% on April 6,^53^ when the government announced that masks were now recommended in public.^S49^ Uptake was gradual, with mask wearing at 16% on April 13, 31% on April 20, 41% on April 27, 49% on May 26, and 58% on June 11.^53^ Another survey confirms mask wear below 30% in March and early April.^52^ By May 9, the per-capita coronavirus-related mortality was 124.3 per million.

### Masks in Australia

In Australia, surveys of the public indicated that 10% wore a mask by March 15, which gradually increased to a high point of 27% by April 19, after which use gradually declined to 17% on June 5.^53^ Another survey confirms mask wear below 25% in March and early April.^52^

### Masks in Latin America and the Caribbean

On April 3, a reporter in Bogotá noted that 90% of the people on the street were wearing face masks.^S63^ On April 4, the government of Colombia mandated masks on public transport and shops.^S62-S65^

On April 6, the Minister of Health in Chile announced that masks would be mandatory on public transport starting April 8.^S57^ Due to the shortage of medical masks, the public was invited to make their own out of cloth.^S57^

Surveys indicate that in Mexico, the prevalence of public mask wear increased steadily from 17% on March 17 to 37% on April 6, 46% on April 13, 60% on April 20, and 67% on April 27.^53^ According to another survey, the prevalence was 31% by March 14, 36% by March 21, 46% by April 4, and 58% by April 9.^52^ Although some states had mandated masks, the federal minister leading the coronavirus response refrained from encouraging the public wearing of masks until May 5.^S180,S181^ By May 9, the per-capita mortality was 26.0 per million.

Ecuador did not require masks early in their outbreak. The first case of COVID-19 in Ecuador was reported on February 29 in a traveler who had arrived from Spain on February 14.^S83^ The first death was reported on March 13.^S84^ By April 3, it was noted in Guayaquil that mortuary facilities were overwhelmed, and bodies were being left on the streets.^S85^ On April 7, the Interior Minister of Ecuador announced that face masks were mandatory in public^S86^—at least 48 days (and possibly 53 days) after the local onset of the outbreak. By May 9, the reported mortality was 97.3 per million.

The first case of COVID-19 in Brazil was reported on February 26.^S37^ In Brazil, the prevalence of mask wear in public was 25% by March 14, 28% by March 21, 39% by April 4, and 56% by April 12^52^—50 days after the virus is estimated to have arrived in the country. By May 9, the per-capita mortality was 50.1 per million.

### Graphical Analysis of Mask Effect

Before the formal statistical analysis, we graphically illustrate the effect of mask wear (Figures 1, 2). The first figure demonstrates the effect of early mask usage (Figure 1). In the countries not using masks by April 16, or not using them until 60 days after the start of the outbreak, the per-capita mortality by May 9 rises dramatically if the infection has persisted in the country over 60 days (Figure 1, red line). On the other hand, countries in which a mask was used from 16 to 30 days after infection onset had per-capita mortality several orders of magnitude less by May 9 (Figure 1, orange line). When countries recommended masks within 15 days of the onset of the outbreak, the mortality was so low that the curve is difficult to distinguish from the x-axis (Figure 1, blue line).

**Figure 2.**
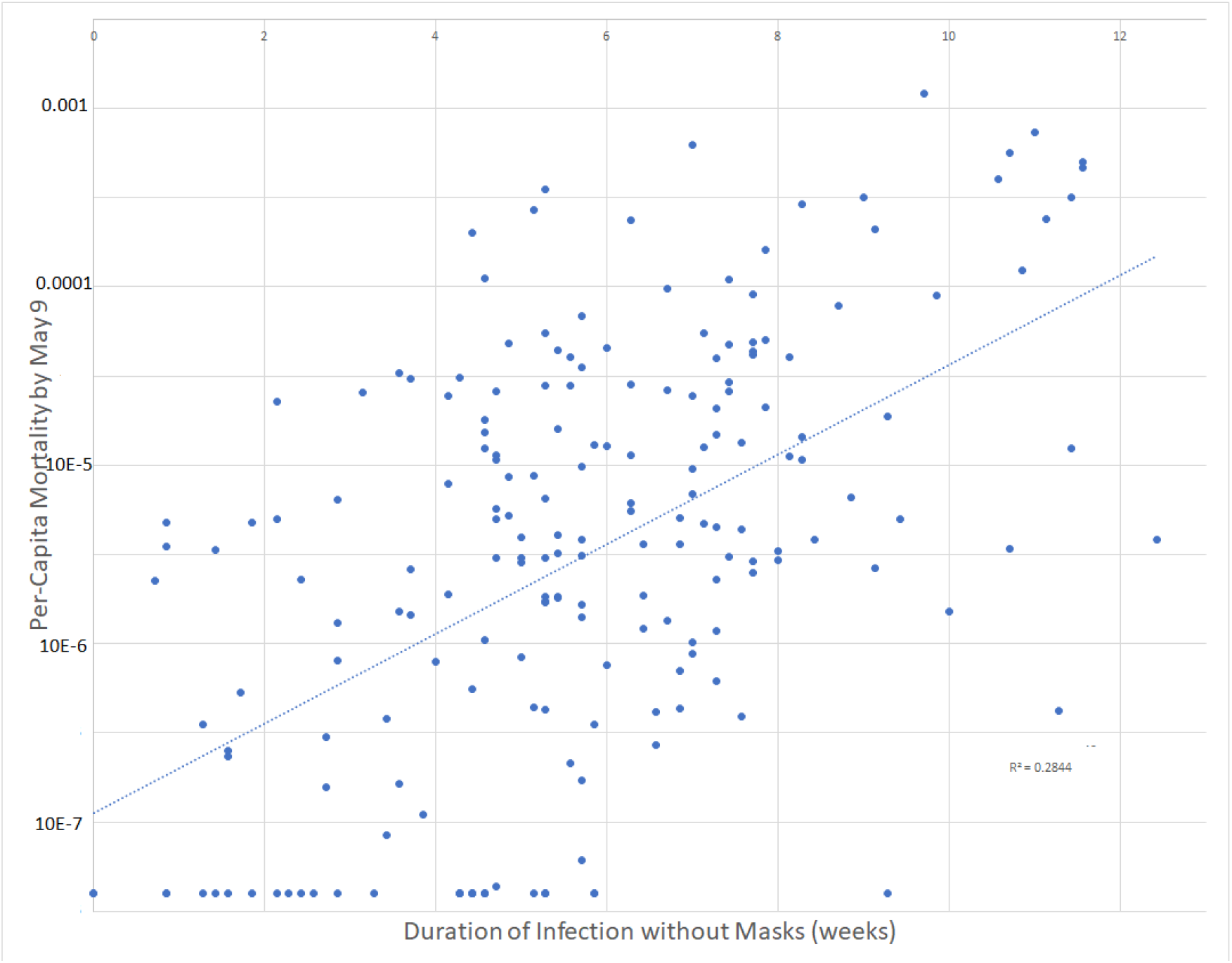
Scatter-plot of per-capita mortality by May 9, 2020 as a function of the period of the country’s outbreak without mask recommendations or norms. The dotted line represents the best fit using least-squares linear regression. Data for graph derived from 200 countries. Start of outbreak defined as 5 days before first case reported, or 23 days before the first death (whichever was earlier).

For instance, for the early mask-wearing countries in which the infection had arrived by January (Thailand, Japan, South Korea, Taiwan, Macau, Hong Kong, Vietnam, Cambodia, Malaysia, the Philippines), the virus was present in the country by 80 or more days by April 16 (Table 2). If masks had no effect, we might have expected these countries to have a mortality well over 200 deaths per million (Figure 1). Instead, the mortality for these 10 regions was 2.1 per million (SD 2.5, Table 2)—approximately a 100-fold reduction.

In order to provide some graphical idea of the scatter of the data when exponential growth is assumed, we graphed per-capita mortality by May 9 on a logarithmic scale as a function of the duration of the country’s outbreak not using masks in all 200 countries (Figure 2). This simple model explained 28.4% of the variation in per-capita mortality.

### Initial multivariable analyses

An initial multivariable analysis was conducted including all 200 countries. By multivariable linear regression, significant predictors of the logarithm of each country’s per-capita coronavirus mortality included: duration of infection in the country, duration of wearing masks (p<0.001), percentage of the population over age 60, and urbanization (all p≤0.009, Appendix Table A2). The association of mortality with the timing of international travel restrictions was of borderline statistical significance (p=0.051). The model explained 48.3% of the variation in per-capita mortality (Table A2).

We also prepared a multivariable model to predict the logarithm of per-capita coronavirus mortality in the 196 countries with obesity data. In this model, lockdown, obesity, temperature, and urbanization were retained due to their plausibility as important factors (Table 3). By multivariable linear regression, significant predictors of the logarithm of each country’s per-capita coronavirus mortality included: duration of infection in the country, duration of wearing masks, and percentage of the population over age 60 (all p<0.001, Table 3). The associations of obesity and or urbanization with increased mortality approached statistical significance (p=0.10, Table 3). When controlling for the duration of infection in the country, there appeared to be a negative association between mortality and time in lockdown (p=0.85) and time with international travel restrictions (p=0.07), though neither association reached statistical significance (Table 3). The model explained 51.0% of the variation in per-capita mortality.

**Table 3.**
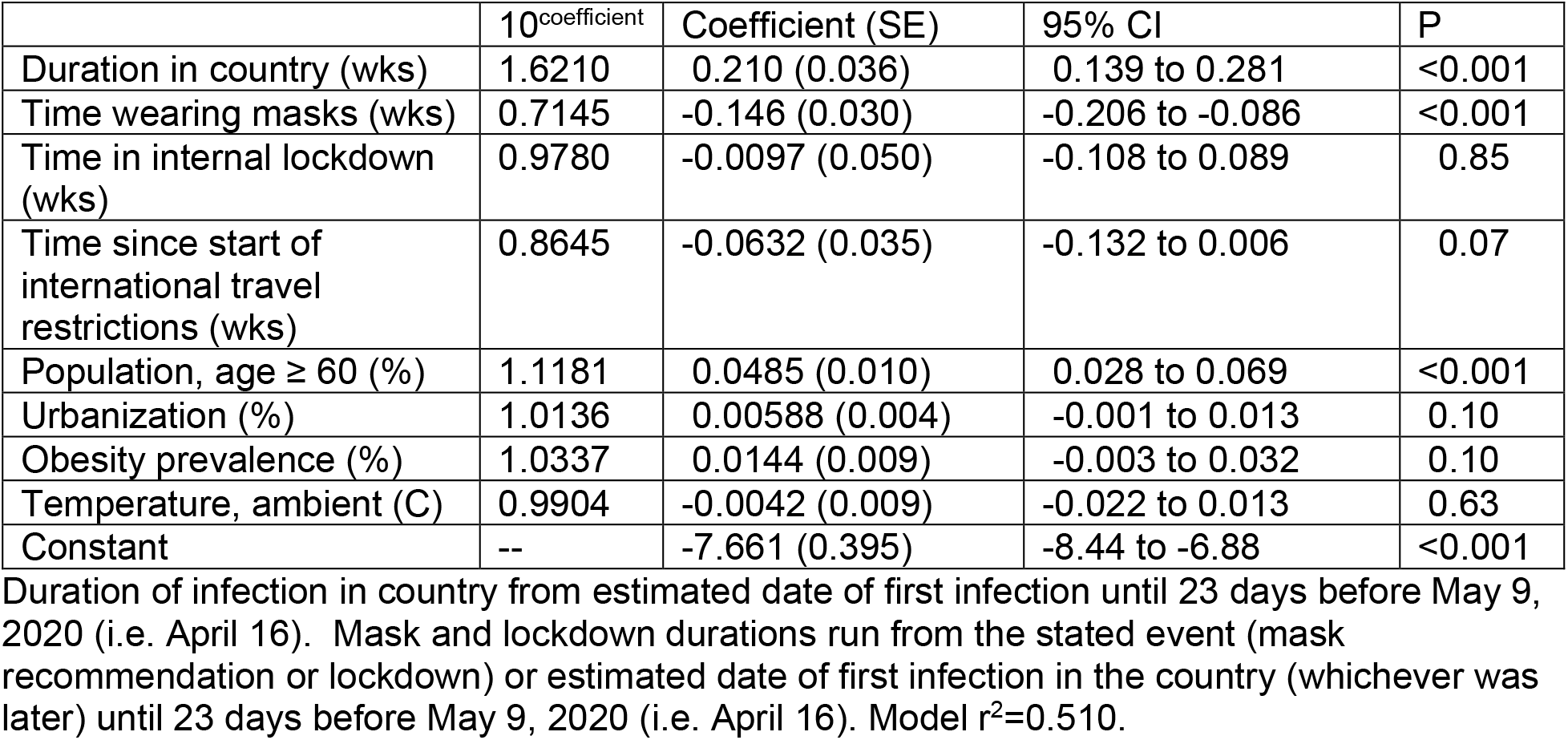
Predictors of (log) Country-wide Per-capita Coronavirus Mortality by May 9 by Multivariable Linear Regression in 196 Countries.

|  | 10<sup>coefficient</sup> | Coefficient (SE) | 95% CI | P |
| --- | --- | --- | --- | --- |
| Duration in country (wks) | 1.6210 | 0.210 (0.036) | 0.139 to 0.281 | <0.001 |
| Time wearing masks (wks) | 0.7145 | -0.146 (0.030) | -0.206 to -0.086 | <0.001 |
| Time in internal lockdown (wks) | 0.9780 | -0.0097 (0.050) | -0.108 to 0.089 | 0.85 |
| Time since start of international travel restrictions (wks) | 0.8645 | -0.0632 (0.035) | -0.132 to 0.006 | 0.07 |
| Population, age ≥ 60 (%) | 1.1181 | 0.0485 (0.010) | 0.028 to 0.069 | <0.001 |
| Urbanization (%) | 1.0136 | 0.00588 (0.004) | -0.001 to 0.013 | 0.10 |
| Obesity prevalence (%) | 1.0337 | 0.0144 (0.009) | -0.003 to 0.032 | 0.10 |
| Temperature, ambient (C) | 0.9904 | -0.0042 (0.009) | -0.022 to 0.013 | 0.63 |
| Constant | -- | -7.661 (0.395) | -8.44 to -6.88 | <0.001 |

Duration of infection in country from estimated date of first infection until 23 days before May 9, 2020 (i.e. April 16). Mask and lockdown durations run from the stated event (mask recommendation or lockdown) or estimated date of first infection in the country (whichever was later) until 23 days before May 9, 2020 (i.e. April 16). Model $r^2=0.510$ .

In countries not recommending masks, the per-capita mortality tended to increase each week by a factor of 1.621, or 62.1%. In contrast, in countries recommending masks, the per-capita mortality tended to increase each week by a factor of 1.6210*0.7145 = 1.158, or just 15.8%. With international travel restrictions in place (without masks), the per-capita mortality increased each week by (1.6210)(0.8645) = 1.401, or 40.1%. Under lockdown (without masks), the per-capita mortality increased each week by (1.6210)(0.9780) = 1.585, or 58.5%, i.e. slightly less than the baseline condition (Table 3).

A country with 10% more of its population living in an urban environment than another country tended to suffer a mortality 14.5% higher (10^0.0588^ = 1.145, Table 3). A country in which the percentage of the population age 60 or over is 10% higher than in another country tended to suffer mortality 206% higher (10^0.485^ = 3.06, Table 3). A country with a prevalence of obesity 10% higher tended to suffer mortality 39% higher (10^0.144^ = 1.39, Table 3).

### Numbers of Viral Tests

Among the 183 countries with viral (PCR) testing data by May 9, per-capita testing performed at all 3 time points was positively associated with per-capita mortality in univariate analysis (all p<0.001, Table 1). By May 9, 2020, low-mortality countries had performed 1 test for every 575 members of the population, while high-mortality countries had performed 1 test for every 81 members of the population (p<0.001, Table 1).

To the multivariable model (Table 3), we added testing by May 9, using data from 179 countries with both testing and obesity data. Duration of infection in the country, the duration that masks were recommended, and age at least 60 years continued to be significant predictors of per-capita mortality (all p≤0.001, Appendix Table A3). The model explained 52.5% of the variation in per-capita mortality. Each week the infection persisted in a country without masks was associated with a 62.7% increase in per-capita mortality (Table A3). In contrast, in countries where masks were recommended, the per-capita mortality tended to increase each week by 19.1% (because (1.6271)(0.7319) = 1.191, Table A3). In this model, the prevalence of obesity was associated with increased country-wide per-capita mortality, though the association was not significant (p=0.09). If the prevalence of obesity increased by 10% (e.g. from 10% to 20% of a population), the per-capita mortality tended to increase by 47% (Table A3)

In this model, a 10-fold increase (i.e. one logarithm) in per-capita testing tended to be associated with a 26.0% increase in reported per-capita mortality, though the trend was not close to reaching statistical significance (p=0.38, Appendix Table A3).

If early testing lowers mortality, one might expect negative regression coefficients. Testing on both April 16 and May 9 were added to the multivariable model of Table 3, using data from the 158 countries with both obesity and testing data by these dates. Per-capita testing (log) by April 16 was not negatively associated with per-capita mortality (log) by May 9 (coefficient 0.211, 95% CI -0.305 to 0.868, p=0.34).

Likewise, testing on both April 4 (the earliest archived data) and May 9 were added to the multivariable model of Table 3, using data from the 131 countries with both obesity and testing data by these dates. Per-capita testing (log) by April 4 was not significantly associated with per-capita mortality (log) by May 9 (coefficient -0.0535, 95% CI -0.380 to 0.273, p=0.75). Given the coefficient, a 10-fold (one log) increase in early testing would be associated with a (non-significant) decrease in per-capita mortality of 11.6%.

Only 5 countries had performed over 1 test for every 10 people in the country by May 9, 2020 (in order of most testing to least): the Faeroe Islands, Iceland, the Falkland Islands, the UAE, and Bahrain. The Faeroe and Falkland Islands reported no coronavirus-related deaths. The highest per-capita mortality among this group was 29.0 per million population (or 1 in 34,480 people), seen in Iceland.

### Containment and Testing Policies

For 169 countries, containment, testing, and health policies were scored by Oxford University.^9^ The following countries with mask policies by April 16 were included in this analysis, but not in the previous multivariable model, for lack of data on numbers of tests performed: China, Macau, Cameroon, Sierra Leone, and Sudan. In univariate analysis, scores for school closing, cancelling public events, international travel controls, and index of containment and health were significantly associated with lower per-capita mortality (all p<0.05, Table 4). Policies regarding workplace closing, restrictions on gatherings, closing public transport, stay at home requirements, internal movement restrictions, public information campaigns, testing, and contact tracing were not significant predictors of mortality (all p>0.05, Table 4). Likewise, overall indices of stringency and government response were not associated with mortality (all p>0.05, Table 4).

**Table 4.**
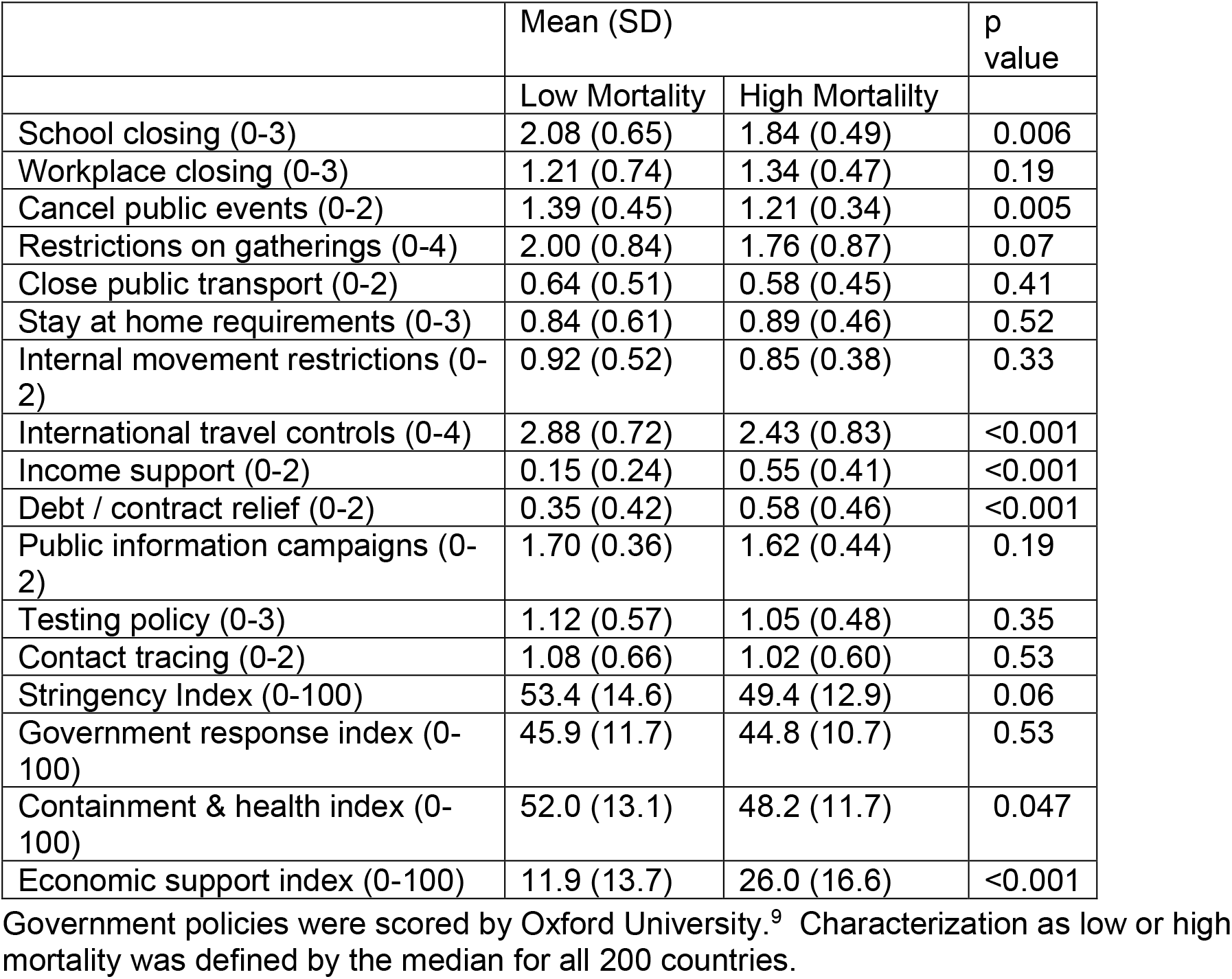
Government policies in 169 countries with low and high per-capita coronavirus mortality by May 9, 2020.

A multivariable model in 169 countries found that duration of the infection, duration masks were recommended, prevalence of age at least 60 years, obesity, and international travel restrictions were independently predictive of per-capita mortality (Table 5). The model explained 66.8% of the variation in per-capita mortality. At baseline, each week of the infection in a country without masks was associated with an increase in per-capita mortality of 50.9% (Table 5). In contrast, for each week that masks were worn, the per-capita mortality was associated with a lesser increase of 12.4% each week (given that 1.5085 (0.7449) = 1.124, Table 5).

**Table 5.** Predictors of (log) Country-wide Per-capita Coronavirus Mortality by May 9 by Multivariable Linear Regression in 169 Countries.

|  | 10<sup>coefficient</sup> | Coefficient (SE) | 95% CI | P |
| --- | --- | --- | --- | --- |
| Duration in country (wks) | 1.5085 | 0.1785 (0.031) | 0.118 to 0.239 | <0.001 |
| Time wearing masks (wks) | 0.7449 | -0.1279 (0.026) | -0.178 to -0.077 | <0.001 |
| Time in lockdown (wks) | 1.0195 | 0.0082 (0.044) | -0.076 to 0.093 | 0.85 |
| Time since start of international travel restrictions (wks) | 0.8291 | -0.0814 (0.029) | -0.140 to -0.023 | 0.006 |
| Population, age ≥ 60 (%) | 1.1725 | 0.0691 (0.009) | 0.051 to 0.087 | <0.001 |
| Urbanization (%) | 1.0149 | 0.0064 (0.003) | -0.0004 to 0.013 | 0.07 |
| Obesity prevalence (%) | 1.0459 | 0.0195 (0.008) | 0.003 to 0.036 | 0.02 |
| Temperature, ambient (C) | 1.0190 | 0.0082 (0.008) | -0.007 to 0.023 | 0.29 |
| Testing policy (0-3) | 1.0286 | 0.0122 (0.111) | -0.207 to 0.232 | 0.91 |
| Contact tracing (0-2) | 0.6737 | -0.172 (0.092) | -0.353 to 0.010 | 0.06 |
| Constant | -- | -7.885 (0.346) | -8.57 to -7.20 | <0.001 |

Duration of infection in country from estimated date of first infection until 23 days before May 9, 2020 (i.e. April 16). Mask and lockdown durations run from the stated event (mask recommendation or lockdown) or estimated date of first infection in the country (whichever was later) until 23 days before May 9, 2020 (i.e. April 16). Policies on testing, contact tracing, and international travel controls were scored by Oxford University. Model $r^2=0.668$ .

International travel restrictions were scored by Oxford as: (0) no measures, (1) screening, (2) quarantine arrivals from high-risk regions; and ban on arrivals from some (3) or all (4) regions. The international travel restrictions were scored as 4 in Greenland, 3.8 in Bermuda, 3.6 in Israel, 3.5 in Czechia and New Zealand, 3.1 in Taiwan, and 2.9 in Australia, and at the other extreme, were scored as 1.1 in Sweden, and as 0 in Iran, Luxembourg, and the UK.

International travel restrictions were associated with lower mortality, regardless of whether incorporated in the model as time since onset, or as mean score during the outbreak. We present the model based on the former because of the strength of the association, and for consistency with the models presented previously. The regression analysis suggested that for each week of travel restrictions (without masks), the per-capita mortality increased by 25.1% (given that 1.5085 (0.8291) = 1.251, Table 5).

Per-capita mortality was not significantly associated with policies regarding either testing policy (p=0.91), or contact tracing (p=0.06, Table 5). Testing policy was scored as: no policy (0), symptomatic with exposure, travel history, hospitalization, or key occupation (1), all symptomatic (2), or open to anyone (3). Testing policy tended to be positively associated with mortality. Contact tracing was scored as: none (0), some cases (1), or all cases (2), and tended to be inversely related with per-capita mortality (though not significantly). These countervailing associations meant that as compared with a country with no testing or tracing policy, a country which opened testing to the entire public with comprehensive contact tracing might be associated with a reported change in mortality of 10^(3*0.0122+2*(−0.172))^ = 0.493, i.e. a 51.7% reduction in per-capita mortality (though statistical significance was not demonstrated). Thus, testing and tracing may be important factors, but seem unlikely to account for the majority of the 100-fold variation in per-capita mortality between low and high mortality countries early in the course of the pandemic.

### Survey-modified Model

Surveys of mask wearing by the public during the exposure period were available for 41 countries (see above). To determine the influence that actual mask-wear, as opposed to mask policies, might have on the model, we scored countries as mask-wearing if at least 50% of the public wore a mask, and non-mask wearing if less than 50% of the population did so.

Based on surveys, Canada, Finland, France, Germany, and Malawi were not considered mask-wearing countries at any time during the exposure period (ending April 16). In contrast, Italy was scored as mask-wearing beginning March 19,^53^ Spain^53^ and India^52^ beginning March 21, Saudi Arabia beginning April 1,^53^ Russia beginning April 4, Singapore beginning April 10,^53^ and the United States, Brazil and Mexico beginning April 12.^52,53^

In this survey-modified model in 200 countries, duration of the outbreak, duration of mask wear, proportion of the population age 60 or over, and urbanization were all significant predictors of per-capita mortality (all p<0.01, Appendix Table A4). Time since the start of international travel restrictions tended to be inversely associated with mortality (p=0.051). Each week that the infection persisted in the country without masks was associated with a 59.9% increase in per-capita mortality. On the other hand, when masks were worn, the per-capita mortality only increased by 9.3% weekly, (1.5993)(0.6836) = 1.093, (Appendix Table A5). The model explained 48.3% of the variance in mortality.

## Discussion

These results confirm that in the first 4 months of 2020, there was marked variation between countries in mortality related to COVID-19. Countries in the lower half of mortality experienced an average COVID-19-related per-capita mortality of 0.99 deaths per million population, in contrast with an average of 93.3 deaths per million in the remaining countries. Depending on the model and dataset evaluated, statistically significant independent predictors of per-capita mortality included urbanization, fraction of the population age 60 years or over, prevalence of obesity, duration of the outbreak in the country, international travel restrictions, and the period of the outbreak subject to cultural norms or government policies favoring mask-wearing by the public.

These results support the universal wearing of masks by the public to suppress the spread of the coronavirus.^1^ Given the low levels of coronavirus mortality seen in the Asian countries which adopted widespread public mask usage early in the outbreak, it seems highly unlikely that masks are harmful.

On April 30, 2020, we originally published the finding that the logarithm of per-capita coronavirus mortality is linearly and positively associated with the duration of the outbreak without mask norms or mandates .^46^ This key finding was recently confirmed by Goldman Sachs chief economist Jan Hatzius, who cited our work.^107^ The regression analysis performed by Goldman Sachs confirms that, for prediction of both infection prevalence and mortality, the significance of the duration of mask mandates or norms in the model persists after controlling for age of the population, obesity, population density, and testing policy.^107^

One major limitation is that evidence concerning the actual prevalence of mask-wearing by the public is unavailable for most countries. Our survey of the literature is one of the more complete evaluations of the question to date. Available scholarship and surveys do corroborate reports in the news media that mask wear was common in public in many Asian countries, including Japan, the Philippines, Hong Kong, Vietnam, Malaysia, Taiwan, Thailand, China, Indonesia, India, Myanmar and Bangladesh (Table 2). Internet search data are consistent with interest in masks developing much earlier in the course of the pandemic in Asia than elsewhere.^108,109^ Mask wear was widespread in some low-mortality countries even before, or in the absence of, a formal government recommendation.

In addition, it is likely that the policies favoring mask-wearing in parts of the Middle East, Africa, Latin America and the Caribbean were markers of a general cultural acceptance of masks that helped to limit spread of the virus. Had there been adequate survey data to fully reflect the early wearing of masks in these regions, it is possible that the association of masks with lower mortality would be even stronger.

Conversely, in Western countries which had no tradition of mask-wearing, and which only recommended (rather than mandated) mask-wearing by the public, such as the United States, the practice has been steadily increasing, but change has not been immediate.

Much of the randomized controlled data on the effect of mask-wearing on the spread of respiratory viruses relates to influenza. One recent meta-analysis of 10 trials in families, students, or religious pilgrims found that the relative risk for influenza with the use of face masks was 0.78, a 22% reduction, though the findings were not statistically significant.^110^ Combining all the trials, there were 29 cases in groups assigned to wear masks, compared with 51 cases in control groups.^110^ The direct applicability of these results to mask-wearing at the population level is uncertain. For instance, there was some heterogeneity in methods of the component trials, with one trial assigning mask wearing to the person with a respiratory illness, another to his close contacts, and the remainder to both the ill and their contacts.^110^ Mask-wearing was inconsistent. The groups living together could not wear a mask when bathing, sleeping, eating, or brushing teeth.^111-113^ In one of the studies reviewed, parents wore a mask during the day, but not at night when sleeping next to their sick child.^113^ In a different trial, students were asked to wear a mask in their residence hall for at least 6 hours daily (rather than all the time).^111^ The bottom line is that it is nearly impossible for people to constantly maintain mask wear around the people with whom they live. In contrast, wearing a mask when on public transit or shopping is quite feasible. In addition, as an infection propagates through multiple generations in the population, the benefits multiply exponentially. Even if one accepts that masks would only reduce transmissions by 22%, then after 10 cycles of the infection, mask-wearing would reduce the level of infection in the population by 91.7%, as compared with a non-mask wearing population, at least during the period of exponential growth (because 0.78^10^ = 0.083). It is highly unlikely that entire countries or populations will ever be randomized to either wear, or not wear, masks. Public policies can only be formulated based on the best evidence available.

Some countries which used masks were better able to maintain or resume normal business and educational activities. For instance, in Taiwan, schools reopened on February 21, 2020, with parents directed to purchase 4 to 5 masks per week for each child.^S265^

Limits on international travel were significantly associated with lower per-capita mortality from coronavirus. On the other hand, nationwide policies to ban large gatherings and to close schools or businesses, tended to be associated with lower mortality, though not in a statistically significant fashion. However, businesses, schools, and individuals made decisions to limit contact, independent of any government policies. The adoption of numerous public health policies at the same time can make it difficult to tease out the relative importance of each.

Colder average monthly temperature was associated with higher levels of COVID-19 mortality in univariate analysis, but not when accounting for other independent variables. One reason that outdoor temperature might have limited association with the spread of the virus is that most viral transmission occurs indoors.^114^ We acknowledge that using the average temperature in the country’s largest city during the outbreak does not model the outbreak as precisely as modelling mortality and temperature separately in each of the thousands of cities around the world. However, to a first approximation, our method did serve to control for whether the country’s climate was tropical, temperate, or polar, and whether the outbreak began in late Winter (Northern hemisphere) or late Summer (Southern hemisphere). Environmental factors which could influence either human behavior or the stability and spread of virus particles are worthy of further study.

Presumably, high levels of testing might identify essentially all coronavirus-related deaths, and still higher levels of testing, combined with contact tracing, might lower mortality. Statistical support for the benefit of mass testing could not be demonstrated. It seems likely that countries which test at a low level are missing many cases. We identified just 5 countries (Iceland, the Faeroe Islands, the UAE, the Falkland Islands, and Bahrain) which had tested over one tenth of their population by May 9. All 5 countries had a mortality of 29 per million (1 in 34,480 people) or less. The degree to which these results would apply to larger, less isolated, or less wealthy countries is unknown. Statistical support for benefit of high levels of testing might be demonstrated if additional and more diverse countries are able to test at this level. The benefits of contact tracing policies with respect to mortality were of marginal statistical significance (p=0.06).

One limitation of our study is that the ultimate source of mortality data is often from governments which may not have the resources to provide a full accounting of their public health crises, or an interest in doing so. It should be noted that the benefit of wearing masks persisted in a model which excluded data from China (because no testing data were available, Appendix Table A3). We also acknowledge that country-wide analyses are subject to the ecologic fallacy.

The source for mortality and testing data we selected is publicly available,^7^ has been repeatedly archived,^11^ contains links to the source government reports for each country, and agrees with other coronavirus aggregator sites.^115^ In the interest of transparency, we presented the per-capita mortality data in Appendix Table A1. One might question whether any of these data sites or governments provide a complete and accurate picture of coronavirus mortality. But we must remember that this information does not exist in a vacuum. Independent sources confirm when mortality has been high. Social media alerted the world to the outbreaks in Wuhan, Iran, Italy, and New York. News reports have used aerial photography to confirm the digging of graves in Iran, New York, and Brazil. Long lines were seen to retrieve remains at crematoria in Wuhan. Mortuary facilities were inadequate to meet the demand in New York, and Guayaquil.^S85^ Conversely, signs of health system overload have been noted to be absent in the countries reporting low mortality. The health systems in Hong Kong, Taiwan, Japan, and South Korea are believed to be transparent. Reporters in Vietnam have even called hospitals and funeral homes to confirm the absence of unusual levels of activity.^S297^ Therefore, while no data source is perfect, we believe that the data used in the paper are consistent with observations from nongovernmental sources, and are comparable in reliability to those in other scholarly works.

It is not the case that countries which reported no deaths due to coronavirus simply were not exposed to the virus. All 200 countries analyzed did report COVID-19 cases. Several countries which traditionally use masks and sustained low mortality (or none) are close to and have strong travel links to China. Some of these countries reported cases early in the global pandemic (Table 2). Community transmission has been described in Vietnam.^116^

The pandemic is a matter of universal concern, but ophthalmologists have specific reasons to understand and prevent infection with SARS-CoV-2. The virus can cause a conjunctivitis, and has been identified in tears.^117,118^ It is possible that transmission can occur by conjunctival exposure to droplets.^117^ Ophthalmology was among the specialties whose residents were at higher risk of coronavirus infection.^119^ COVID-19 claimed the lives of 3 ophthalmologists from Wuhan Central Hospital, including 33-year-old Li Wenliang, who was admonished for sharing news of the novel pneumonia online.^117,S61^ As of April 15, 2020, at least 8 ophthalmologists had died from COVID.^120^

In summary, older age of the population, urbanization, obesity, and longer duration of the outbreak in a country were independently associated with higher country-wide per-capita coronavirus mortality. International travel restrictions were associated with lower per-capita mortality. However, other containment measures, testing and tracing polices, and the amount of viral testing were not statistically significant predictors of country-wide coronavirus mortality, after controlling for other variables. In contrast, societal norms and government policies supporting mask-wearing by the public were independently associated with lower per-capita mortality from COVID-19. The use of masks in public is an important and readily modifiable public health measure.

## Data Availability

Data can be requested from the first author.

## Acknowledgements / Disclosures. Funding

None.

## Author contributions

Design of the study (CL, EI); collection of data (CL, EI, JL, MH); management and analysis of data (CL, EI); interpretation of the data (CL, EI, JL, CM, AG); preparation of the manuscript (CL,EI,JL); review and approval of the manuscript (CL, EI, JL, MH, CM, AG).

## Appendix

### Supplemental Tables

**Table A1.** **Per-capita COVID-19 Mortality by May 9 and Date of Mask Recommendation or Widespread Use Based on Cultural Norms**.

| Country. | COVID-19 Mortality (per M. pop.) by May 9. | Date Masks Recommended or Widely Used by Cultural Norms. |
| --- | --- | --- |
| Afghanistan | 3.0 |  |
| Albania | 10.8 |  |
| Algeria | 11.3 | 5/18/2020 |
| Andorra | 621.2 | 4/17/2020 |
| Angola | 0.1 | 4/23/2020 |
| Antigua & Barbuda | 30.6 | 4/5/2020 |
| Argentina | 6.6 | 4/18/2020 |
| Armenia | 14.8 | 5/14/2020 |
| Aruba | 28.1 |  |
| Australia | 3.8 |  |
| Austria | 68.3 | 3/30/2020 |
| Azerbaijan | 3.1 | 5/1/2020 |
| Bahamas | 28.0 | 4/19/2020 |
| Bahrain | 4.7 | 4/9/2020 |
| Bangladesh | 1.3 | 3/19/2020 |
| Barbados | 24.4 | 4/11/2020 |
| Belarus | 13.3 |  |
| Belgium | 740.4 | 4/24/2020 |
| Belize | 5.0 | 5/1/2020 |
| Benin | 0.2 | 4/6/2020 |
| Bermuda | 112.4 |  |
| Bhutan | 0.0 | 3/11/2020 |
| Bolivia | 9.8 |  |
| Bosnia & Herzegov. | 31.1 | 3/29/2020 |
| Botswana | 0.4 | 5/1/2020 |
| Brazil | 50.1 |  |
| British Virgin Is. | 33.1 | 4/28/2020 |
| Brunei | 2.3 | 3/22/2020 |
| Bulgaria | 13.0 | 3/30/2020 |
| Burkina Faso | 2.3 | 4/20/2020 |
| Burundi | 0.1 |  |
| Cabo Verde | 3.6 | 5/5/2020 |
| Cambodia | 0.0 | 1/28/2020 |
| Cameroon | 4.1 | 4/9/2020 |
| Canada | 124.3 | 4/6/2020 |
| Carib. Netherlands | 0.0 |  |
| Cayman Is. | 15.2 | 3/30/2020 |
| Central Afric. Rep. | 0.0 |  |
| Chad | 1.9 | 4/13/2020 |
| Channel Is. | 235.8 |  |
| Chile | 15.9 | 4/6/2020 |
| China | 3.2 | 1/20/2020 |
| Colombia | 8.7 | 4/4/2020 |
| Congo (Brazzaville) | 1.8 | 4/30/2020 |
| Costa Rica | 1.2 |  |
| Croatia | 21.2 | 4/24/2020 |
| Cuba | 6.5 | 4/2/2020 |
| Curacao | 6.1 |  |
| Cyprus | 12.4 | 4/3/2020 |
| Czechia | 25.8 | 3/19/2020 |
| Dem. Rep. Congo | 0.4 |  |
| Denmark | 90.8 |  |
| Djibouti | 3.0 | 5/10/2020 |
| Dominica | 0.0 | 4/9/2020 |
| Dominican Republic | 35.5 | 4/6/2020 |
| Ecuador | 97.3 | 4/7/2020 |
| Egypt | 5.0 | 5/31/2020 |
| El Salvador | 2.6 | 4/4/2020 |
| Equatorial Guinea | 2.9 | 4/14/2020 |
| Eritrea | 0.0 |  |
| Estonia | 45.2 | 4/5/2020 |
| Eswatini | 1.7 |  |
| Ethiopia | 0.0 | 4/11/2020 |
| Faeroe Islands | 0.0 |  |
| Falkland Islands | 0.0 |  |
| Fiji | 0.0 |  |
| Finland | 47.8 | 4/14/2020 |
| France | 403.1 | 4/3/2020 |
| French Polynesia | 0.0 |  |
| Gabon | 3.6 | 4/15/2020 |
| Gambia | 0.4 |  |
| Georgia | 2.5 | 4/17/2020 |
| Germany | 90.1 | 4/1/2020 |
| Ghana | 0.7 | 4/19/2020 |
| Gibraltar | 0.0 |  |
| Greece | 14.5 | 4/27/2020 |
| Greenland | 0.0 |  |
| Grenada | 0.0 | 4/3/2020 |
| Guatemala | 1.3 | 4/9/2020 |
| Guinea | 0.8 | 4/13/2020 |
| Guinea-Bissau | 1.5 | 5/11/2020 |
| Guyana | 12.7 | 4/9/2020 |
| Haiti | 1.1 | 5/4/2020 |
| Honduras | 10.8 | 4/6/2020 |
| Hong Kong | 0.5 | 1/24/2020 |
| Hungary | 41.9 | 4/29/2020 |
| Iceland | 29.3 |  |
| India | 1.5 | 4/4/2020 |
| Indonesia | 3.5 | 2/24/2020 |
| Iran | 78.4 | 3/29/2020 |
| Iraq | 2.7 | 4/19/2020 |
| Ireland | 292.8 | 5/15/2020 |
| Isle of Man | 270.5 |  |
| Israel | 28.5 | 4/1/2020 |
| Italy | 502.7 | 4/28/2020 |
| Ivory Coast | 0.8 | 4/4/2020 |
| Jamaica | 3.0 | 4/8/2020 |
| Japan | 4.8 | 1/16/2020 |
| Jordan | 0.9 | 4/27/2020 |
| Kazakhstan | 1.7 | 5/26/2020 |
| Kenya | 0.6 | 4/4/2020 |
| Kuwait | 11.5 | 3/23/2020 |
| Kyrgyzstan | 1.8 | 5/10/2020 |
| Laos | 0.0 | 3/6/2020 |
| Latvia | 9.5 | 4/27/2020 |
| Lebanon | 3.8 | 4/25/2020 |
| Liberia | 4.0 | 4/24/2020 |
| Libya | 0.4 | 4/16/2020 |
| Liechtenstein | 26.2 | 5/15/2020 |
| Lithuania | 18.0 | 3/26/2020 |
| Luxembourg | 161.3 | 4/15/2020 |
| Macao | 0.0 | 1/23/2020 |
| Madagascar | 0.0 | 4/20/2020 |
| Malawi | 0.2 | 4/4/2020 |
| Malaysia | 3.3 | 1/30/2020 |
| Maldives | 5.5 | 5/19/2020 |
| Mali | 1.8 | 5/10/2020 |
| Malta | 11.3 | 5/1/2020 |
| Mauritania | 0.2 | 5/6/2020 |
| Mauritius | 7.9 | 3/31/2020 |
| Mayotte | 40.3 | 5/11/2020 |
| Mexico | 26.0 | 5/5/2020 |
| Moldova | 39.9 | 5/7/2020 |
| Mongolia | 0.0 | 1/31/2020 |
| Montenegro | 12.7 | 4/30/2020 |
| Montserrat | 200.3 | 4/29/2020 |
| Morocco | 5.0 | 4/6/2020 |
| Mozambique | 0.0 | 4/4/2020 |
| Myanmar | 0.1 | 4/5/2020 |
| Namibia | 0.0 | 5/2/2020 |
| Nepal | 0.0 | 3/25/2020 |
| Netherlands | 316.4 | 5/6/2020 |
| New Caledonia | 0.0 | 4/30/2020 |
| New Zealand | 4.4 |  |
| Nicaragua | 0.8 |  |
| Niger | 1.9 | 5/12/2020 |
| Nigeria | 0.6 | 4/14/2020 |
| North Macedonia | 43.7 | 4/23/2020 |
| Norway | 40.4 |  |
| Oman | 3.3 | 5/18/2020 |
| Pakistan | 2.9 | 5/31/2020 |
| Palestine | 0.4 | 5/5/2020 |
| Panama | 54.9 | 4/7/2020 |
| Papua New Guinea | 0.0 | 4/24/2020 |
| Paraguay | 1.4 | 4/7/2020 |
| Peru | 55.0 | 4/3/2020 |
| Philippines | 6.4 | 1/30/2020 |
| Poland | 20.7 | 4/10/2020 |
| Portugal | 110.4 | 4/27/2020 |
| Qatar | 4.5 | 4/22/2020 |
| Réunion | 0.0 | 5/7/2020 |
| Romania | 48.8 | 4/22/2020 |
| Russia | 12.5 | 5/11/2020 |
| Rwanda | 0.0 | 4/18/2020 |
| Saint Kitts & Nevis | 0.0 | 4/2/2020 |
| Saint Lucia | 0.0 | 4/7/2020 |
| San Marino | 1208.3 | 4/17/2020 |
| São Tomé & Príncipe | 22.8 | 4/22/2020 |
| Saudi Arabia | 6.9 | 4/28/2020 |
| Senegal | 1.0 | 4/17/2020 |
| Serbia | 24.4 | 4/29/2020 |
| Seychelles | 0.0 | 6/9/2020 |
| Sierra Leone | 2.3 | 4/1/2020 |
| Singapore | 3.4 | 4/3/2020 |
| Sint Maarten | 349.8 | 4/18/2020 |
| Slovakia | 4.8 | 3/15/2020 |
| Slovenia | 48.6 | 3/29/2020 |
| Somalia | 3.0 |  |
| South Africa | 3.1 | 4/10/2020 |
| South Korea | 5.0 | 1/30/2020 |
| South Sudan | 0.0 | 4/29/2020 |
| Spain | 566.3 | 4/11/2020 |
| Sri Lanka | 0.4 | 4/11/2020 |
| St. Vincent & Gren. | 0.0 | 4/26/2020 |
| Sudan | 1.5 | 3/16/2020 |
| Suriname | 1.7 | 5/31/2020 |
| Sweden | 318.8 |  |
| Switzerland | 211.4 | 6/15/2020 |
| Syria | 0.2 |  |
| Taiwan | 0.3 | 1/27/2020 |
| Tanzania | 0.4 |  |
| Thailand | 0.8 | 1/28/2020 |
| Timor-Leste | 0.0 | 3/28/2020 |
| Togo | 1.2 | 4/19/2020 |
| Trinidad & Tobago | 5.7 | 4/5/2020 |
| Tunisia | 3.8 | 4/7/2020 |
| Turkey | 44.3 | 4/3/2020 |
| Turks and Caicos | 25.8 | 4/30/2020 |
| Uganda | 0.0 | 5/1/2020 |
| Ukraine | 8.6 | 3/30/2020 |
| United Arab Emir. | 18.7 | 3/27/2020 |
| United Kingdom | 465.3 | 5/11/2020 |
| United States | 241.8 | 4/3/2020 |
| Uruguay | 5.2 | 4/10/2020 |
| Uzbekistan | 0.3 | 3/25/2020 |
| Venezuela | 0.4 | 3/13/2020 |
| Vietnam | 0.0 | 1/27/2020 |
| Yemen | 0.2 |  |
| Zambia | 0.4 | 4/4/2020 |
| Zimbabwe | 0.3 | 5/1/2020 |

**Table A2.** Predictors of (log) Country-wide Per-capita Coronavirus Mortality by May 9 by Multivariable Linear Regression in 200 Countries.

| | $10^{\text{coefficient}}$ | Coefficient (SE) | 95% CI | P |
| --- | --- | --- | --- | --- |
| Duration in country (weeks) | 1.5993 | 0.2039 (0.037) | 0.131 to 0.277 | <0.001 |
| Time wearing masks (weeks) | 0.6836 | -0.1652 (0.030) | -0.224 to -0.106 | <0.001 |
| Time since international travel restrictions (weeks) | 0.8529 | -0.0691 (0.035) | -0.139 to 0.0004 | 0.051 |
| Time in internal lockdown (weeks) | 1.0210 | 0.0090 (0.051) | -0.092 to 0.110 | 0.86 |
| Population, age≥60 (%) | 1.1367 | 0.0556 (0.010) | 0.035 to 0.076 | <0.001 |
| Urbanization (%) | 1.0185 | 0.00796 (0.003) | 0.002 to 0.014 | 0.009 |
| Temperature (C) | 0.9988 | -0.00052 (0.009) | -0.018 to 0.017 | 0.95 |
| Constant | -- | -7.66 (0.393) | -8.43 to -6.88 | <0.001 |

Duration of infection in country from estimated date of first infection until 23 days before May 9, 2020 (i.e. April 16). Mask and lockdown durations run from the stated event (mask recommendation or lockdown) or estimated date of first infection in the country (whichever was later) until 23 days before May 9, 2020 (i.e. April 16). Model $r^2=0.483$ .

**Table A3.** Predictors of (log) Country-wide Per-capita Coronavirus Mortality by May 9 by Multivariable Linear Regression in 179 Countries.

|  | 10<sup>coefficient</sup> | Coefficient (SE) | 95% CI | P |
| --- | --- | --- | --- | --- |
| Duration in country (weeks) | 1.6271 | 0.211 (0.038) | 0.136 to 0.287 | <0.001 |
| Time wearing masks (weeks) | 0.7319 | -0.136 (0.032) | -0.199 to -0.072 | <0.001 |
| Time in lockdown (weeks) | 0.9877 | -0.0054 (0.054) | -0.113 to 0.102 | 0.92 |
| International travel controls (time since start, weeks) | 0.8686 | -0.0612 (0.038) | -0.135 to 0.013 | 0.11 |
| Population, % age 60 or over | 1.0909 | 0.0378 (0.012) | 0.015 to 0.061 | 0.001 |
| Urbanization (%) | 1.0132 | 0.00568 (0.004) | -0.002 to 0.013 | 0.14 |
| Obesity prevalence (%) | 1.0395 | 0.0168 (0.010) | -0.003 to 0.036 | 0.09 |
| Temperature (C) | 0.9824 | -0.0077 (0.009) | -0.026 to 0.010 | 0.40 |
| Testing (log per cap., by May 9) | 1.2604 | 0.101 (0.115) | -0.127 to 0.328 | 0.38 |
| Constant | -- | -7.309 (0.621) | -8.54 to -6.08 | <0.001 |

Based on 179 countries with both obesity and testing data by May 9. Duration of infection in country from estimated date of first infection until 23 days before May 9, 2020 (i.e. April 16). Mask and lockdown durations run from the stated event (mask recommendation or lockdown) or estimated date of first infection in the country (whichever was later) until 23 days before May 9, 2020 (i.e. April 16). Model $r^2=0.525$ .

**Table A4.** Predictors of (log) Country-wide Per-capita Coronavirus Mortality by May 9 by Multivariable Linear Regression in 200 Countries, with Mask Wear Determined by Recommendations and Surveys (When Available).

|  | 10coefficient | Coefficient (SE) | 95% CI | P |
| --- | --- | --- | --- | --- |
| Duration in country (weeks) | 1.5993 | 0.204 (0.037) | 0.131 to 0.277 | <0.001 |
| Time wearing masks (weeks) | 0.6836 | -0.165 (0.030) | -0.224 to -0.106 | <0.001 |
| Time in lockdown (weeks) | 0.9021 | 0.0090 (0.051) | -0.092 to 0.110 | 0.86 |
| Time since start of international travel controls (weeks) | 0.8529 | -0.0691 (0.035) | -0.139 to 0.0004 | 0.051 |
| Population, age≥60 (%) | 1.1367 | 0.0556 (0.010) | 0.035 to 0.076 | <0.001 |
| Urbanization (%) | 1.0185 | 0.00796 (0.003) | 0.002 to 0.014 | 0.009 |
| Temperature | 0.9988 | -0.00052 (0.009) | -0.018 to 0.017 | 0.95 |
| Constant | -- | -7.658 (0.393) | -8.434 to -6.882 | <0.001 |

` Duration of infection in country from estimated date of first infection until 23 days before May 9, 2020 (i.e. April 16). Mask and lockdown durations run from the stated event (mask recommendation or lockdown) or estimated date of first infection in the country (whichever was later) until 23 days before May 9, 2020 (i.e. April 16). Model $r^2=0.483$ .

## References

1. Leffler CT, Ing E, McKeown CA, Pratt D, Grzybowski A. Final Country-wide Mortality from the Novel Coronavirus (COVID-19) Pandemic and Notes Regarding Mask Usage by the Public. April 4, 2020. Available from: https://www.researchgate.net/publication/340438732_Country-wide_Mortality_from_the_Novel_Coronavirus_COVID-19_Pandemic_and_Notes_Regarding_Mask_Usage_by_the_Public Accessed: May 20, 2020.

2. Leffler CT, Hogan MC. Age-dependence of mortality from novel coronavirus disease (COVID-19) in highly exposed populations: New York transit workers and residents and Diamond Princess passengers. MedRxiv. Available from: https://doi.org/10.1101/2020.05.14.20094847 Accessed May 20, 2020.

3. Jordan RE, Adab P, Cheng KK. Covid-19: risk factors for severe disease and death. BMJ. 2020;368:m1198. Available from: https://www.bmj.com/content/368/bmj.m1198.long Accessed May 20, 2020.

4. Zhu Y, Xie J. Association between ambient temperature and COVID-19 infection in 122 cities from China. Science of The Total Environment. 2020 Mar 30:138201. Available from: https://www.ncbi.nlm.nih.gov/pmc/articles/PMC7142675/ Accessed May 20, 2020.

5. Squalli J. Evaluating the determinants of COVID-19 mortality: A cross-country study. medRxiv. 2020 Jan 1. Available from: https://www.medrxiv.org/content/10.1101/2020.05.12.20099093v1 Accessed May 20, 2020.

6. Syed Q, Sopwith W, Regan M, Bellis MA. Behind the mask. Journey through an epidemic: some observations of contrasting public health responses to SARS. Journal of Epidemiology & Community Health. 2003 Nov 1;57(11):855–6.

7. Worldometers. COVID-19 Coronavirus Pandemic. Available from: https://www.worldometers.info/coronavirus/?utm_campaign=homeAdUOA?Si Accessed July 4, 2020.

8. European Centre for Disease Prevention and Control. Download today’s data on the geographic distribution of COVID-19 cases worldwide. Available from: https://www.ecdc.europa.eu/en/publications-data/download-todays-data-geographic-distribution-covid-19-cases-worldwide Accessed April 16, 2020.

9. Oxford Coronavirus Government Response Tracker. University of Oxford. Available from: https://www.bsg.ox.ac.uk/research/research-projects/coronavirus-government-response-tracker Accessed July 4, 2020.

10. Hale T, Angrist N, Kira B, Phillips T, Webster S. Variation in government responses to COVID-19. BSG-WP-2020/032. Version 6.0. University of Oxford. Blavatnik School of Government. May 2020 Available from: https://www.bsg.ox.ac.uk/sites/default/files/2020-05/BSG-WP-2020-032-v6.0.pdf Accessed May 31, 2020.

11. Worldometers. COVID-19 Coronavirus Pandemic. April 5, 2020. Available from: https://web.archive.org/web/20200405020252/ https://www.worldometers.info/coronavirus/ Accessed May 17, 2020.

12. No author listed. World Climate Guide. Available from: https://www.climatestotravel.com/ Accessed May 14, 2020.

13. No author listed. List of Cities by Average Temperature. Available from: https://en.wikipedia.org/wiki/List_of_cities_by_average_temperature Accessed May 14, 2020.

14. Laestadius MP, Wang Y, Taleb ZB, Kalan ME, Cho Y, Manganello J. Online National Health Agency Mask Guidance for the Public in Light of COVID-19: Content Analysis. JMIR Public Health Surveillance. 2020; 6(2):e19501. Available from: https://publichealth.jmir.org/2020/2/e19501/PDF Accessed June 7, 2020.

15. Howard J. What Countries Require Masks in Public or Recommend Masks? #Masks4all Available from: https://masks4all.co/what-countries-require-masks-in-public/ Accessed May 30, 2020.

16. Countries in the world by population (2020). Available from: https://www.worldometers.info/world-population/population-by-country/ Accessed May 16, 2020.

17. United Nations Population Division, New York. World Population Prospects: The 2019 Revision. Population, surface area, and density. Available from: http://data.un.org/_Docs/SYB/PDFs/SYB61_T02_Population,%20Surface%20Area%20and%20Density.pdf Accessed May 16, 2020.

18. International Monetary Fund. World Economic Outlook Database, October 2019. Report for Selected Countries and Subjects. Available from: https://www.imf.org/en/Publications/SPROLLs/world-economic-outlook-databases#sort=%40imfdate%20descending Accessed April 11, 2020.

19. World Bank. Urban population (% of total population). United Nations Population Division. World Urbanization Prospects: 2018 Revision. Available from: https://data.worldbank.org/indicator/SP.URB.TOTL.in.zs Accessed May 16, 2020.

20. The American Cancer Society. The Tobacco Atlas. 2020 Available from: https://tobaccoatlas.org/topic/prevalence/ Accessed April 11, 2020.

21. WHO Report on the Global Tobacco Epidemic, 2009. Surveys of adult tobacco use in WHO Member States. Available from: https://www.who.int/tobacco/mpower/2009/Appendix_VIII-table_1.pdf Accessed May 16, 2020.

22. World Bank. Smoking prevalence, males (% of adults). Available from: https://data.worldbank.org/indicator/SH.PRV.SMOK.MA?locations=ME Accessed May 16, 2020.

23. World Bank. Smoking prevalence, females (% of adults). Available from: https://data.worldbank.org/indicator/SH.PRV.SMOK.FE?locations=ME Accessed May 16, 2020.

24. Central Intelligence Agency. The World Factbook. Country Comparison. Obesity. Adult Prevalence Rate. Available from: https://www.cia.gov/library/publications/the-world-factbook/rankorder/2228rank.html Accessed May 29, 2020.

25. Parris D. The Alarming Increase of Diabetes in Bermuda. West Indian Med J. 2014; 63(7): 685–686. Available from: https://www.ncbi.nlm.nih.gov/pmc/articles/PMC4668975/ Accessed May 29, 2020.

26. Fu FH. A comparison of lifestyle management practices of residents of three Chinese cities: Hong Kong, Macau and Weihai (Shandong). International Journal of Sports. 2013;3:115–26.

27. Ramachandran A, Snehalatha C. Rising burden of obesity in Asia. Journal of obesity. 2010 Aug 30;2010.

28. Chobanyan N, Allison Kruger K, Nebb S, Jackson G, Asin V. Evaluation of Environmental Risk Factors for Type 2 Diabetes in Sint Maarten. J Environ Anal Toxicol. 2016;6(386):2161–0525.

29. Bjerregaard P, Jørgensen ME, Greenland Population Study Group. Prevalence of obesity among Inuit in Greenland and temporal trend by social position. American Journal of Human Biology. 2013 May;25(3):335–40.

30. Veyhe AS, Andreassen J, Halling J, Grandjean P, Petersen MS, Weihe P. Prevalence of type 2 diabetes and prediabetes in the Faroe Islands. Diabetes research and clinical practice. 2018 Jun 1;140:162–73.

31. Abdeen Z, Jildeh C, Dkeideek S, Qasrawi R, Ghannam I, Al Sabbah H. Overweight and obesity among Palestinian adults: analyses of the anthropometric data from the first national health and nutrition survey (1999-2000). Journal of Obesity. 2012 Feb 21;2012.

32. Grol ME, Eimers JM, Alberts JF, Bouter LM, Gerstenbluth I, Halabi Y, Van Sonderen E, Van den Heuvel WJ. Alarmingly high prevalence of obesity in Curacao: data from an interview survey stratified for socioeconomic status. International journal of obesity. 1997 Nov;21(11):1002–9.

33. Corsenac P, Annesi-Maesano I, Hoy D, Roth A, Rouchon B, Capart I, Taylor R. Overweight and obesity in New Caledonian adults: Results from measured and adjusted self-reported anthropometric data. Diabetes research and Clinical Practice. 2017 Nov 1;133:193–203.

34. Favier F, Jaussent I, Le Moullec N, Debussche X, Boyer MC, Schwager JC, Papoz L, REDIA Study Group. Prevalence of Type 2 diabetes and central adiposity in La Reunion Island, the REDIA Study. Diabetes research and clinical practice. 2005 Mar 1;67(3):234–42.

35. Grievink L, Alberts JF, O’niel J, Gerstenbluth I. Waist circumference as a measurement of obesity in the Netherlands Antilles; associations with hypertension and diabetes mellitus. European journal of clinical nutrition. 2004 Aug;58(8):1159–65.

36. Solet JL, Baroux N, Pochet M, Benoit-Cattin T, De Montera AM, Sissoko D, Favier F, Fagot-Campagna A. Prevalence of type 2 diabetes and other cardiovascular risk factors in Mayotte in 2008: the MAYDIA study. Diabetes & metabolism. 2011 Jun 1;37(3):201–7.

37. Daigre JL, Atallah A, Boissin JL, Jean-Baptiste G, Kangambega P, Chevalier H, Balkau B, Smadja D, Inamo J. The prevalence of overweight and obesity, and distribution of waist circumference, in adults and children in the French Overseas Territories: the PODIUM survey. Diabetes & metabolism. 2012 Nov 1;38(5):404–11.

38. Guariguata L, Brown C, Sobers N, Hambleton I, Samuels TA, Unwin N. An updated systematic review and meta-analysis on the social determinants of diabetes and related risk factors in the Caribbean. Revista Panamericana de Salud Pública. 2018;42: e171.

39. World Health Organization. Nutrition, Physical Activity and Obesity San Marino. Available from: http://www.euro.who.int/__data/assets/pdf_file/0016/243322/San-Marino-WHO-Country-Profile.pdf?ua=1 Accessed May 29, 2020.

40. States of Jersey. Health Profile for Jersey 2014…with comparisons of Guernsey, English regions and Europe. Available from: https://www.gov.je/SiteCollectionDocuments/Government%20and%20administration/R%20Health%20Profile%20Jersey%202014%2020140410%20MM%20v1.pdf Accessed May 29, 2020.

41. Isle of Man Government. Island’s plan to tackle childhood obesity launched. January 22, 2013. Available from: https://www.gov.im/news/2013/jan/22/islands-plan-to-tackle-childhood-obesity-launched/ Accessed May 29, 2020.

42. Gibraltar Health Authority. Health and Lifestyle. Survey Report of the Adult Population of Gibraltar. 2015. Available from: https://www.gha.gi/wp-content/uploads/2016/12/GHA_Lifestyle_Report2015LR.pdf Accessed May 29, 2020.

43. James J, Soyibo AK, Hurlock L, Gordon-Strachan G, Barton EN. Cardiovascular Risk Factors in an Eastern Caribbean Island: Prevalence of Non-Communicable Chronic Diseases and Associated Lifestyle Risk Factors for Cardiovascular Morbidity and Mortality in the British Virgin Islands. West Indian Med J 2012; 61(4):429–36.

44. Lauer SA, Grantz KH, Bi Q, Jones FK, Zheng Q, Meredith HR, Azman AS, Reich NG, Lessler J. The incubation period of coronavirus disease 2019 (COVID-19) from publicly reported confirmed cases: estimation and application. Annals of Internal Medicine. Mar 10, 2020.

45. Verity R, Okell LC, Dorigatti I, Winskill P, Whittaker C, Imai N, Cuomo-Dannenburg G, Thompson H, Walker P, Fu H, Dighe A. Estimates of the severity of COVID-19 disease. Lancet Infectious Disease. 2020: 1–9.

46. Leffler CT, Ing E, Lykins JD, McKeown CA, Grzybowski A. Prevention of the spread of coronavirus using masks. Annals of Internal Medicine. April 30, 2020. Available from: https://www.acpjournals.org/doi/10.7326/M20-1342 Accessed May 20, 2020.

47. European Centre for Disease Prevention and Control. COVID-19 Coronavirus data. Available from: https://data.europa.eu/euodp/en/data/dataset/covid-19-coronavirus-data Accessed April 16, 2020.

48. Feng S, Shen C, Xia N, Song W, Fan M, Cowling BJ. Rational use of face masks in the COVID-19 pandemic. Lancet Respir Med. 2020 Mar 20. pii: S2213-2600(20)30134-X. doi: 10.1016/S2213-2600(20)30134-X.

49. Mandavilli A. W.H.O. Finally Endorses Masks to Prevent Coronavirus Transmission. New York Times. June 5, 2020. Available from: https://www.nytimes.com/2020/06/05/health/coronavirus-masks-who.html Accessed June 13, 2020.

50. Erkhembayar R, Dickinson E, Badarch D, Narula I, Thomas GN, Ochir C, Manaseki-Holland S. Early policy actions and emergency response to the COVID-19 pandemic in Mongolia: experiences and challenges. Lancet Global Health. July 23, 2020. Available from: https://doi.org/10.1016/S2214-109X(20)30295-3 Accessed July 31, 2020.

51. Kamata K, Ohmagari N, Tokuda Y. Universal public use of surgical mask and respiratory viral infection. J Gen Fam Med 2020; 21: 35–6.

52. Bhatia M. 3 in 4 Indians wearing masks to protect themselves from COVID-19 – Ipsos 15-Nation Survey. Highest surge in mask adoption seen for Vietnam, China, Italy, Japan and India. April 21, 2020. Avaliable from: https://www.ipsos.com/en-in/3-4-indians-wearing-masks-protect-themselves-covid-19-ipsos-15-nation-survey Accessed June 21, 2020.

53. Bhatia P. YouGov COVID-19 behavior changes tracker: wearing a face mask when in public places. Available from: https://in.yougov.com/en-hi/news/2020/04/20/yougovs-international-covid-19-tracker-reveals-cha/ Accessed June 19, 2020.

54. Tam VC, Tam SY, Poon WK, Law HK, Lee SW. A reality check on the use of face masks during the COVID-19 outbreak in Hong Kong. Lancet. 2020 May 1;22.

55. Nguyen NP, Hoang TD, Tran VT, Vu CT, Siewe Fodjo JN, Colebunders R, Dunne MP, Vo VT. Preventive behavior of Vietnamese people in response to the COVID-19 pandemic. medRxiv. 2020. Available from: https://www.medrxiv.org/content/10.1101/2020.05.14.20102418v1 Accessed June 15, 2020.

56. Chiu WT, Laporte RP, Wu J. Determinants of Taiwan’s Early Containment of COVID-19 Incidence. AJPH. 2020; 110(7): 943–4.

57. Chiang CH, Chiang CH, Chiang CH. Maintaining mask stockpiles in the COVID-19 pandemic: Taiwan as a learning model. Infection Control & Hospital Epidemiology 2020; 1–2.

58. Zeng N, Li Z, Ng S, Chen D, Zhou H. Epidemiology reveals mask wearing by the public is crucial for COVID-19 control. Medicine in Microecology May 13, 2020. Available from: https://doi.org/10.1016/j.medmic.2020.100015 Accessed June 19, 2020.

59. Han G, Zhou YH. Possibly critical role of wearing masks in general population in controlling COVID-19. Journal of Medical Virology. 2020; 1–3. Available from: https://onlinelibrary.wiley.com/doi/pdf/10.1002/jmv.25886 Accessed June 19, 2020.

60. Ardan M, Rahman FF, Geroda GB. The influence of physical distance to student anxiety on COVID-19, Indonesia. Journal of Critical Reviews. 2020;7(17):1126–32.

61. Doung-ngern P, Suphanchaimat R, Panjagampatthana A, Janekrongtham C, Ruampoom D, Daochaeng N, Eungkanit N, Pisitpayat N, Srisong N, Yasopa O, Plernprom P. Associations between wearing masks, washing hands, and social distancing practices, and risk of COVID-19 infection in public: a cohort-based case-control study in Thailand. medRxiv. Accessed June 19, 2020.

62. Banda J, Dube A, Brumfield S, Amoah A, Crampin A, Reniers G, Helleringer S. Knowledge and behaviors related to the COVID-19 pandemic in Malawi. medRxiv. 2020 Jan 1.

63. U.S. Embassy in Gabon. COVID-19 Information for Gabon and São Tomé and Príncipe. April 23, 2020. Available from: https://web.archive.org/web/20200426105023/ https://ga.usembassy.gov/u-s-citizen-services/coronavirus-update/ Accessed May 30, 2020.

64. Wadood A, Mamun AS, Rafi A, Islam K, Mohd S, Lee LL, Hossain G. Knowledge, attitude, practice and perception regarding COVID-19 among students in Bangladesh: Survey in Rajshahi University. medRxiv (2020).

65. Ferdousa MZ, Islama MS, Sikdera MT, Md AS. Knowledge, attitude, and practice regarding COVID-19 outbreak in Bangladesh: An online-based cross-sectional study.

66. Herman T, Maarek E, Wilde N, Adao F, Abousaada S. COVID-19: Initial responses of certain African countries. Africa Notes. April 30, 2020. Available from: https://www.lexology.com/library/detail.aspx?g=9ddb5508-c7eb-4519-b2d5-1eeae40268c8 Accessed July 7, 2020.

67. Mousa KN, Saad MM, Abdelghafor MT. Knowledge, attitudes, and practices surrounding COVID-19 among Sudan citizens during the pandemic: an online cross-sectional study. Sudan Journal of Medical Sciences (SJMS). 2020; 15:32–45.

68. Mya Kyaw S, Aye SM, Hlaing Win A, Hlaing Su S, Thida A. Awareness, perceived risk and protective behaviours of Myanmar adults on COVID-19. International Journal of Community Medicine and Public Health. 2020; 7:1627–36.

69. Huang C, Wang Y, Li X, Ren L, Zhao J, Hu Y, Zhang L, Fan G, Xu J, Gu X, Cheng Z. Clinical features of patients infected with 2019 novel coronavirus in Wuhan, China. Lancet. 2020;395(10223):497–506.

70. Adhikari SP, et al. Epidemiology, causes, clinical manifestation and diagnosis, prevention and control of coronavirus disease (COVID-19) during the early outbreak period: a scoping review. Infectious Diseases of Poverty 2020;9.1: 1–12.

71. Sharma V, Ortiz MR, Sharma N. Risk and Protective Factors for Adolescent and Young Adult Mental Health Within the Context of COVID-19: A Perspective From Nepal. J Adolesc Health. May 20, 2020. Available from: https://www.ncbi.nlm.nih.gov/pmc/articles/PMC7237905/ Accessed June 10, 2020.

72. Singh DR, Sunuwar DR, Karki K, Ghimire S, Shrestha N. Knowledge and Perception Towards Universal Safety Precautions During Early Phase of the COVID-19 Outbreak in Nepal. Journal of Community Health. May 13, 2020. Available from: https://link.springer.com/content/pdf/10.1007/s10900-020-00839-3.pdf Accessed June 10, 2020.

73. Alam K, Palaian S, Shankar PR, Jha N. General public’s knowledge and practices on face mask use during the COVID-19 pandemic: a cross-sectional exploratory survey from Dharan, Nepal. Available from: https://assets.researchsquare.com/files/rs-42148/v1/6800c8ad-e059-4fe9-a56a-3587646bb2fc.pdf Accessed August 1, 2020.

74. Gudi SK, Undela K, Venkataraman R, Mateti UV, Chhabra M, Nyamagoud S, Tiwari KK. Knowledge and Beliefs towards Universal Safety Precautions to flatten the curve during Novel Coronavirus Disease (nCOVID-19) Pandemic among general Public in India: Explorations from a National Perspective. Available from: https://www.medrxiv.org/content/10.1101/2020.03.31.20047126v1 Accessed July 31, 2020.

75. Mousavi SH, Abdi M, Zahid SU, Wardak K. Coronavirus disease 2019 (COVID-19) outbreak in Afghanistan: Measures and challenges. Infect Control Hosp Epidemiol. 2020 May 15 : 1–2.

76. Malik S. Knowledge of COVID-19 Symptoms and Prevention among Pakistani Adults: A Cross-sectional Descriptive Study. PsyArXiv Available from: https://psyarxiv.com/wakmz Accessed June 28, 2020.

77. Mirza TM, Ali R, Musarrat Khan HM. The knowledge and perception of COVID-19 and its preventive measures, in public of Pakistan. Pak Armed Forces Med J 2020; 70 (2): 338–45.

78. Alkhamees AA, Alrashed SA, Alzunaydi AA, Almohimeed AS, Aljohani MS. The psychological impact of COVID-19 pandemic on the general population of Saudi Arabia. Comprehensive Psychiatry. 2020; 102. Available from: https://doi.org/10.1016/j.comppsych.2020.152192 Accessed July 31, 2020.

79. Begum F. Knowledge, Attitudes, and Practices towards COVID-19 among B. Sc. Nursing Students in Selected Nursing Institution in Saudi Arabia during COVID-19 Outbreak: An Online Survey. Saudi Journal of Nursing and Health Care. 2020;3(7):194–8.

80. Samir Abdelhafiz A, Mohammed Z, Ibrahim ME, Ziady HH, Alorabi M, Ayyad M, Sultan EA. Knowledge, perceptions, and attitudes of Egyptians towards the novel coronavirus disease (COVID-19). Journal of Community Health. April 21, 2020. Available from: https://link.springer.com/content/pdf/10.1007/s10900-020-00827-7.pdf Accessed June 19, 2020.

81. Kakemam E, Ghoddoosi-Nejad D, Chegini Z, Momeni K, Salehinia H, Hassanipour S, Ameri H, Arab-Zozani M. Knowledge, attitudes, and practices among the general population during COVID-19 outbreak in Iran: A national cross-sectional survey. medRxiv. 2020.

82. Tuite AR, Bogoch II, Sherbo R, Watts A, Fisman D, Khan K. Estimation of coronavirus disease 2019 (COVID-19) burden and potential for international dissemination of infection from Iran. Annals of Internal Medicine. 2020 May 19;172(10):699–701.

83. Zhuang Z, Zhao S, Lin Q, Cao P, Lou Y, Yang L, He D. Preliminary estimation of the novel coronavirus disease (COVID-19) cases in Iran: A modelling analysis based on overseas cases and air travel data. International Journal of Infectious Diseases. 2020 May 1;94:29–31.

84. Olaimat AN, Aolymat I, Elsahoryi N, Shahbaz HM, Holley RA. Attitudes, Anxiety, and Behavioral Practices Regarding COVID-19 among University Students in Jordan: A Cross-Sectional Study. Am J Trop Med Hygiene. 2020 July 8, 2020. Available from: http://www.ajtmh.org/docserver/fulltext/10.4269/ajtmh.20-0418/tpmd200418.pdf?expires=1596199449&id=id&accname=guest&checksum=37D8DD443CB1475F07A83B5F6BEA76DC Accessed July 31, 2020.

85. Kadi HO. Yemen is free of COVID-19. International Journal of Clinical Virology. 2020: 32–3. Available from: http://www.yemenuniversity.com/ar/content/uploads/2020/05/ijcv-aid1012.pdf Accessed June 30, 2020.

86. Alhamid A, Aljarad Z, Alhamid A. Knowledge and behaviors towards COVID-19 among University of Aleppo students: an online cross-sectional survey. Available from: https://www.medrxiv.org/content/10.1101/2020.07.11.20151035v1 Accessed July 31, 2020.

87. Akalu Y, Ayelign B, Molla MD. Knowledge, Attitude and Practice Towards COVID-19 Among Chronic Disease Patients at Addis Zemen Hospital, Northwest Ethiopia. Infection and Drug Resistance. 2020 Jun 24;13:1949–60.

88. Kebede Y, Yitayih Y, Birhanu Z, Mekonen S, Ambelu A. Knowledge, perceptions and preventive practices towards COVID-19 among Jimma University Medical Center visitors, Southwest Ethiopia. Researchsquare. doi: 10.21203/rs.3.rs-25865/v1

89. Kassaw C. The Psychological Impact of COVID-19 Pandemic among Communities Living in Dilla Town, Ethiopia, April 2020. June 30, 2020. Available from: https://www.preprints.org/manuscript/202006.0356/v1 Accessed July 31, 2020.

90. Bekele D, Tolossa T, Tsegaye R, Teshome W. The knowledge and practice towards COVID-19 pandemic prevention among residents of Ethiopia. An online cross-sectional study. BioRxiv. 2020 Jan 1.

91. Nicholas T, Mandaah FV, Esemu SN, Vanessa AB, Gilchrist KT, Vanessa LF, Shey ND. COVID-19 knowledge, attitudes and practices in a conflict affected area of the South West Region of Cameroon. May-Aug 2020; 36 https://www.panafrican-med-journal.com/content/series/35/2/34/full/ Accessed June 28, 2020.

92. Davy AA, Victor AD, Valery NN. Socio-Eco-nomic Household Surveys on the Application of Basic Preven-tive Measures Dictated by the WHO to Stop the Spread of COVID-19 in the Northern Zone of Cameroon. Res Rev Infect Dis. 2020;3(1):44–8.

93. Akwa TE, Muthini MJ, Ning TR. Assessing the Perceptions and Awareness of COVID-19 (Coronavirus) in Cameroon. European Journal of Medical and Educational Technologies. Apr 25, 2020. Available from: https://papers.ssrn.com/sol3/papers.cfm?abstract_id=3628380 Accessed June 29, 2020.

94. Mbiya BM, Djeugoue SL, Kanda EL, Mbuyi DK, Malundu TB, Mushiya RC, Disashi GT. Coronavirus-19 in the Democratic Republic of Congo: Public Views, Attitudes, and Beliefs in an Unaffected Area: The Case of the City of Mbujimayi. Available from: https://www.preprints.org/manuscript/202006.0317/v1 Accessed June 29, 2020.

95. Bonful H, Addo-Lartey A, Aheto J, Sarfo B, Aryeetey R. Limiting Spread of COVID-19 in Ghana: Compliance audit of selected transportation stations in the Greater Accra region of Ghana. medRxiv. 2020.

96. Ogoina D. COVID-19: The Need for Rational Use of Face Masks in Nigeria. American Society of Tropical Medicine and Hygiene. May 15, 2020. Available from: https://www.ajtmh.org/content/journals/10.4269/ajtmh.20-0433 Accessed June 10, 2020. Also, personal communication, Dimie Ogoina, June 8, 2020.

97. Isah MB, Abdulsalam M, Bello A, Ibrahim MI, Usman A, Nasir A, Abdulkadir B, Usman AR, Matazu KI, Sani A, Shuaibu A. Corona Virus Disease 2019 (COVID-19): Knowledge, attitudes, practices (KAP) and misconceptions in the general population of Katsina State, Nigeria. medRxiv. 2020.

98. No author listed. Réouverture des plages, quatorzaine Outre-mer, port du masque: Édouard Philippe détaille le plan de deconfinement. Franceinfo. May 7, 2020. Available from: https://la1ere.francetvinfo.fr/coronavirus-edouard-philippe-detaille-le-plan-de-deconfinement-a-quatre-jours-du-11-mai-830390.html Accessed July 11, 2020.

99. Perrotta D, Grow A, Rampazzo F, Cimentada J, Del Fava E, Gil-Clavel S, Zagheni E. Behaviors and attitudes in response to the COVID-19 pandemic: Insights from a cross-national Facebook survey. medRxiv. 2020.

100. Matusiak Ł, Szepietowska M, Krajewski P, BiaŁynicki-Birula R, Szepietowski JC. Inconveniences due to the use of face masks during the COVID-19 pandemic: a survey study of 876 young people. Dermatologic Therapy. May 14, 2020. Available from: https://onlinelibrary.wiley.com/doi/pdf/10.1111/dth.13567?casa_token=XcjP5hoHNVQAAAAA:6W_DYS_Ks5ObyZffz2abviHXOoyahPPIMfW9C40ONQu0aVwr91Divct2fysDS48DYTldkdaDVgWxS7_x Accessed June 19, 2020.

101. Cvetković VM, Nikolić N, Radovanović Nenadić U, Öcal A, K Noji E, Zečević M. Preparedness and Preventive Behaviors for a Pandemic Disaster Caused by COVID-19 in Serbia. International Journal of Environmental Research and Public Health. 2020 Jun; 17(11):4124.

102. Holshue ML, DeBolt C, Lindquist S, Lofy KH, Wiesman J, Bruce H, Spitters C, Ericson K, Wilkerson S, Tural A, Diaz G. First case of 2019 novel coronavirus in the United States. New England Journal of Medicine. 2020; 382: 929–936.

103. United States Centers for Disease Control. Recommendation Regarding the Use of Cloth Face Coverings, Especially in Areas of Significant Community-Based Transmission. Available from: https://www.cdc.gov/coronavirus/2019-ncov/preventgetting-sick/cloth-face-cover.html Accessed April 3, 2020.

104. Beckage B, Buckley T, Beckage ME. Prevalence of mask wearing in northern Vermont in response to SARS-CoV-2. medRxiv. 2020.

105. Haischer MH, Beilfuss R, Hart MR, Opielinski L, Wrucke D, Zirgaitis G, Uhrich TD, Hunter SK. Who is wearing a mask? Gender-, age-, and location-related differences during the COVID-19 pandemic. medRxiv. 2020.

106. Naraynsingh V, Harnanan D, Maharaj R, Naraynsingh R. COVID-19 in the West Indies: Trinidad and Tobago Experience. Journal of Lumbini Medical College. 2020; 8(1): 1–2. Available from: https://www.jlmc.edu.np/index.php/JLMC/article/view/347 Accessed June 28, 2020.

107. Hatzius J, Struyven D, Rosenberg I. Face Masks and GDP. Goldman Sachs Research. June 29, 2020. Available from: https://www.goldmansachs.com/insights/pages/face-masks-and-gdp.html Accessed July 2, 2020.

108. Chin CY, Wang CL. Effectiveness of COVID-19 Pandemic Prevention: A cross-country Comparison of Digital Footprint of Google Search Data. Advances in Management and Applied Economics. 2020 Jul 1;10(4):23–34.

109. Wong SH, Teoh JY, Leung CH, Wu WK, Yip BH, Wong MC, Hui DS. Covid-19 and public interest in face mask use. American Journal of Respiratory and Critical Care Medicine. 2020 Jun 15.

110. Xiao J, Shiu EY, Gao H, Wong JY, Fong MW, Ryu S, Cowling BJ. Nonpharmaceutical Measures for Pandemic Influenza in Nonhealthcare Settings-Personal Protective and Environmental Measures. Emerging Infectious Diseases. 2020; 26(5): 967–75.

111. Aiello AE, Perez V, Coulborn RM, Davis BM, Uddin M, Monto AS. Facemasks, hand hygiene, and influenza among young adults: a randomized intervention trial. PLoS One. 2012;7:e29744.

112. Cowling BJ, Fung RO, Cheng CK, Fang VJ, Chan KH, Seto WH, et al. Preliminary findings of a randomized trial of non-pharmaceutical interventions to prevent influenza transmission in households. PLoS One. 2008;3:e2101.

113. MacIntyre CR, Cauchemez S, Dwyer DE, Seale H, Cheung P, Browne G, et al. Face mask use and control of respiratory virus transmission in households. Emerg Infect Dis. 2009;15:233–41.

114. Qian H, Miao T, Liu L, Zheng X, Luo D, Li Y. Indoor transmission of SARS-CoV-2. medRxiv 2020.04.04.20053058; doi: https://doi.org/10.1101/2020.04.04.20053058 Accessed June 13, 2020.

115. COVID-19 Dashboard by the Center for Systems Science and Engineering (CSSE) at Johns Hopkins University (JHU). Available from: https://coronavirus.jhu.edu/map.html Accessed June 10, 2020.

116. Pham TQ, Rabaa M, Duong LH, Dang TQ, Dai Tran Q, Quach HL, Hoang NA, Phung DC, Ngu ND, Tran AT, La NQ. The first 100 days of SARS-CoV-2 control in Vietnam. medRxiv. 2020.

117. Kuo IC, O’Brien TP. COVID-19 and ophthalmology: an underappreciated occupational hazard. Infect Control Hosp Epidemiol. 2020 May 15: 1–2. Available from: https://www.ncbi.nlm.nih.gov/pmc/articles/PMC7256213/ Accessed July 31, 2020.

118. Xia J, Tong J, Liu M, Shen Y, Guo D. Evaluation of coronavirus in tears and conjunctival secretions of patients with SARS-CoV-2 infection. Journal of Medical Virology. 2020 Jun;92(6):589–94.

119. Breazzano MP, Shen J, Abdelhakim AH, Glass LR, Horowitz JD, Xie SX, de Moraes CG, Chen-Plotkin A, Chen RW. Resident physician exposure to novel coronavirus (2019-nCoV, SARS-CoV-2) within New York City during exponential phase of COVID-19 pandemic: Report of the New York City Residency Program Directors COVID-19 Research Group. medRxiv. 2020 Jan 1.

118. Ing EB, Xu QA, Salimi A, Torun N. Physician deaths from corona virus (COVID-19) disease. Occupational Medicine. 2020; 70(5): 370–4. Available from: https://doi.org/10.1093/occmed/kqaa088 Accessed August 1, 2020.

## Supplemental References by Country

S1. Smith SS. Service Delivery in Taliban-Influenced Areas of Afghanistan. United States Institute of Peace. April 2020. Available from: https://www.usip.org/sites/default/files/2020-04/20200430-sr_465-_service_delivery_in_taliban_influenced_areas_of_afghanistan-sr.pdf Accessed June 28, 2020.

S2. No author listed. COVID-19: Algeria imposes wearing of masks. Africa News. May 19, 2020. Available from: https://www.africanews.com/2020/05/19/covid-19-algeria-imposes-wearing-of-masks/ Accessed August 1, 2020.

S3. Ignacio P. En Andorra ya se permite hacer ejercicio: mascarilla obligatoria y el ciclismo sigue prohibido. Brujulabike. April 18, 2020. Available from: https://www.brujulabike.com/andorra-permite-ejercicio-mascarilla-obligatoria-prohibido-ciclismo/ Accessed July 6, 2020.

S4. No author listed. Angola: COVID-19 - Angola Detects Seven New Positive Cases. All Africa. May 9, 2020. Available from: https://allafrica.com/stories/202005110170.html Accessed July 6, 2020.

S5. No author listed. Angola reports first two confirmed COVID-19 cases. Xinhua. March 21, 2020. Available from: http://www.china.org.cn/world/Off_the_Wire/2020-03/21/content_75842926.htm Accessed July 7, 2020.

S6. No author listed. Covid-19 faz as primeiras duas vitimas em Angola. Platina Line. March 29, 2020. Available from: https://web.archive.org/web/20200330145319/ http://platinaline.com/covid-19-as-primeiras-duas-vitimas-angola/ Accessed July 7, 2020.

S7. No author listed. Covid-19: Use of a mask becomes mandatory. Agencia Angola Press. April 16, 2020. Available from:http://www.angop.ao/angola/en_us/noticias/saude/2020/3/16/Covid-Use-mask-becomes-mandatory,a665b68b-834b-4758-b3c5-b856a7854911.html Accessed July 7, 2020.

S8. No author listed. Angola: Authorities extend state of emergency to May 10. Garda. April 24, 2020.https://www.garda.com/crisis24/news-alerts/336006/angola-authorities-extend-state-of-emergency-to-may-10-update-6 Accessed July 7, 2020.

S9. De Shong D. Antigua and Barbuda records first case of the novel coronavirus. Loop News Barbados. March 13, 2020. Available from:http://www.loopnewsbarbados.com/content/antigua-and-barbuda-records-first-case-coronavirus-4 Accessed June 17, 2020.

S10. No author listed. Mandatory wearing of face masks will soon be enforceable. Antigua Nice. April 6, 2020. Available from: http://www.antiguanice.com/v2/client.php?id=943&news=12425 Accessed May 29, 2020.

S11. No author listed. Argentina: Health Alert. US Embassy in Argentina. April 18, 2020. Available from:https://ar.usembassy.gov/argentina-health-alert-8/ Accessed July 6, 2020.

S12. Harutyunyan A. Wearing face masks in public transport to be compulsory in Armenia starting May 18. Armen press. May 14, 2020. Available from:https://armenpress.am/eng/news/1015331/ Accessed July 6, 2020.

S13. ORF. Regierung verschärft Maßnahmen. March 30, 2020. Available from: https://orf.at/stories/3159909/ Accessed May 15, 2020.

S14. No author listed. Austria widening face-mask requirement while loosening lockdown. Reuters. April 6, 2020. Available from:https://www.reuters.com/article/health-coronavirus-austria-masks/austria-widening-face-mask-requirement-while-loosening-lockdown-idUSV9N28601G Accessed May 15, 2020.

S15. No author listed. People not using medical masks to be penalized: Raids launched. AZTV. May 8, 2020. Available from:http://www.aztv.az/en/news/6905/people-not-using-medical-masks-to-be-penalized-raids-launched Accessed July 6, 2020.

S16. No author listed. Bahamas PM’s National Press Conference: Update on COVID-19 Response – April 19th, 2020. Eleuthera News. Available from:http://eleutheranews.com/?p=22315 Accessed July 6, 2020.

S17. No author listed. Bahrain makes face masks compulsory in public, allows shops to re-open. Arab News. April 9, 2020. Available from:https://www.arabnews.com/node/1656026/middle-east Accessed May 30, 2020.

S18. Paul R. Bangladesh confirms its first three cases of coronavirus. Reuters. March 8, 2020. Available from:https://www.reuters.com/article/us-health-coronavirus-bangladesh-idUSKBN20V0FS Accessed August 2, 2020.

S19. No author listed. Coronavirus: Bangladesh confirms first death, 4 new cases. Bangla News. March 18, 2020. Available from:https://web.archive.org/web/20200326113612/ https://www.banglanews24.com/english/national/article/83210/Coronavirus-Bangladesh-confirms-first-death-4-new-cases Accessed August 2, 2020.

S20. Broomes K. Barbados records two cases of COVID-19. Nation News. March 17, 2020. Available from: https://www.nationnews.com/nationnews/news/244434/barbados-records-covid-19 Accessed August 2, 2020.

S21. No author listed. Shopping Schedule During COVID -19 Curfew. Barbados Government Information Service. April 11, 2020. Available from:https://gisbarbados.gov.bb/blog/shopping-schedule-during-covid-19-curfew/ Accessed July 8, 2020.

S22. No author listed. COVID-19: Curfew extended for Barbados, but with relaxed restrictions. April 11, 2020. Available from:https://www.looptt.com/content/covid-19-curfew-extended-barbados-relaxed-restrictions-5 Accessed July 8, 2020.

S23. Charles J, Tavel J, Wyss J, Gamez Torres N. COVID-19 continues to surge in Latin America, Caribbean. Miami Herald. May 1, 2020, as updated May 31, 2020. Available from:https://web.archive.org/web/20200531181056/ https://www.miamiherald.com/news/nation-world/world/americas/haiti/article241249651.html Accessed July 8, 2020.

S24. No author listed. Face mask required to travel on PSVs. Nation News [Barbados]. May 12, 2020. Available from: https://www.nationnews.com/nationnews/news/245494/mask-required-travel-psvs Accessed July 6, 2020.

S25. Dellanna A. Belgian government pledges free masks for everyone as part of its COVID-19 lockdown exit strategy. Euronews. April 25, 2020. Available from:https://www.euronews.com/2020/04/25/belgian-government-pledges-free-masks-for-everyone-as-part-of-its-covid-19-lockdown-exit-s Accessed July 6, 2020.

S26. No author listed. Belize Makes Face Masks Mandatory in Public. Caribbean Culture and Lifestyle. May 2, 2020. Available from:https://medium.com/@caribbeanlifestylebelize/belize-makes-face-masks-mandatory-in-public-bd3b3f4f8e88 Accessed July 6, 2020.

S27. No author listed. More African countries confirm first coronavirus cases as Jack Ma pledges aid. Reuters. March 16, 2020. Available from:https://www.reuters.com/article/us-health-coronavirus-africa/somalia-liberia-benin-and-tanzania-confirm-first-coronavirus-cases-idUSKBN2131IA Accessed June 17, 2020.

S28. Coronavirus: Benin recommends use of face mask. Pana Press. April 6, 2020. Available from:https://www.panapress.com/Coronavirus-Benin-recommends-use-a_630636073-lang2.html Accessed June 17, 2020.

S29. Agence France Presse. Benin Orders Citizens To Don Anti-virus Masks. April 7, 2020. Available from: https://www.barrons.com/news/benin-orders-citizens-to-don-anti-virus-masks-01586263504 Accessed June 17, 2020.

S30. Benin Police Enforce Mask Wearing In Bid To Stop Virus. Agence France Presse. April 8, 2020. Available from:https://www.barrons.com/news/benin-police-enforce-mask-wearing-in-bid-to-stop-virus-01586363106 Accessed May 30, 2020.

S31. No author listed. Bhutan confirms first coronavirus case. Economic Times. March 6, 2020. Avaliable from:https://economictimes.indiatimes.com/news/international/world-news/bhutan-confirms-first-coronavirus-case/articleshow/74506428.cms Accessed June 22, 2020.

S32. Ministry of Health, Royal Government of Bhutan. When to use a mask. March 11, 2020. Available from:https://www.facebook.com/MoHBhutan/posts/when-to-use-maskcoronavirus-covid/2986288851432711/ Accessed June 22, 2020.

S33. No author listed. Bosnia confirms its first case of coronavirus. N1 News. March 5, 2020. Available from: http://ba.n1info.com/English/NEWS/a414110/Bosnia-confirms-its-first-case-of-Coronavirus.html Accessed June 14, 2020.

S34. No author listed. Bosnia’s Federation entity loosens curfew, introduced mandatory masks. N1 News. March 29, 2020. Available from:http://ba.n1info.com/English/NEWS/a420560/Bosnia-s-Federation-entity-loosens-curfew-introduced-mandatory-masks.html Accessed June 10, 2020.

S35. Health Alert: U.S. Embassy Sarajevo, Bosnia and Herzegovina. Available from: https://ba.usembassy.gov/health-alert-u-s-embassy-sarajevo-bosnia-and-herzegovina-march-30-2020/ Accessed May 6, 2020.

S36. No author listed. May 1: Face masks compulsory in public, shared spaces. Africa News [Botswana]. May 1, 2020. Available from:https://www.africanews.com/2020/07/03/botswana-president-in-self-isolation-after-namibia-trip/ Accessed July 6, 2020.

S37. No author listed. Brasil confirma primeiro caso do novo coronavírus. Folha de S. Paulo. February 26, 2020. Available from:https://www1.folha.uol.com.br/equilibrioesaude/2020/02/brasil-confirma-primeiro-caso-do-novo-coronavirus.shtml Accessed June 26, 2020.

S38. Soverall AJ. BVI residents advised on the best way to wear face masks in public. Ministry of Health & Social Development. April 28, 2020. Available from:https://bvi.gov.vg/media-centre/bvi-residents-advised-best-way-wear-face-masks-public Accessed July 7, 2020.

S39. Brunei reports more coronavirus cases. The Star. March 10, 2020. Available from:https://web.archive.org/web/20200312164055/ https://www.thestar.com.my/news/regional/2020/03/10/brunei-reports-more-coronavirus-cases Accessed July 6, 2020.

S40. No author listed. Sultan: Brunei to expand virus testing capacity with new virology lab. The Star. March 22, 2020. Available from:https://www.thestar.com.my/news/regional/2020/03/22/sultan-brunei-to-expand-virus-testing-capacity-with-new-virology-lab Accessed July 6, 2020.

S41. Borissov B. Здравният министър даде заден за задължителните маски. March 31, 2020. Available from:https://www.segabg.com/hot/category-bulgaria/gledayte-na-zhivo-vutreshniyat-i-zdravniyat-ministur-razyasnyavat-merkite/ Accessed May 15, 2020.

S42. No author listed. COVID-19 Alert: Burkina Faso Updates State of Emergency Measures as of April 20. Worldaware. April 21, 2020. Available from:https://www.worldaware.com/covid-19-alert-burkina-faso-updates-state-emergency-measures-april-20 Accessed July 6, 2020.

S43. No author listed. Coronavirus | Cabo Verde – Miscellaneous. ICLG. May 5, 2020. Available from: https://iclg.com/briefing/12092-coronavirus-cabo-verde-miscellaneous Accessed July 6, 2020.

S44. Khan S. Cambodia Confirms First Coronavirus Case. VOA Khmer. January 27, 2020. Available from:https://www.voanews.com/science-health/coronavirus-outbreak/cambodia-confirms-first-coronavirus-case Accessed May 9, 2020.

S45. Khan S. Cambodian Businesses Embrace Protective Measures Against Coronavirus. VOA Khmer. March 30, 2020. Available from:https://www.voacambodia.com/a/cambodian-businesses-embrace-protective-measures-against-coronavirus/5352023.html Accessed May 9, 2020.

S46. Deviller S, Rungjirajittranon M. Fig leaf or first defence? Deploying flimsy masks against virus. Medical Xpress. January 28, 2020. Available from:https://medicalxpress.com/news/2020-01-fig-leaf-defence-deploying-flimsy.html Accessed June 6, 2020.

S47. Unah L, Mussa C. Masks, bans and questions: Inside Cameroon’s COVID-19 response. April 23, 2020. Available from:https://www.aljazeera.com/news/2020/04/masks-bans-questions-cameroon-covid-19-response-200422134140013.html Accessed May 26, 2020.

S48. No author listed. Cameroon: Authorities announce mandatory face mask use in public places as of April 13. Garda. April 10, 2020. Available from:https://www.garda.com/crisis24/news-alerts/331246/cameroon-authorities-announce-mandatory-face-mask-use-in-public-places-as-of-april-13-update-4 Accessed July 7, 2020.

S49. Chase S. Theresa Tam offers new advice: Wear a non-medical face mask when shopping or using public transit. The Globe and Mail. April 6, 2020. Available from:https://www.theglobeandmail.com/canada/article-tam-offers-new-advice-wear-a-non-medical-mask-when-shopping-or-using/ Accessed May 7, 2020.

S50. No author listed. Wearing Masks & Face Coverings In The Cayman Islands. Cayman Islands Government April 6 2020. Available from:https://www.exploregov.ky/coronavirus-blog/when-why-to-wear-a-mask Accessed July 7, 2020.

S51. Young K. Cayman embraces homemade face masks. Cayman Compass. April 1, 2020. Available from: https://www.caymancompass.com/2020/04/01/cayman-embraces-homemade-face-masks/ Accessed July 7, 2020.

S52. No author listed. Chad confirms first case of coronavirus: government statement. Reuters. March 19, 2020. Available from:https://www.reuters.com/article/us-health-coronavirus-chad/chad-confirms-first-case-of-coronavirus-government-statement-idUSKBN2162LO Accessed July 7, 2020.

S53. Presidence de la Republique du Tchad. Communique du Gouvernement. April 13, 2020. Available from:https://www.presidence.td/fr-news-4198.html Accessed July 7, 2020.

S54. No author listed. Chad: Authorities announce nationwide COVID-19 restrictions from May 7. Garda. May 11, 2020. Available from:https://www.garda.com/crisis24/news-alerts/340916/chad-authorities-announce-nationwide-covid-19-restrictions-from-may-7-update-9 Accessed July 7, 2020.

S55. No author listed. Chad Isolates Capital N’Djamena Due To Coronavirus. Barron’s. May 6, 2020. Available from:https://www.barrons.com/news/chad-isolates-capital-n-djamena-due-to-coronavirus-01588790404 Accessed August 2, 2020.

S56. No author listed. In several countries in central Africa, the mask becomes mandatory. Times Famous. April 15, 2020. Available from:https://timesfamous.com/worldnews/dans-plusieurs-pays-dafrique-centrale-le-masque-devient-obligatoire/ Accessed July 7, 2020.

S57. No author listed. Chile afirma estar en una ‘guerra’ por los recursos contra el COVID-19. Diario Libre. April 6, 2020. Available from:https://www.diariolibre.com/actualidad/internacional/chile-afirma-estar-en-una-guerra-por-los-recursos-contra-el-covid-19-OJ18118776 Accessed May 7, 2020.

S58. Wu H, Yu S. Masks on, Chinese start holiday travels as alarm mounts over mystery virus. Reuters. Jan. 20, 2020. Available from:https://www.reuters.com/article/us-china-health-pneumonia-masks/masks-on-chinese-start-holiday-travels-as-alarm-mounts-over-mystery-virus-idUSKBN1ZJ0VU Accessed May 26, 2020.

S59. No author listed. All people in public places in Wuhan required to wear masks, local government says. China Daily. Jan. 22, 2020. Available from: http://www.chinadaily.com.cn/a/202001/22/WS5e285cada310128217272d84.html Accessed May 26, 2020.

S60. Epidemic Prevention and Control Group. CDC. Notice regarding the issuance of guidelines for the protection of people with different risks of new coronavirus infection and guidelines for the use of pneumonia masks for the prevention of new coronavirus infection. Jan. 31, 2020. Available from: http://www.nhc.gov.cn/jkj/s7916/202001/a3a261dabfcf4c3fa365d4eb07ddab34.shtml Accessed May 26, 2020.

S61. No author listed. Wuhan doctor, colleague of virus whistleblower Li Wenliang, dies from coronavirus. Global Times. March 9, 2020. Available from: https://www.globaltimes.cn/content/1182009.shtml Accessed July 31, 2020.

S62. No author listed. Which countries have made wearing face masks compulsory? Al Jazeera News. May 5, 2020. Available from:https://www.aljazeera.com/news/2020/04/countries-wearing-face-masks-compulsory-200423094510867.html Accessed May 7, 2020.

S63. Hide S. Coronavirus in Colombia: April 3 update. Bogota Post. April 3, 2020. Available from:https://thebogotapost.com/coronavirus-in-colombia-april-3-update/45493/ Accessed June 24, 2020.

S64. Peckham R. Colombia President Ivan Duque announced April 4 that his administration likely will decide early this week whether and how strict the current national Coronavirus quarantine will extend beyond the presumptive April 13 expiration. Medellin Herald. April 5, 2020. Available from:https://www.medellinherald.com/ln/item/861-colombia-president-to-define-post-april-13-quarantine-measures-this-week-mask-mandate-now-national Accessed June10, 2020.

S65. Hide S. Coronavirus in Colombia: April 5 update. Bogota Post. April 5, 2020. Available from:https://thebogotapost.com/coronavirus-in-colombia-april-5-update/45624/ Accessed June 24, 2020.

S66. Shaban AR. Congo Republic extends coronavirus lockdown. Africa News. May 2, 2020. Available from:https://www.africanews.com/2020/05/02/congo-republic-extends-coronavirus-lockdown// Accessed July 6, 2020.

S67. Rogulj D. COVID-19 in Croatia: Recommendations for Public Transportation. Croatia News. April 25, 2020. Available from:https://www.total-croatia-news.com/travel/43138-croatia Accessed July 6, 2020.

S68. Cuba Suspends International Passenger Flights. Xinhua in Telesur. Available from:https://www.telesurenglish.net/news/Cuba-Suspends-International-Passenger-Flights-20200402-0012.html Accessed May 29, 2020.

S69. Agapiou G. Coronavirus: govt adviser says imperative to wear masks in public spaces. Cyprus Mail. April 3, 2020. Available from:https://cyprus-mail.com/2020/04/03/coronavirus-govt-adviser-says-imperative-to-wear-masks-in-public-spaces/ Accessed May 7, 2020.

S70. No author listed. V česku jsou tři lidé nakažení koronavirem. Předtím byli v Itálii. čT24. česka Televize 24. March 1, 2020. Available from:https://ct24.ceskatelevize.cz/domaci/3056228-v-cesku-jsou-tri-lide-nakazeni-koronavirem Accessed June 14, 2020.

S71. Veronika B. Could Czech’s Measure to Fight Coronavirus Save Thousands of Lives? Prague Morning. April 4, 2020. Available from:https://www.praguemorning.cz/could-czechs-measure-to-fight-coronavirus-save-thousands-of-lives-2/ Accessed May 7, 2020.

S72. No author listed. Republic of Djibouti: COVID-19 Situation Report #7, 10 May 2020. Reliefweb. 11 May 2020. Available from:https://reliefweb.int/report/djibouti/republic-djibouti-covid-19-situation-report-7-10-may-2020 Accessed July 6, 2020.

S73. Baptiste D. Dominica records first case of coronavirus. Loop Jamaica. March 22, 2020. Available from:http://www.loopjamaica.com/content/watch-dominica-records-first-case-coronavirus-3 Accessed July 6, 2020.

S74. No author listed. COVID-19: Dominica’s president welcomes bi-partisan support for state of emergency and curfew extension. Dominica News Online. April 9, 2020. Available from:https://dominicanewsonline.com/news/homepage/homepage-carousel/covid-19-dominicas-president-welcomes-bi-partisan-support-for-state-of-emergency-and-curfew-extension/ Accessed July 6, 2020.

S75. No author listed. COVID-19 is a global war says Dominican PM Dr Skerrit on DBS. Wic News. March 31, 2020. Available from:https://wicnews.com/caribbean/dominica/covid-19-global-war-says-dominican-pm-dr-skerrit-dbs-291326849/ Accessed July 6, 2020.

S76. King A. Guidance for wearing, removing, and discarding protective masks. GIS Dominica. April 7, 2020. Available from: https://m.facebook.com/watch/?v=867718350318637&_rdr Accessed July 6, 2020.

S77. No author listed. 3 Covid cases left in Dominica. Dominica Daily. April 25, 2020. Available from: https://www.thedominicadaily.com/2020/04/3-covid-cases-left-in-dominica.html Accessed July 6, 2020.

S78. Dominica has done well in preventing the spread of COVID-19 – Dr. Sam Christian. Dominica News Online. April 21, 2020. Available from:https://dominicanewsonline.com/news/homepage/homepage-carousel/sam-christian-says-dominica-has-done-well-in-preventing-the-spread-of-covid-19/ Accessed July 6, 2020.

S79. No author listed. Dominica: International organizations caution against stigmatization of people affected by COVID-19. International Organization for Migration. April 20, 2020. Available from:https://www.programamesoamerica.iom.int/en/news/dominica-international-organizations-caution-against-stigmatization-people-affected-covid-19 Accessed July 7, 2020.

S80. No author listed. Nine recover from COVID-19 in Dominica. Dominica News Online. April 22, 2020. Available from:https://dominicanewsonline.com/news/homepage/news/nine-recover-from-covid-19-in-dominica/ Accessed July 7, 2020.

S81. No author listed. In the absence of medical face masks, headscarves, recommends Minister of Public Health. Dominican Today. April 6, 2020. Available from:https://dominicantoday.com/dr/local/2020/04/06/in-the-absence-of-medical-face-masks-headscarves-recommends-minister-of-public-health/ Accessed July 7, 2020.

S82. No author listed. El uso de mascarillas en el país será obligatorio en espacios públicos y lugares de trabajo. Listin Diario. April 16, 2020. Available from:https://listindiario.com/economia/2020/04/16/613491/el-uso-de-mascarillas-en-el-pais-sera-obligatorio-en-espacios-publicos-y-lugares-de-trabajo Accessed May 7, 2020.

S83. No author listed. Ministerio de Salud confirma primer caso de coronavirus en Ecuador. El Comercio. February 29, 2020. Available from:https://www.elcomercio.com/actualidad/salud-confirma-primer-caso-coronavirus.html Accessed June 27, 2020.

S84. No author listed. Ecuador confirma primera muerte por coronavirus. Reuters. March 13, 2020. Available from:https://www.infobae.com/america/agencias/2020/03/13/ecuador-confirma-primera-muerte-por-coronavirus/ Accessed June 27, 2020.

S85. Armus T. Bodies of coronavirus victims are left on the streets in Ecuador’s largest city. San Antonio News Express. April 3, 2020. Available from:https://www.expressnews.com/news/article/Bodies-of-coronavirus-victims-are-left-on-the-15176423.php Accessed June 24, 2020.

S86. No author listed. Face masks now required in public; New virus cases trend down; Prostitution ruled non-essential; Ecuador, Peru target border crossings. Cuenca High Life. April 7, 2020. Available from:https://cuencahighlife.com/face-masks-now-required-in-public-new-virus-cases-trend-down-prostitution-ruled-non-essential-ecuador-peru-target-border-crossings/ Accessed June 24, 2020.

S87. Fouly M. Feature: Young Egyptians launch campaign to provide face masks, raise coronavirus awareness in streets. Xinhua. March 20, 2020. Available from:http://www.xinhuanet.com/english/2020-03/21/c_138902950.htm Accessed June 19, 2020.

S88. Morsi A. Masks in public places: ‘Even a piece of cloth will do’. Ahram Online. June 2, 2020. Available from:http://english.ahram.org.eg/NewsContent/50/1201/370399/AlAhram-Weekly/Egypt/Masks-in-public-places-%E2%80%98Even-a-piece-of-cloth-will.aspx Accessed July 7, 2020.

S89. Gómez R. Primer caso de COVID-19 en El Salvador pudo haber entrado por punto ciego en Metapán. La Prensa Grafica. March 18, 2020. Available from:https://www.laprensagrafica.com/elsalvador/Primer-caso-de-COVID-19-en-El-Salvador-pudo-haber-entrado-por-punto-ciego-en-Metapan-se-ha-activado-cerco-sanitario-por-48-horas-en-ese-municipio-20200318-0064.html

S90. Corchado A. El Salvador duplicará pruebas para detectar coronavirus. AS. April 4, 2020. Available from:https://us.as.com/us/2020/04/05/tikitakas/1586044839_590985.html Accessed July 8, 2020.

S91. Delcid M. Disponen el uso obligatorio de mascarillas en San Salvador para prevenir coronavirus. CNN Espanol. April 8, 2020. Available from:https://cnnespanol.cnn.com/2020/04/08/disponen-el-uso-obligatorio-de-mascarillas-en-san-salvador-para-prevenir-coronavirus/ Accessed July 8, 2020.

S92. No author listed. Coronavirus.-El Salvador obliga al uso de mascarillas fuera de las viviendas por el coronavirus. Notiamerica. April 12, 2020. Available from:https://www.notimerica.com/sociedad/noticia-coronavirus-salvador-obliga-uso-mascarillas-fuera-viviendas-coronavirus-20200412072353.html Accessed July 8, 2020.

S93. Health Alert: Equatorial Guinea, Government Extends COVID-19 Containment Measures. Overseas Security Advisory Council. April 15, 2020. Available from:https://www.osac.gov/Country/EquatorialGuinea/Content/Detail/Report/f079c408-dbc7-421c-b6b1-1873c40040dd Accessed May 7, 2020.

S94. No author listed. Prime minister: We are unfortunately still in coronavirus deepening phase. ERR News. April 5, 2020. Available from:https://news.err.ee/1073236/prime-minister-we-are-unfortunately-still-in-coronavirus-deepening-phase Accessed June 24, 2020.

S95. Samuel G. Ethiopia Outlaws Handshakes, Obliges Masks in Public Places. Addis Fortune. April 12, 2020. Available from:https://addisfortune.news/ethiopia-outlaws-handshakes-obliges-masks-in-public-places/ Accessed May 30, 2020.

S96. Finland encourages use of face masks in policy turnaround. Medical Press. April 14, 2020. Available from: https://medicalxpress.com/news/2020-04-finland-masks-policy-turnaround.html Accessed May 7, 2020

S97. 213X. Coronavirus en France : le parcours des trois patients. Franceinfo. January 25, 2020. Available from:https://www.francetvinfo.fr/sante/maladie/coronavirus/coronavirus-en-france-le-parcours-des-trois-patients_3799837.html Accessed June 20, 2020.

S98. 214X. No author listed. Coronavirus: First death confirmed in Europe. BBC News. February 15, 2020. Available from:https://www.bbc.com/news/world-europe-51514837 Accessed June 20, 2020.

S99. 215X. Coronavirus: U-turn on face masks in France and the United States. France 24. April 4, 2020. Available from:https://www.france24.com/en/20200404-new-york-is-in-a-race-against-time-against-virus-as-trump-says-masks-are-voluntary Accessed June 20, 2020.

S100. Gambia: COVID-19 and the mad rush for face masks! African Press Agency News. March 18, 2020. Available from: http://apanews.net/mobile/uneInterieure_EN.php?id=4937708 Accessed May 7, 2020.

S101. Gambia Ministry of Health. The Gambia COVID-19 Outbreak Situational Report. May 26, 2020. Available from:http://www.moh.gov.gm/wp-content/uploads/2020/05/Gambia_The_COVID-19_Sitreps-26-05-2020.pdf Accessed June 10, 2020.

S102. U.S. Embassy in Georgia. Alert: Prohibition on car traffic starting 12 noon Friday, April 17. April 17, 2020. Available from:https://ge.usembassy.gov/alert-prohibition-on-car-traffic-starting-12-noon-friday-april-17/ Accessed July 7, 2020.

S103. No author listed. Bayerische Behörden bestätigen ersten Fall in Deutschland. Der Spiegel. January 28, 2020. Available from:https://www.spiegel.de/wissenschaft/medizin/corona-virus-erster-fall-in-deutschland-bestaetigt-a-19843b8d-8694-451f-baf7-0189d3356f99 Accessed June 20, 2020.

S104. Chambers M. German city introduces face masks for shoppers as coronavirus spreads. Reuters. March 31, 2020. Available from:https://www.reuters.com/article/us-health-coronavirus-germany-masks/german-city-introduces-face-masks-for-shoppers-as-coronavirus-spreads-idUSKBN21I10K Accessed May 7, 2020.

S105. No author listed. ‘They could reduce the risk’: Germany’s public health institute updates stance on face masks. The Local de. April 2, 2020. Available from:https://www.thelocal.de/20200402/latest-face-masks-in-public-could-help-to-reduce-spread-of-coronavirus-says-germanys-robert-koch-institute Accessed June 20, 2020.

S106. No author listed. Coronavirus: Germany’s states make face masks compulsory. BBC News. April 22, 2020. Available from: https://www.bbc.com/news/world-europe-52382196 Accessed July 7, 2020.

S107. No author listed. Every Ghanaian must wear masks. ETV Ghana. April 20, 2020. Available from: https://www.etvghana.com/every-ghanaian-must-wear-masks-nana-addo/ Accessed July 6, 2020.

S108. Ghana Health Service. Ministerial directive on wearing masks in public places to prevent transmission of COVID-19. Ghana Health Service. April 25, 2020. Available from: https://ghanahealthservice.org/covid19/downloads/covid_19_nose_mask.pdf Accessed June 29, 2020.

S109. No author listed. ςτους 136 οι νεκροί – Υποχρεωτική η χρήςη μάςκας ςε ΜΜΜ και κλειςτούς χώρους. The Press Project. April 27, 2020. Available from:https://thepressproject.gr/stous-136-i-nekri-ypochreotiki-i-chrisi-maskas-se-mmm-ke-klistous-chorous/ Accessed July 7, 2020.

S110. Wong M. Grenada records first COVID-19 case. loopnewsbarbados. March 22, 2020. Available from:http://www.loopnewsbarbados.com/content/grenada-records-first-covid-19-case-4 Accessed June 17, 2020.

S111. Ministry of Health Grenada. Be COVID-19 smart—wear a mask. April 3, 2020. Available from:https://www.facebook.com/HealthGrenada/posts/be-covid19-smart-wear-a-maskmasks-are-effective-in-helping-to-slow-the-spread-of/738937303305031/ Accessed June 17, 2020.

S112. Cabinet of Grenada. Emergency Powers (Covid-19) (No. 3) Regulations, 2020. April 6, 2020. Available from:https://www.nowgrenada.com/2020/04/emergency-powers-covid-19-no-3-regulations-2020/ Accessed June 17, 2020.

S113. Health Alert: Guatemala, Government Mandates Wearing of Masks in Public Spaces. Oversea Security Advisory Council. April 9, 2020. Available from: https://www.osac.gov/Country/Guatemala/Content/Detail/Report/01cbc8c3-6795-4fc8-a941-1867c560f0df Accessed May 7, 2020.

S114. Conde A. #COVID19 Le port de masque communautaire ou bavette est maintenant OBLIGATOIRE pour tout citoyen à compter du samedi 18 avril 2020. Twitter. April 13, 2020. Available from: https://twitter.com/alphacondepresi/status/1249793942368980996 Accessed July 18, 2020.

S115. No author listed. Guinea-Bissau’s president extends state of emergency untill May 26. Xinhua. May 11, 2020. Available from:http://www.xinhuanet.com/english/2020-05/12/c_139048499.htm Accessed July 7, 2020.

S116. Charles J, Tavel J, Wyss J, Gamez Torres N. Latin America and Caribbean yet to hit coronavirus surge; most are tightening measures. Miami Herald. April 17, 2020. Available from:https://web.archive.org/web/20200418093401/ https://www.miamiherald.com/news/nation-world/world/americas/haiti/article241249651.html Accessed July 7, 2020.

S117. Charles J. Prime minister orders Haitians to wear masks in public and orders firms to make more masks. Miami Herald. May 5, 2020. Available from:https://www.miamiherald.com/news/nation-world/world/americas/haiti/article242507476.html Accessed July 8, 2020.

S118. Charles J, Tavel J, Wyss J, Gamez Torres N. As region eases restrictions on fighting coronavirus, PAHO expresses concerns. Miami Herald. May 01, 2020 [May 7 update] Available from:https://web.archive.org/web/20200516122255/ https://www.miamiherald.com/news/nation-world/world/americas/haiti/article241249651.html Accessed July 8, 2020.

S119. Archeta K. Honduras makes it mandatory to wear face masks in public. COVID-19 World News. April 7, 2020. Available from:https://covid19data.com/2020/04/07/honduras-makes-it-mandatory-to-wear-face-masks-in-public/ Accessed May 7, 2020.

S120. Department of Health, Hong Kong. Latest recommendations by Scientific Committee on Emerging and Zoonotic Diseases and Scientific Committee on Infection Control after reviewing cases of novel coronavirus infection. January 24, 2020. Available from: https://www.info.gov.hk/gia/general/202001/24/P2020012400762.htm Accessed May 9, 2020.

S121. Cheung E. China coronavirus: death toll almost doubles in one day as Hong Kong reports its first two cases. South China Morning Post. January 22, 2020. Available from:https://www.scmp.com/news/hong-kong/health-environment/article/3047193/china-coronavirus-first-case-confirmed-hong-kong Accessed May 9, 2020.

S122. Wu P, Tsang TK, Wong JY, et al. Suppressing COVID-19 transmission in Hong Kong: an observational study of the first four months. Available from:https://assets.researchsquare.com/files/rs-34047/v1/e26fcc0f-8101-4007-9c2a-6fb1b24bc8ea.pdf Accessed June 19, 2020.

S123. Szakacs G. Hungary eases coronavirus restrictions outside Budapest. National Post. April 29, 2020. Available from:https://nationalpost.com/pmn/health-pmn/hungary-eases-coronavirus-restrictions-outside-budapest-pm Accessed July 8, 2020.

S124. Reid D. India confirms its first coronavirus case. CNBC. January 30, 2020. Available from:https://www.cnbc.com/2020/01/30/india-confirms-first-case-of-the-coronavirus.html Accessed June 19, 2020.

S125. Thacker T. Amid outbreak, Health Ministry recommends homemade face masks. Economic times. Apr 4, 2020. Available from:https://economictimes.indiatimes.com/industry/healthcare/biotech/healthcare/amid-outbreak-health-ministry-recommends-homemade-face-masks/articleshow/74978175.cms Accessed May 29, 2020.

S126. Soeriaatmadja W. Coronavirus: Price of a box of N95 masks cost more than a gram of gold in Indonesia. Straits Times. February 10, 2020. Available from:https://www.straitstimes.com/asia/se-asia/coronavirus-price-of-a-box-n95-masks-cost-more-than-a-gram-of-gold-in-indonesia

S127. No author listed. Indonesia confirms first cases of coronavirus. Bangkok Post. March 2, 2020. Available from:https://www.bangkokpost.com/world/1869789/indonesia-confirms-first-cases-of-coronavirus Accessed June 19, 2020.

S128. Afriyadi AD. Telkom Buka Suara Ada Karyawannya Meninggal Positif Corona. Detik Finance. March 16, 2020. Available from:https://finance.detik.com/berita-ekonomi-bisnis/d-4940364/telkom-buka-suara-ada-karyawannya-meninggal-positif-corona Accessed June 19, 2020.

S129. Yulisman L. Coronavirus: Indonesia makes face masks compulsory as death toll nears 200. Straits Times. April 5, 2020. Available from:https://www.straitstimes.com/asia/se-asia/coronavirus-indonesia-orders-citizens-to-wear-masks-as-infections-rise Accessed May 29, 2020.

S130. No author listed. Two Iranians die after testing positive for coronavirus. Reuters. February 19, 2020. Available from:https://www.cnbc.com/2020/02/19/two-iranians-die-after-testing-positive-for-coronavirus.html Accessed June 27, 2020.

S131. Borger J. Satellite images show Iran has built mass graves amid coronavirus outbreak. The Guardian. March 12, 2020. Available from:https://www.theguardian.com/world/2020/mar/12/coronavirus-iran-mass-graves-qom Accessed June 27, 2020.

S132. Wright R. How Iran became a new epicenter of the coronavirus outbreak. The New Yorker. 2020 Feb 28.

S133. No author listed. Covid-19: Iraq announces changes to curfew, other restrictions. Government of Iraq. April 20, 2020. Available from:https://gds.gov.iq/covid-19-iraq-announces-changes-to-curfew-other-restrictions/ Accessed July 8, 2020.

S134. No author listed. Taoiseach advises public to wear face coverings on public transport and in retail stores. JournalIE. May 15, 2020. Available from:https://www.thejournal.ie/face-coverings-on-public-transport-and-in-retail-outlets-5098881-May2020/ Accessed July 8, 2020.

S135. Staff T. Netanyahu urges wearing masks outside; announces stipends for kids, elderly. The Times of Israel. April 1, 2020. Available from:https://www.timesofisrael.com/netanyahu-tells-israelis-to-wear-masks-outside-gives-stipends-for-kids-elderly Accessed May 8, 2020.

S136. Severgnini C. Coronavirus, primi due casi in Italia «Sono due cinesi in vacanza a Roma» Sono arrivati a Milano il 23 gennaio. Corriere Della Sera. January 31, 2020. Available from:https://www.corriere.it/cronache/20_gennaio_30/coronavirus-italia-corona-9d6dc436-4343-11ea-bdc8-faf1f56f19b7.shtml?refresh_ce-cp Accessed June 20, 2020.

S137. Godin M. Why Is Italy’s Coronavirus Outbreak So Bad? Time. March 10, 2020. Available from: https://time.com/5799586/italy-coronavirus-outbreak/ Accessed June 20, 2020.

S138. No author listed. Coronavirus: Lombardy, Tuscany make face masks compulsory. April 6, 2020. ANSA. Available from:https://www.ansa.it/english/news/2020/04/06/coronavirus-lombardy-makes-face-masks-compulsory_a852ffdb-a0dd-4c55-a725-e852c5a2fc43.html Accessed June 20, 2020.

S139. No author listed. UPDATE: When and where do you need to wear a face mask in Italy? The Local IT. April 28, 2020. Available from:https://www.thelocal.it/20200428/coronavirus-where-should-you-wear-a-face-mask-in-italy Accessed July 8, 2020.

S140. No author listed. Ivory Coast confirms first case of coronavirus. Reuters. Daily Sabah. March 11, 2020. Available from:https://www.dailysabah.com/world/ivory-coast-confirms-first-case-of-coronavirus/news Accessed June 13, 2020.

S141. No author listed. Coronavirus: Ivory Coast protesters target testing centre. BBC News. April 6, 2020. Available from:https://www.bbc.com/news/world-africa-52189144 Accessed June 18, 2020.

S142. Mundle T. PM Announces Order For Wearing Of Masks. Jamaica Information Service. April 9, 2020. Available from:https://jis.gov.jm/pm-announces-order-for-wearing-of-masks/ Accessed July 12, 2020.

S143. Mundle T. All Jamaicans Must Wear A Mask In Public – PM. Jamaica Information Service. April 21, 2020. Available from: https://jis.gov.jm/all-jamaicans-must-wear-a-mask-in-public-pm/ Accessed July 8, 2020.

S144. Takahashi. Amid virus outbreak, Japan stores scramble to meet demand for face masks. Japan Times. Available from:https://www.japantimes.co.jp/news/2020/01/31/national/coronavirus-japan-surgical-masks/#.Xrc8Z2hKhPY Accessed May 9, 2020.

S145. Kawai Y. New China virus spurs 24-hour output of surgical masks in Japan. Nikkei Asian Review. January 18, 2020. Available from:https://asia.nikkei.com/Business/Business-trends/New-China-virus-spurs-24-hour-output-of-surgical-masks-in-Japan Accessed June 6, 2020.

S146. No author listed. Face masks and hand sanitizers in short supply across Asia amid deadly virus panic. Japan Times. January 24, 2020. Available from:https://www.japantimes.co.jp/news/2020/01/24/business/face-masks-hand-sanitizers-asia-coronavirus/#.Xtv0JWhKhPY Accessed June 6, 2020.

S147. No author listed. King meets with governors, urges alleviating burden on citizens by monitoring prices. Jordan Times. April 27, 2020. Available from:https://www.jordantimes.com/news/local/king-meets-governors-urges-alleviating-burden-citizens-monitoring-prices Accessed July 8, 2020.

S148. No author listed. Epidemiological situation in Jordan is stable. Jordan Times. April 29, 2020. Available from:https://menafn.com/1100093174/Epidemiological-situation-in-Jordan-is-stable Accessed July 8, 2020.

S149. No author listed. Wearing face masks in public places obligatory – President. Kazinform. May 26, 2020. Available from:https://www.inform.kz/en/wearing-face-masks-in-public-places-obligatory-president_a3654072 Accessed July 8, 2020.

S150. Ministry of Health, Republic of Kenya. First case of coronavirus disease confirmed in Kenya. March 13, 2020. Available from: https://www.health.go.ke/first-case-of-coronavirus-disease-confirmed-in-kenya/ Accessed June 20, 2020.

S151. Munde C. First Kenyan dies of Covid-19 - CS Kagwe. The Star. March 26, 2020. Available from:https://www.the-star.co.ke/covid-19/2020-03-26-first-kenyan-dies-of-covid-19-cs-kagwe/ Accessed June 20, 2020.

S152. Muraya J. Masks Are Not Optional, Kagwe Tells Passengers As Virus Threat Heightened. Capitol News. April 3, 2020. Available from:https://www.capitalfm.co.ke/news/2020/04/masks-are-not-optional-kagwe-tells-passengers-as-virus-threat-heightened/ Accessed June 20, 2020.

S153. Muraya J. Kenya: Masks Now Mandatory in Public Places, Kenya Declares. All Africa. April 5, 2020. Available from:https://allafrica.com/stories/202004060049.html Accessed May 29, 2020.

S154. Austrian K, Abuya T. We wanted to know how coronavirus affects Nairobi’s slum residents. What we found. The Conversation. May 5, 2020. Available from:https://theconversation.com/we-wanted-to-know-how-coronavirus-affects-nairobis-slum-residents-what-we-found-137621 Accessed June 20, 2020.

S155. No author listed. COVID-19 alert: Kyrgyzstan lifts state of emergency in major urban areas, maintains several restrictions as of May 11. WorldAware. May 11, 2020. Available from:https://www.worldaware.com/covid-19-alert-kyrgyzstan-lifts-state-emergency-major-urban-areas-maintains-several-restrictions Accessed July 9, 2020.

S156. Uy MH. Lao morning news for March 6. AEC News. March 6, 2020. Available from: https://aecnewstoday.com/2020/lao-morning-news-for-march-6-3/ Accessed June 6, 2020.

S157. No author listed. Laos reports no case of COVID-19, to import face masks from Vietnam. Vietnam Times. March 20, 2020. Available from:https://vietnamtimes.org.vn/laos-reports-no-case-of-covid-19-to-import-face-masks-from-vietnam-18566.html Accessed May 9, 2020.

S158. No author listed. Latvian PM urges everyone to wear face masks in public places. Latvian Public Broadcasting. April 28, 2020. Available from:https://eng.lsm.lv/article/society/health/latvian-pm-urges-everyone-to-wear-face-masks-in-public-places.a357722/ Accessed July 9, 2020.

S159. Sly L. Lebanon is in a big mess. But on coronavirus, it’s doing something right. Washington Post. April 22, 2020. Available from:https://www.washingtonpost.com/world/middle_east/lebanon-is-in-a-big-mess-but-on-coronavirus-its-doing-something-right/2020/04/21/a024496a-83e0-11ea-81a3-9690c9881111_story.html Accessed June 24, 2020.

S160. No author listed. Lebanon divided over face masks in virus battle. Arab News. April 5, 2020. Available from: https://www.arabnews.com/node/1653251/middle-east Accessed May 8, 2020.

S161. Houssari N. Lebanese must wear face masks despite coronavirus lockdown transition period. Arab News. April 25, 2020. Available from:https://arab.news/nzbdk Accessed June 24, 2020.

S162. Houssari N. Lebanon issues fines to enforce wearing of face masks. Arab News. May 9, 2020. Available from:https://www.arabnews.com/node/1681831/middle-east Accessed August 2, 2020.

S163. No author listed. Pres. Weah Extends “STAY HOME” Order Under State of Emergency. Executive Mansion [Liberia]. April 24, 2020. Available from:https://www.emansion.gov.lr/2press.php?news_id=5144&related=7&pg=sp Accessed July 9, 2020.

S164. Karabacak S. Libya to impose 10-day curfew to combat COVID-19. Anadolu Agency. April 16, 2020. Available from:https://www.aa.com.tr/en/africa/libya-to-impose-10-day-curfew-to-combat-covid-19/1806641 Accessed May 8, 2020.

S165. No author listed. Measures Taken in Liechtenstein in Response to the Coronavirus Pandemic. Embassy of the Principality of Liechtenstein. July 3, 2020. Available from:http://www.liechtensteinusa.org/article/measures-taken-in-liechtenstein-in-response-to-the-coronavirus-pandemic Accessed July 9, 2020.

S166. No author listed. Lithuania to keep quarantine in place until April 13. Baltic News Network. March 26, 2020. Available from:https://bnn-news.com/lithuania-to-keep-quarantine-in-place-until-april-13-211764 Accessed June 24, 2020.

S167. Jačauskas I. Lithuanian government extends quarantine, makes facemasks mandatory. LRT English. April 8, 2020. Available from:https://www.lrt.lt/en/news-in-english/19/1161456/lithuanian-government-extends-quarantine-makes-facemasks-mandatory Accessed May 8, 2020.

S168. Oglesby K. Construction back in action, masks mandatory from Monday. Luxembourg Times. April 15, 2020. Available from:https://luxtimes.lu/luxembourg/40429-construction-back-in-action-masks-mandatory-from-monday Accessed July 9, 2020.

S169. Chung K. Macau confirms second patient infected with Chinese coronavirus. South China Morning Post. Available from:https://www.scmp.com/news/china/article/3047337/macau-confirms-second-patient-infected-chinese-coronavirus Accessed May 26, 2020.

S170. No author listed. Madagascar: COVID-19 lockdown measures begin to ease in major cities April 20. Garda World. April 21, 2020. Available from:https://www.garda.com/crisis24/news-alerts/334481/madagascar-covid-19-lockdown-measures-begin-to-ease-in-major-cities-april-20-update-6 Accessed July 9, 2020.

S171. No author listed. Just In: Malawi registers first COVID-19 death. Face of Malawi. April 7, 2020. Available from:https://www.faceofmalawi.com/2020/04/covid-19-first-death-in-malawi/ Accessed June 6, 2020.

S172. Chilunga Z. Malawi: Mutharika Urges Malawi Unity and ‘Steadfast’ in COVID-19 Fight - Announce New Measures to Stop Spread of Outbreak. Nyasa Times. April 4, 2020. Available from:https://allafrica.com/stories/202004060182.html Accessed May 29, 2020.

S173. Harun HN, Yusof TA, Solhi F. Demand for face masks, hand sanitisers soars. New Straits Times. January 30, 2020. Available from: https://www.nst.com.my/news/nation/2020/01/561250/demand-face-masks-hand-sanitisers-soars Accessed May 9, 2020.

S174. Hadi AA. Reopening Maldives: Safe resort licenses, testing tourists for COVID-19. Sun. May 19, 2020. Available from:https://en.sun.mv/60440 Accessed July 10, 2020.

S175. No author listed. Health Alert – U.S. Embassy Bamako, Mali – May 11, 2020. Available from:https://ml.usembassy.gov/health-alert-u-s-embassy-bamako-mali-may-11-2020/ Accessed July 10, 2020.

S176. No author listed. Masks to be required when shopping or on the bus. Times of Malta. May 1, 2020. Available from:https://timesofmalta.com/articles/view/masks-to-be-required-when-shopping-or-on-the-bus.789347 Accessed July 10, 2020.

S177. No author listed. Mauritania Eases Pandemic Restrictions. Barrons. May 7, 2020. Available from: https://www.barrons.com/news/mauritania-eases-pandemic-restrictions-01588842004 Accessed July 10, 2020.

S178. Réouverture des supermarchés et boutiques : voici ce qu’il faut retenir. Le DefiMedia Group. March 31, 2020. Available from:https://defimedia.info/reouverture-des-supermarches-et-boutiques-voici-ce-quil-faut-retenir Accessed May 8, 2020.

S179. Département de Mayotte. Covid-19: un masque pour tous les Mahorais. May 11, 2020. Available from:https://www.facebook.com/conseildepartementalMayotte/posts/1615212958643116/ Accessed July 10, 2020.

S180. No author listed. Face masks are not necessarily effective, nor are they convenient. Coronavirus point man repeats reservations over value of face masks. Mexico News Daily. April 28, 2020. Available from:https://mexiconewsdaily.com/news/coronavirus/coronavirus-point-man-repeats-reservations-over-value-of-face-masks/ Accessed July 10, 2020.

S181. Mexico announces ‘new normality’ in plan to reopen economy. Aljazeera. May 13, 2020. Available from:https://www.aljazeera.com/news/2020/05/mexico-announces-normality-plan-reopen-economy-200512231541957.html Accessed July 10, 2020.

S182. No author listed. COVID-19 Information. U.S. Embassy in Moldova. July 9, 2020. Available from: https://md.usembassy.gov/u-s-citizen-services/covid-19-information/ Accessed July 10, 2020.

S183. Baljmaa. T. SEC: MNT 150,000 fine for not wearing face masks. April 14, 2020. Available from: https://www.montsame.mn/en/read/222259 Accessed May 9, 2020.

S184. Amarsaikhan S. Ulaanbaatar City Mayor’s Order A/108. January 27, 2020. Available from:https://ulaanbaatar.mn/Home/Docdetail?dataID=47003 Accessed August 1, 2020.

S185. Stjepčević A. COVID-19 in Montenegro: No New Daily Cases, Update April 30, 2020. Total Montenegro News. April 30, 2020. Available from:https://www.total-montenegro-news.com/news/5504-covid-19-in-montenegro-no-new-daily-cases-update-april-30-2020 Accessed July 10, 2020.

S186. Farrell JE. COVID 19 Weekly Message by Premier, Hon. Joseph E. Farrell [of Montserrat] – Measures from May 1 -May 7, 2020. April 29, 2020. Available from:http://www.gov.ms/covid-19-weekly-message-by-premier-hon-joseph-e-farrell-measures-from-may-1-may-7-2020/ Accessed July 10, 2020.

S187. Morocco makes face masks compulsory due to coronavirus. Reuters. April 6, 2020. Available from:https://www.reuters.com/article/us-health-coronavirus-morocco/morocco-makes-face-masks-compulsory-due-to-coronavirus-idUSKBN21O31E Accessed May 8, 2020.

S188. Mozambique confirms first coronavirus case. National Post. Available from: https://nationalpost.com/pmn/health-pmn/mozambique-confirms-first-coronavirus-case Accessed June 6, 2020.

S189. No author listed. Just In: Coronavirus. Mozambique announces first Covid-19 patient recovery. Club of Mozambique. April 4, 2020. Available from:https://clubofmozambique.com/news/just-in-coronavirus-mozambique-announces-first-covid-19-patient-recovery-156979/ Accessed June 19, 2020.

S190. No author listed. Mozambique: Government Orders Wearing of Masks. allAfrica. April 9, 2020. Available from:https://allafrica.com/stories/202004091000.html Accessed May 30, 2020.

S191. Myanmar confirms first two coronavirus cases. Straits Times. March 24, 2020. Available from: https://www.straitstimes.com/asia/se-asia/myanmar-confirms-first-coronavirus-cases Accessed June 28, 2020.

S192. Nachemson A. Fears of coronavirus catastrophe as Myanmar reports first death. Al Jazeera. April 1, 2020. Available from:https://www.aljazeera.com/news/2020/06/500000-dead-coronavirus-live-updates-200628233313992.html Accessed June 28, 2020.

S193. Dissemination of Preliminary Findings of Public Compliance Survey on Ministry of Health and Sports Guidelines for COVID-19 Prevention. Ministry of Health and Sports (Myanmar). June 3, 2020. Available from:https://www.mohs.gov.mm/page/10385 Accessed July 14, 2020.

S194. Ministry of Health and Sports (Myanmar). Frequently Asked Questions on Different Types of Mask Use in COVID-19 Prevention and Control. April 5, 2020. Available from:http://mohs.gov.mm/su/gejx3143GE Accessed July 14, 2020.

S195. Htet KS. Myanmar State Counsellor makes own face mask. Myanmar Times. April 7, 2020. Available from: https://www.mmtimes.com/news/myanmar-state-counsellor-makes-own-face-mask.html Accessed July 15, 2020.

S196. No author listed. Myanmar Steps up Coronavirus Measures as Medical Profession Feels Risks. Radio Free Asia. April 16, 2020. Available from:https://www.rfa.org/english/news/myanmar/medical-profession-04162020205725.html Accessed July 10, 2020.

S197. No author listed. Use of face masks in public becomes mandatory in Namibia. Namibian Broadcasting Corporation. May 2, 2020. Available from:https://www.nbc.na/news/use-face-masks-public-becomes-mandatory-namibia.30485 Accessed July 10, 2020.

S198. No author listed. Coronavirus: Students wear face masks to school in Nepal. Manila Bulletin Online. January 29, 2020. Available from:https://www.youtube.com/watch?v=gjdykIOjqSU Accessed June 10, 2020.

S199. Nepalis rush to buy face masks amidst coronavirus outbreak but there are none available. Kathmandu Post. February 3, 2020. Available from:http://webcache.googleusercontent.com/search?q=cache:BLTdc-faXCkJ https://kathmandupost.com/national/2020/02/03/nepalis-rush-to-buy-face-masks-amidst-coronavirus-outbreak-but-there-are-none-available&hl=en&gl=us&strip=1&vwsrc=0 Accessed June 10, 2020.

S200. Khatiwada SA, Poudel KR. After the outbreak of novel coronavirus in China, consumption of masks in Nepali markets has significantly increased, leading to their acute shortage. Rising Nepal Daily. February 8, 2020. Available from:https://webcache.googleusercontent.com/search?q=cache:fz4AvW5wF3sJ https://risingnepaldaily.com/main-news/masks-can-do-only-so-much-to-protect-us+&cd=28&hl=en&ct=clnk&gl=us Accessed June 10, 2020.

S201. No author listed. Decisions of the High-Level Coordination Committee for the Prevention and Control of COVID-19 [Nepal]. March 25, 2020. Available from: https://webcache.googleusercontent.com/search?q=cache:Gkx0jxclr6MJ: https://us.nepalembassy.gov.np/decisions-of-the-high-level-coordination-committee-for-the-prevention-and-control-of-covid-19-2/+&cd=17&hl=en&ct=clnk&gl=us Accessed June 10, 2020.

S202. Sijapati A. Protecting Nepal’s elderly from COVID-19. Nepali Times. March 25, 2020. Available from: https://www.nepalitimes.com/banner/protecting-nepals-elderly-from-covid-19/ Accessed June 10, 2020.

S203. Pascoe R. Dutch start easing coronavirus restrictions, face masks a must on public transport. Dutch News NL. May 6, 2020. Available from:https://www.dutchnews.nl/news/2020/05/dutch-start-easing-coronavirus-restrictions-face-masks-a-must-on-public-transport/ Accessed July 10, 2020.

S204. No author listed. New Caledonia: Most COVID-19 restrictions to be lifted May 4. Garda. April 30, 2020. Available from:https://www.garda.com/crisis24/news-alerts/337661/new-caledonia-most-covid-19-restrictions-to-be-lifted-may-4-update-4 Accessed July 10, 2020.

S205. No author listed. Le couvre-feu est levé à Niamey. Niamey et les 2 Jours. May 13, 2020. Available from:https://www.niameyetles2jours.com/la-gestion-publique/sante/1305-5443-le-couvre-feu-est-leve-a-niamey Accessed July 11, 2020.

S206. Adebowale N. Coronavirus: Nigeria’s health minister recommends use of improvised face masks. Premium Times. April 14, 2020. Available from:https://www.premiumtimesng.com/news/headlines/387761-coronavirus-nigerias-health-minister-recommends-use-of-improvised-face-masks.html Accessed June 10, 2020.

S207. No author listed. North Macedonia: Face masks required in public spaces April 23. Garda World. April 23, 2020. Available from:https://www.garda.com/crisis24/news-alerts/335411/north-macedonia-face-masks-required-in-public-spaces-april-23-update-7 Accessed July 11, 2020.

S208. No author listed. Supreme Committee bans all Eid celebrations, wearing masks mandatory. Oman Daily Observer. May 18, 2020. Available from:https://www.omanobserver.om/supreme-committee-bans-all-eid-celebrations-wearing-masks-mandatory/ Accessed July 11, 2020.

S209. No author listed. Pakistan makes face masks mandatory in public. CNN. May 31, 2020. Available from:https://edition.cnn.com/world/live-news/coronavirus-pandemic-05-31-20-intl/h_1ea61f10e67d78e1644aa489f88ae7c4 Accessed July 11, 2020.

S210. No author listed. Wearing “forced” masks and gloves in Palestine and penalties for violators. Sada News. May 5, 2020. Available from:https://www.sadanews.ps/news/59116.html Accessed July 11, 2020.

S211. No author listed. Es ‘obligatorio’ usar mascarilla si va a salir de casa, según ministra consejera de Salud. TVN Noticias. April 7, 2020. Available from:https://www.tvn-2.com/nacionales/Coronavirus-en-Panama-obligatorio-usar-mascarilla-salir-casa_0_5550944856.html Accessed May 8, 2020.

S212. Kombra U. Department of Education [Papua New Guinea]. Office of the Secretary. Secretary’s Circular Instruction. No. 5 of 2020. April 24, 2020. Available from:https://covid19.info.gov.pg/files/28042020/Secretary%20Circular%20Instruction%205%20of%202020.pdf.pdf Accessed July 11, 2020.

S213. 242X. Ministerio de Salud Publica y Bienestar Social. Tapabocas de tela para uso en locales cerrados. April 7, 2020. Available from:https://www.mspbs.gov.py/portal/20722/tapabocas-de-tela-para-uso-en-locales-cerrados.html Accessed June 17, 2020.

S214. Aquino M. Mascarillas gratis: Perú decreta su uso obligatorio para enfrentar el coronavirus. Infobae April 3, 2020. Available from:https://www.infobae.com/america/agencias/2020/04/03/mascarillas-gratis-peru-decreta-su-uso-obligatorio-para-enfrentar-el-coronavirus-3/ Accessed May 8, 2020.

S215. Lasco G. Why Face Masks Are Going Viral. Sapiens. February 7, 2020. Available from:https://www.sapiens.org/culture/coronavirus-mask/ Accessed May 9, 2020.

S216. Gawlowski J, Ptak A. Vending machines selling face masks appear on Warsaw streets. Reuters. April 10, 2020. Available from:https://www.reuters.com/article/us-health-coronavirus-poland-vending/vending-machines-selling-face-masks-appear-on-warsaw-streets-idUSKCN21S1MJ Accessed June 19, 2020.

S217. Almeida H. Portugal May Require Use of Masks When It Eases Confinement. Bloomberg. April 27, 2020. Available from:https://www.bloomberg.com/news/articles/2020-04-27/portugal-may-require-masks-in-public-when-it-eases-confinement Accessed July 11, 2020.

S218. Qatar: Face masks required for several sectors and public spaces April 22. Garda World. April 23, 2020. Available from:https://www.garda.com/crisis24/news-alerts/335421/qatar-face-masks-required-for-several-sectors-and-public-spaces-april-22-update-17 Accessed July 11, 2020.

S219. No author listed. Masks compulsory in Romania when virus lockdown ends. MSN. April 22, 2020. Available from:https://www.msn.com/en-xl/europe/top-stories/masks-compulsory-in-romania-when-virus-lockdown-ends/ar-BB132Efe Accessed July 11, 2020.

S220. No author listed. В России выявили первые два случая заражения коронавирусом. TASS. 31 January 2020. Available from: https://tass.ru/obschestvo/7656549 Accessed June 26, 2020.

S221. Roache M. How Russia’s Coronavirus Outbreak Became One of the World’s Worst. Time. May 15, 2020. Available from: https://time.com/5836890/russia-coronavirus/ Accessed July 11, 2020.

S222. Tasamba J. COVID-19: Rwanda, DR Congo make mask wearing mandatory. Anadolu Agency. April 19, 2020. Available from:https://www.aa.com.tr/en/africa/covid-19-rwanda-dr-congo-make-mask-wearing-mandatory/1810165 Accessed July 11, 2020.

S223. Nurse M. St. Kitts And Nevis Confirms Two COVID-19 Cases. Caricom Today. March 25, 2020. Available from: https://today.caricom.org/2020/03/25/st-kitts-and-nevis-confirms-two-covid-19-cases/ Accessed August 2, 2020.

S224. Health Official Recommends Wearing Face Masks in Public to Slow COVID-19 Spread. The St. Kitts & Nevis Observer. April 3, 2020. Available from:https://www.thestkittsnevisobserver.com/health-official-recommends-wearing-face-masks-in-public-to-slow-covid-19-spread/ Accessed July 11, 2020.

S225. Saint Christopher and Nevis Statutory Rules and Orders. No. 12 of 2020. Emergency Powers (COVID-19) (No. 4) Regulations. April 7, 2020. Available from:https://www.covid19.gov.kn/wp-content/uploads/2020/04/SRO_12_of_2020.pdf Accessed July 11, 2020.

S226. No author listed. UK National Is Saint Lucia’s First Coronavirus Case. St. Lucia Times. March 13, 2020. Available from: https://stluciatimes.com/uk-national-is-saint-lucias-first-coronavirus-case/ Accessed August 2, 2020.

S227. No author listed. All of St Lucia’s COVID-19 patients have now recovered. Loop News. April 22, 2020. Available from: https://www.loopslu.com/content/all-st-lucias-covid-19-patients-have-now-recovered Accessed July 8, 2020.

S228. Belmar-George S. Press Statement by Chief Medical Officer, Dr. Sharon Belmar-George on the safe use of medical masks. Government of Saint Lucia. April 7, 2020. Available from: https://www.facebook.com/watch/?v=263340921365573 Accessed July 8, 2020.

229. See St. Vincent & Grenadines.

S229. No author listed. San Marino. Mascherine e guanti: quando devono essere indossati. Libertas. April 18, 2020. Available from:http://www.libertas.sm/notizie/2020/04/18/san-marino-mascherine-e-guanti-quando-devono-essere-indossati.html Accessed July 12, 2020.

S230. Covid-19: São Tomé e Príncipe com quatro casos de infecção. Radio France Internationale (in Portuguese). April 6, 2020. Available from:https://www.rfi.fr/pt/s%C3%A3o-tom%C3%A9-e-pr%C3%ADncipe/20200406-covid-19-s%C3%A3o-tom%C3%A9-e-pr%C3%ADncipe-com-quatro-casos-de-infec%C3%A7%C3%A3o Accessed July 7, 2020.

S231. No author listed. Saudi Arabia announces first case of coronavirus. Arab News. March 3, 2020. Available from:https://www.arabnews.com/node/1635781/saudi-arabia Accessed June 20, 2020.

S232. Al-Khudair D. Saudi health ministry: Face masks alone do not protect from COVID-19. Arab News. April 28, 2020. Available from:https://www.arabnews.com/node/1666456/saudi-arabia Accessed July 12, 2020.

S233. Ahmad K. Saudi Arabia Introduces New Social Distancing Rules Harpers Bazaar. May 31, 2020. Available from:https://www.harpersbazaararabia.com/culture/saudi-introduces-fine-for-not-wearing-a-face-mask-in-the-public-covid-19-coronavirus-rules Accessed July 12, 2020.

S234. No author listed. Health and Travel Alert: April 29, 2020. U.S. Embassy in Serbia. April 29, 2020. Available from:https://rs.usembassy.gov/health-and-travel-alert-april-29-2020/ Accessed July 12, 2020.

S235. No author listed. Seychelles: People in Seychelles Asked to Wear Face Masks in Crowded Areas to Prevent COVID-19. Allafrica. June 9, 2020. Available from: https://allafrica.com/stories/202006100130.html Accessed July 12, 2020.

S236. No author listed. Sierra Leone has confirmed its first case of coronavirus, president says. Reuters. March 31, 2020. Available from:https://www.reuters.com/article/us-health-coronavirus-leone-idUSKBN21I1MY Accessed June 6, 2020.

S237. No author listed. Sierra Leone announces three-day lockdown against coronavirus. Medical Xpress. April 1, 2020. Available from:https://medicalxpress.com/news/2020-04-sierra-leone-three-day-lockdown-coronavirus.html Accessed June 6, 2020.

S238. Grieco K, Yusuf Y, Meriggi N. Rapid country study: Sierra Leone. COVID-19 Series. May 2020. Available from:https://maintainsprogramme.org/wp-content/uploads/29-May-FinalV-Maintains-Covid-Rapid-Country-Report-Sierra-Leone_Revised-6.pdf Accessed June 28, 2020.

S239. Ministry of Health, Singapore. Updates on COVID-19 (Coronavirus Disease 2019) Local Situation. https://www.moh.gov.sg/covid-19 Accessed April 4, 2020.

S240. Ministry of Health, Singapore. Official Update of COVID -19 Situation in Singapore. Available from:https://experience.arcgis.com/experience/7e30edc490a5441a874f9efe67bd8b89 Accessed April 4, 2020.

S241. Sim D. Coronavirus: what’s behind Singapore’s U-turn on wearing masks? South China Morning Post. April 3, 2020. Available from:https://www.scmp.com/week-asia/health-environment/article/3078399/coronavirus-whats-behind-singapores-u-turn-wearing Accessed May 30, 2020.

S242. No author listed. Lockdown op Sint Maarten met drie weken verlengd. Antilliaans Dagblad. April 19, 2020. Available from:https://antilliaansdagblad.com/sint-maarten/21349-lockdown-op-sint-maarten-met-drie-weken-verlengd Accessed July 8, 2020.

S243. Štefúnová I. Slovensko pritvrdilo v boji s vírusom. Nosenie rúšok je povinné. Pravda. March 15, 2020. Available from:https://spravy.pravda.sk/domace/clanok/545657-slovensko-pritvrdilo-v-boji-s-virusom-nosenie-rusok-je-povinne/ Accessed May 15, 2020.

S244. Public Health Authority, Slovakia. “Opatrenie Úradu verejného zdravotníctva Slovenskej republiky pri ohrození verejného zdravia” March 24, 2020. Available from: http://www.uvzsr.sk/docs/info/covid19/Opatrenie_UVZSR_povinnost_nosit_ruska_24032020.pdf Accessed May 15, 2020.

S245. STA. COVID-19 & Slovenia, Night 29 March: Movement Restrictions, Mandatory Masks, More Aid for Individuals. Total Slovenia News. March 29, 2020. Available from:https://www.total-slovenia-news.com/politics/5951-covid-19-slovenia Accessed May 8, 2020.

S246. Human Sciences Research Council. HSRC Responds to the COVID-19 Outbreak. Available from:http://www.hsrc.ac.za/uploads/pageContent/11529/COVID-19%20MASTER%20SLIDES%2026%20APRIL%202020%20FOR%20MEDIA%20BRIEFING%20FINAL.pdf Accessed July 1, 2020.

S247. Dr Zweli Mkhize recommends the widespread use of cloth masks. Republic of South Africa Health Department. April 10, 2020. Available from:https://sacoronavirus.co.za/2020/04/10/dr-zweli-mkhize-recommends-the-widespread-use-of-cloth-masks/ Accessed May 8, 2020.

S248. No author listed. South Africa: Mandatory wearing of masks under partial easing of restrictions from May 1. April 26, 2020. Available from:https://www.garda.com/crisis24/news-alerts/336291/south-africa-mandatory-wearing-of-masks-under-partial-easing-of-restrictions-from-may-1-update-12 Accessed July 6, 2020.

S249. No author listed. With fears of Coronavirus spreading, do face masks really work? Associated Press. Available from:https://fox8.com/news/health/with-fears-of-coronavirus-spreading-do-face-masks-really-work/ Accessed June 6, 2020.

S250. Taylor K. Costco is selling out of surgical masks in South Korea, as the country battles the spread of the coronavirus. Business Insider. February 3, 2020. Available from: https://www.businessinsider.com/costco-is-selling-out-of-surgical-masks-in-south-korea-2020-2 Accessed June 6, 2020.

S251. Kim ET. How South Korea Solved Its Face Mask Shortage: Neighborhood pharmacists and government intervention were the secret weapons. New York Times. April 1, 2020. Available from: https://www.nytimes.com/2020/04/01/opinion/covid-face-maskshortage.html Accessed April 17, 2020.

S252. Ajack M. South Sudan 51st of 54 African nations to report virus case. AP News. April 5, 2020 Available from:https://apnews.com/5307b01c4db913048387a3702aefbbe2 Accessed August 2, 2020.

S253. No author listed. South Sudan: COVID-19 Update, 20 April – 03 May 2020. Reliefweb. May 3, 2020. Available from:https://reliefweb.int/report/south-sudan/south-sudan-covid-19-update-20-april-03-may-2020 Accessed July 12, 2020.

S254. No author listed. First confirmed coronavirus case in Spain in La Gomera, Canary Islands. Outbreak News Today. February 3, 2020. Available from:http://outbreaknewstoday.com/first-confirmed-coronavirus-case-in-spain-in-la-gomera-canary-islands-20628/ Accessed June 20, 2020.

S255. Alcuten J. Valencia confirma el primer muerto con coronavirus en España: un hombre de 69 años que falleció el 13 de febrero. 20 Minutos. March 3, 2020. Available from: https://www.20minutos.es/noticia/4174137/0/primer-muerto-coronavirus-espana/ Accessed June 20, 2020.

S256. Sawer P. Spain to hand out free face masks for commuters to help return to work. The Telegraph. April 11, 2020. Available from:https://www.telegraph.co.uk/news/2020/04/11/spain-hand-face-masks-allow-limited-return-work/ Accessed May 8, 2020.

S257. No author listed. Sri Lanka adopts prevention measures after first coronavirus case. Outlook. January 29, 2020. Available from:https://www.outlookindia.com/newsscroll/sri-lanka-adopts-prevention-measures-after-first-coronavirus-case/1720276 Accessed June 7, 2020.

S258. Sri Lanka makes wearing face masks mandatory when stepping out. Colombo Page. April 11, 2020. Available from: http://www.colombopage.com/archive_20A/Apr11_1586618131CH.php Accessed May 8, 2020.

S259. Keizer-Beache S. HEOC/COVID-19 Task Force Advisory: social (physical) distancing –spiritual and social wellbeing – Easter weekend. Ministry of Health, Wellness, and the Environment. Saint Vincent and the Grenadines. April 5, 2020. Available from: http://health.gov.vc/health/images/PDF/stories/HEOC_COVID-19-advisory---Easter-2020---1.pdf Accessed July 12, 2020.

S260. Keizer-Beache S. Health Services Subcommittee Advisory: Safe Use of Face Coverings – Masks. Ministry of National Security. St. Vincent and the Grenadines. April 26, 2020. Available from:http://health.gov.vc/health/images/PDF/News_Release_HEALTH_SERVICES_COMMITTEE_MASK_ADVISORY_NEOC_7_FINAL.pdf Accessed July 12, 2020.

S261. No author listed. Face mask and fruit prices soar in Sudan. Dabanga. March 16, 2020. Available from: https://www.dabangasudan.org/en/all-news/article/face-mask-and-fruit-prices-soar-in-sudan

S262. No author listed. Sudan Flash Update, 16 Mar 2020: Sudan creates two COVID-19 isolation centres in Khartoum State. Available from:https://reliefweb.int/report/sudan/sudan-flash-update-16-mar-2020-sudan-creates-two-covid-19-isolation-centres-khartoum Accessed May 26, 2020.

S263. No author listed. Breaking: Al 23 Covid-besmettingen in Suriname (update). Star Nieuws. May 31, 2020. Available from:https://www.starnieuws.com/index.php/welcome/index/nieuwsitem/58911 Accessed July 12, 2020.

S264. No author listed. Swiss parliament to debate compulsory mask requirement on Friday. The Local. June 17, 2020. Available from:https://www.thelocal.ch/20200610/will-switzerland-introduce-a-mask-requirement Accessed July 12, 2020.

S265. Blanchard B. Taiwan ups Chinese visitor curbs, to stop mask exports. Reuters. January 27, 2020. Available from:https://www.reuters.com/article/us-china-health-taiwan/taiwan-ups-chinese-visitor-curbs-to-stop-mask-exports-idUSKBN1ZQ1C6 Accessed May 9, 2020.

S266. Thongthab S. Ministry of Public Health Enhances Disease Prevention and Control. The Bangkok Insight. January 31, 2020. Available from: https://www.thebangkokinsight.com/282702/ Accessed May 9, 2020.

S267. Bureau of Information Office of the Permanent Secretary of MOPH [of Thailand]. Novel Coronavirus 2019, January 28, 2020 News Report. Available from:https://pr.moph.go.th/?url=pr/detail/2/04/137810/ Accessed June 6, 2020.

S268. Bureau of Information Office of the Permanent Secretary of MOPH [of Thailand]. Novel Coronavirus (2019-nCoV) Report, January 31, 2020. Available from:https://pr.moph.go.th/?url=pr/detail/2/04/137947/ Accessed June 6, 2020.

S269. Bureau of Information Office of the Permanent Secretary of MOPH [of Thailand]. Novel Coronavirus (2019-nCoV) News Report, January 30, 2020. Available from:https://pr.moph.go.th/?url=pr/detail/2/04/137895/ Accessed June 6, 2020.

S270. No author listed. East Timor confirms first case of coronavirus: health ministry. Reuters. March 21, 2020. Available from:https://www.reuters.com/article/us-health-coronavirus-timor/east-timor-confirms-first-case-of-coronavirus-health-ministry-idUSKBN2180BI Accessed August 2, 2020.

S271. No author listed. Health Alert – U.S. Embassy Dili, Timor-Leste (April 7, 2020). April 7, 2020. Available from:https://tl.usembassy.gov/health-alert-u-s-embassy-dili-timor-leste-april-7-2020/ Accessed July 12, 2020.

S272. No author listed. Coronavirus: Le Togo avance progressivement vers l’option du “Masque pour tous”. Togo First. April 20, 2020. Available from:https://www.togofirst.com/fr/sante/2004-5370-coronavirus-le-togo-avance-progressivement-vers-loption-du-masque-pour-tous Accessed July 12, 2020.

S273. 248X. No author listed. Wear Masks. Trinidad Express Newspaper. April 5, 2020. Available from:https://trinidadexpress.com/newsextra/wear-masks/article_4167a0d4-7765-11ea-99b6-8b1c79a31427.html Accessed May 9, 2020.

S274. No author listed. La Tunisie décide l’obligation du port du masque pour toute la population. GNet News. April 7, 2020. Available from:https://news.gnet.tn/la-tunisie-decide-lobligation-du-port-du-masque-pour-toute-la-population/ Accessed May 30, 2020.

S275. No author listed. Coronavirus: Turkey imposes curfew on youth, shuts borders of 31 cities. Middle East Eye. April 3, 2020. Available from:https://www.middleeasteye.net/news/coronavirus-turkey-erdogan-youth-under-curfew-restrictions-curb-pandemic Accessed May 9, 2020.

S276. Cartwright-Robinson S. Premier’s National COVID-19 Address. Turks and Caicos. April 30, 2020. Available from:https://www.gov.tc/moh/coronavirus/news/147-premier-s-national-covid-19-address-30-april-2020 Accessed July 14, 2020.

S277. No author listed. Museveni: Uganda’s lockdown will ease, but facemasks will be required by all. CGTN Africa. May 1, 2020. Available from:https://webcache.googleusercontent.com/search?q=cache:pmS86NKyGyQJ: https://africa.cgtn.com/2020/05/01/museveni-ugandas-lockdown-will-ease-but-facemask-mandate-will-remain/+&cd=20&hl=en&ct=clnk&gl=us Accessed July 14, 2020.

S278. No author listed. Ukraine tightens restrictions to fight coronavirus spread. Reuters. April 3, 2020. Available from:https://www.reuters.com/article/us-health-coronavirus-ukraine-measures/ukraine-tightens-restrictions-to-fight-coronavirus-spread-idUSKBN21L24L Accessed June 24, 2020.

S279. No author listed. За шашлики у парку та відсутність маски на вулиці поки не штрафуватимуть, - МВС. Zik. March 30, 2020. Available from: https://zik.ua/news/ludyna/za_shashlyky_u_parku_ta_vidsutnist_masky_na_vulytsi_poky_ne_shtrafuvatymut_mvs_963838 Accessed July 25, 2020.

S280. Nandkeolyar KH. Coronavirus in UAE: Four of a family infected. Gulf News. January 29, 2020. Available from:https://gulfnews.com/uae/health/coronavirus-in-uae-four-of-a-family-infected-1.1580273983681 Accessed June 20, 2020.

S281. Venkataraman V. Coronavirus: UAE Ministry of Health Warns Against Usage Of Masks. Curly Tales. February 29, 2020. Available from:https://curlytales.com/coronavirus-uae-ministry-of-health-warns-against-usage-of-masks/ Accessed June 20, 2020.

S282. No author listed. Health Alert: U.S. Embassy Abu Dhabi and U.S. Consulate General Dubai (March 28, 2020). Available from:https://ae.usembassy.gov/health-alert-u-s-embassy-abu-dhabi-and-u-s-consulate-general-dubai-march-28-2020/ Accessed June 20, 2020.

S283. Ball T. Hunt for contacts of coronavirus-stricken pair in York. The Sunday Times. January 31, 2020. Available from:https://www.thetimes.co.uk/article/hunt-for-contacts-of-coronavirus-stricken-pair-in-york-dh363qf8k Accessed June 20, 2020.

S284. No author listed. Coronavirus: Cover faces in some public areas, people in England told. BBC. May 11, 2020. Available from:https://www.bbc.com/news/uk-52620556 Accessed July 14, 2020.

S285. No author listed. Face coverings to be made compulsory on public transport in England. June 4, 2020. Available from:https://www.theguardian.com/world/2020/jun/04/face-masks-to-be-made-compulsory-on-public-transport-in-england Accessed July 14, 2020.

S286. Ritter Z, Brenan M. New April Guidelines Boost Perceived Efficacy of Face Masks. Gallup. May 13, 2020. Available from:https://news.gallup.com/poll/310400/new-april-guidelines-boost-perceived-efficacy-face-masks.aspx Accessed June 1, 2020.

S287. No author listed. Informe de situación en relación al coronavirus COVID-19 en Uruguay del 10/4/20. Ministerio de Salud Publica. April 10, 2020. Available from:https://www.gub.uy/sistema-nacional-emergencias/comunicacion/comunicados/informe-situacion-relacion-coronavirus-covid-19-uruguay-del-10420 Accessed July 14, 2020.

S288. No author listed. Uzbekistan confirms first coronavirus case – govt. Reuters. March 15, 2020. Available from:https://www.reuters.com/article/health-coronavirus-uzbekistan/uzbekistan-confirms-first-coronavirus-case-govt-idUSL8N2B802F Accessed June 10, 2020.

S289. COVID-19 Information. US Embassy in Uzbekistan. May 1, 2020. Available from: https://uz.usembassy.gov/covid-19-information/ Accessed May 9, 2020.

S290. AFP. Coronavirus: Maduro tells Venezuelans to make their own masks. March 17, 2020. Available from: https://www.youtube.com/watch?v=EjTsjJNCAuc Accessed May 15, 2020.

S291. Berwick A, Nava M, Kinosian S. Venezuela confirms coronavirus cases amid public health concerns. Reuters. March 14, 2020. Available from:https://www.physiciansweekly.com/venezuela-confirms-coronavirus-cases/ Accessed May 15, 2020.

S292. Valderrama S. Venezuelans sew homemade face masks amid coronavirus quarantine. Reuters. March 20, 2020. Available from:https://www.reuters.com/article/us-health-coronavirus-venezuela-masks/venezuelans-sew-homemade-face-masks-amid-coronavirus-quarantine-idUSKBN2171W2 Accessed May 9, 2020.

S293. No author listed. Venezuela confirms first coronavirus death: official. Reuters. March 26, 2020. Available from: https://www.reuters.com/article/us-health-coronavirus-venezuela-death-idUSKBN21D3Q3 Accessed July 15, 2020.

S294. Charles J, Tavel J, Wyss J, Gamez Torres N. Coronavirus update: Some of the measures taken by Latin American, Caribbean nations. Miami Herald. March 24, 2020. Available from:https://web.archive.org/web/20200325195723/ https://www.miamiherald.com/news/nation-world/world/americas/haiti/article241249651.html Accessed July 7, 2020.

S295. Taylor K. I’ve been traveling in Asia for 3 weeks amid the deadly coronavirus outbreak, and actually catching the virus is far from my biggest fear. Business Insider. February 18, 2020. Available from:https://www.businessinsider.com/travel-in-asia-during-coronavirus-outbreak-2020-2#so-for-now-my-travels-continue-17 Accessed May 9, 2020.

S296. Minh A. Coronavirus fears spur sales of face masks. VN Express. January 27, 2020. Available from:https://e.vnexpress.net/news/business/economy/coronavirus-fears-spur-sales-of-face-masks-4047111.html Accessed January 6, 2020.

S297. Vu K, Nguyen P, Pearson J. After aggressive mass testing, Vietnam says it contains coronavirus outbreak. Reuters. April 29, 2020. Available from:https://www.reuters.com/article/us-health-coronavirus-vietnam-fight-insi/after-aggressive-mass-testing-vietnam-says-it-contains-coronavirus-outbreak-idUSKBN22B34H Accessed June 13, 2020.

S298. Chisenga O, Mbulo E. Wear masks to mitigate COVID-19-Chilufya. The Mast. April 5, 2020. Available from: https://www.themastonline.com/2020/04/05/wear-masks-to-mitigate-covid-19-chilufya/ Accessed June 18, 2020.

S299. Center for International Health, Education, and Biosecurity. University of Maryland. COVID-19 Clinical Guidance and Mask Making in Zambia. Available from:http://ciheb.org/NEWS/COVID-19-Clinical-Guidance-and-Mask-Making-in-Zambia/ Accessed June 18, 2020.

S300. U.S. Embassy Lusaka Zambia. Health Alert: Zambia, Wearing Of Masks In Public Now Mandatory. April 16, 2020. Available from:https://www.osac.gov/Country/Zambia/Content/Detail/Report/251986cc-38ee-4e4c-ad78-1875ef77459e Accessed June 18, 2020.

S301. Dirani TH. Zimbabwe Will Now Arrest Anyone Not Wearing Face Mask In Public During Lockdown. iHarare. May 3, 2020. Available from: https://iharare.com/people-not-wearing-masks-face-arrest/ Accessed July 14, 2020.

